# Associations of cannabis use, tobacco use and incident anxiety, mood, and psychotic disorders: a systematic review and meta-analysis

**DOI:** 10.1101/2023.03.17.23287299

**Authors:** Chloe Burke, Tom P Freeman, Hannah Sallis, Robyn E. Wootton, Annabel Burnley, Jonas Lange, Rachel Lees, Katherine Sawyer, Gemma Taylor

## Abstract

**Importance:** Traditional observational epidemiological studies have consistently found an association between tobacco use, cannabis use and subsequent mental ill-health. However, the extent to which this association reflects an increased risk of new-onset mental ill-health is unclear and may be biased by unmeasured confounding.

**Objective:** To examine the association between cannabis use, tobacco use and risk of incident mood, anxiety, and psychotic disorders, and explore risk of bias.

**Data Sources:** CINAHL, Embase, MEDLINE, PsycINFO and ProQuest Dissertation and Theses were searched from inception until November 2022, in addition to supplementary searches.

**Study Selection:** Longitudinal studies assessing tobacco use and cannabis use and their association with incident mood, anxiety or psychotic disorders were included. Studies conducted in populations selected on health status (e.g., pregnancy) or other highly-selected characteristics (e.g., incarcerated persons) were excluded.

**Data Extraction and Synthesis:** A modified Newcastle Ottawa Scale was used to assess study quality. The confounder matrix and E-Values were used to assess potential bias due to unmeasured confounding. Summary risk ratios (RR) were calculated in random-effects meta-analyses using the generic inverse variance method.

**Main Outcome(s) and Measure(s):** Exposures were measured via self-report and defined through status (e.g., current use) or heaviness of use (e.g., cigarettes per day). Outcomes were measured through symptom-based scales, interviews, registry codes and self-reported diagnosis or treatment. Effect estimates extracted were risk of incident disorders by exposure status.

**Results:** Seventy-five out of 27789 records were included. Random effects meta-analysis demonstrated a positive association between tobacco use and mood disorder (RR:1.39, 95%CI:1.30–1.47) and psychotic disorder (RR:3.45, 95%CI:2.63-4.53), but not anxiety disorder (RR:1.21, 95%CI:0.87–1.68). Cannabis use was positively associated with psychotic disorders (RR:3.19, 95%CI:2.07-4.90), but not mood disorders (RR:1.31, 95%CI:0.92-1.86) or anxiety disorders (RR:1.10, 95%CI:0.99-1.22). Confounder matrix and E-value assessment indicated estimates were moderately biased by unmeasured confounding.

**Conclusions and Relevance:** This systematic review and meta-analysis presents evidence for a longitudinal, positive association between both substances and incident psychotic disorders and tobacco use and mood disorders. There was no evidence to support an association between cannabis use and common mental health conditions. Existing evidence across all outcomes was limited by inadequate adjustment for potential confounders. Future research should prioritise methods allowing for stronger causal inference, such as Mendelian randomization and evidence triangulation.

## Introduction

Tobacco and cannabis are two of the most commonly used recreational drugs worldwide. In 2019, approximately 1.14 billion adults globally had smoked tobacco regularly and an estimated 200 million people used cannabis in the last year.^1^ Existing observational evidence demonstrates prospective associations between cannabis use, tobacco use and mental ill-health; including depression,^2–12^ anxiety,^7–10, 12–16^ and psychosis.^10, 17–26^ However, it remains unclear if the associations in question are causal or if they result from observational data biases (e.g., confounding, reverse causality).^27^ Numerous reviews of these substances and mental ill-health highlight confounding as a key limitation when interpreting results.^3–5, 9, 18, 19^ However, no comprehensive assessment of the strength of potential confounding bias has been conducted. For example, confounding can be reduced if appropriate controls are implemented (e.g., multivariable regression), but in-practice it is difficult to measure all confounders and without error.^28^

A further difficulty for tobacco and cannabis research is that co-use of these substances is highly common.^29–31^ Cannabis-tobacco co-use comprises both ‘concurrent use’ (i.e., use of both products in a pre-defined time period) and ‘co-administration’ (i.e., simultaneous use within the same delivery method).^31^ Considering the high co-occurrence and associations with mental ill-health, there has been debate as to which, if any, has a more important role to play in the development of subsequent mental illness.^32, 33^ To our knowledge, few reviews examining links with psychological outcomes have considered evidence for *both* substances independently,^8^ or jointly.^34–36^ These reviews have a range of limitations such as synthesising predominantly cross-sectional studies,^34, 35^ focusing on specific geographic regions or clinical populations,^8, 36^ lack of quality and confounding assessment.^34, 36^

As such, we aimed to synthesise longitudinal studies examining the association of cannabis and tobacco use with incident mental ill-health, with a focus on critically assessing biases that limit causal interpretation.

## Methods

We pre-registered our protocol on PROPSERO (CRD42021243903) and the Open Science Framework (https://osf.io/5t2pu/). Protocol changes have been reported in **eSupplement,** and we have followed MOOSE reporting guidelines (**eSupplement**).^37^

### Search and Selection

We searched CINAHL, Embase, MEDLINE, PsycINFO and ProQuest Dissertation and Theses from inception to November 2022. Searches were conducted using MeSH headings and text words relating to exposures, outcomes, and study design (**eSupplement**). CB and AB/RL/KS independently assessed title/abstracts and full texts. Discrepancies were resolved through discussion amongst the reviewers, or a third reviewer where necessary (GT).

### Eligibility Criteria

We included prospective longitudinal studies that (1) measured cannabis, tobacco, or co-use as an exposure, (2) used a ‘non-exposed’ comparator group, (3) reported a relevant effect estimate (e.g., risk ratio, odds ratio) and its variance, or necessary raw data. There were no restrictions on publication status, article language or publication date. To minimise risk of bias from reverse causation, we only included studies where participants with the outcome of interest were excluded at baseline (i.e., ‘incidence’). Studies were also excluded if participants were selected on a specific health status (e.g., pregnancy), or other highly selected characteristics (e.g., incarcerated persons). Full details in **eSupplement**.

### Data Extraction

Standardised forms were used to extract study information by two independent reviewers (CB and JL). A modified Newcastle Ottawa Scale (NOS) was used to evaluate study quality (**eSupplement**).^38^ The NOS evaluates studies across selection, comparability, and outcome assessment. A standardised assessment sheet was used (CB) and calibrated with a second-rater (JL) for ∼20% of the included studies. Disagreements were raised with an independent third reviewer (GT).

To explore the impact of bias due to unmeasured confounding across the included studies we used a combination of approaches. The E-value represents the minimum strength of association, on a risk ratio (RR) scale, an unmeasured confounder would need to have to fully explain a specific exposure–outcome association [i.e., fully reducing a RR to 1].^39^ What constitutes a small or large E-value is context dependent, relative to the exposure, outcome and measured covariates.^40^ To support interpretation of the E-values, we used a ‘confounder matrix’ assessment and a directed acyclic graph (DAG).^41^ The confounder matrix provides improved assessment and visualization of confounding control in reviews of observational studies. Based on our DAG (**eSupplement**), studies in the primary meta-analyses were assessed on adjustment for six constructs: other substance use; psychiatric comorbidity; socioeconomic status; sociodemographic factors; psychological factors; and other lifestyle factors (**eSupplement**).

### Statistical Analysis

We used RR and corresponding 95% confidence intervals (95%CIs), as the summary estimate. Included studies presented varied effect estimates and approach for conversion to RR is described in **eSupplement**. Adjusted and unadjusted, or minimally adjusted (i.e., age and sex), effect estimates were pooled separately. Random-effects meta-analysis using generic inverse variance approach was conducted. Between-study heterogeneity was explored through visual inspection of forest plots and tau-squared (*τ*^2^), and statistical inconsistency quantified using the *I*^2^ statistic.^42^ Prediction Intervals (PI) were additionally calculated i.e., 95% range of true effect estimates to be expected in exchangeable studies.^43^ Small-study effects were examined using Doi plots and asymmetry was quantified using the Luis Furuya-Kanamori (LFK) index.^44^ Where ≥10 studies were available, sources of heterogeneity in the primary analyses were explored through pre-planned subgroup analyses and meta-regressions.^42^ Additional exploratory sensitivity analyses were conducted for outliers and confounder matrix assessment. Meta-analyses were conducted in R, using the ‘meta’ package.^45^ Data and R scripts are available on GitHub (https://github.com/chloeeburke/tobcanmeta). The ‘E-Value’ online calculator (https://www.evalue-calculator.com/) and ‘metaconfoundr’^46^ package were used for sensitivity analyses.

## Results

### Literature Search

Of the 27789 records screened, 486 studies were retained for full-text screening (**eSupplement**). We identified 75 studies for inclusion,^13, 47–120^ of which 59 were included in the primary meta-analyses.^13, 47, 49, 50, 52, 53, 55–68, 71–73, 75–78, 80–82, 84, 85, 87–89, 91, 92, 94–96, 100, 102–118, 120^ A list of studies excluded at full text stage is available in **eSupplement**). Studies included in the primary meta-analyses consisted of 1733679 participants at risk of incident outcomes. Follow-up length ranged from 6-months to 63-years. Exposures were measured according to heaviness (e.g., cigarettes per day; k=28) or status of use (e.g., current use; k=31). Outcomes were assessed using symptom-based scales (k=21), interviews (k=18), registry codes (k=14), self-reported treatment/diagnosis (k=2) and composites (k=4). Study characteristics are presented in **eSupplement**.

### Meta-Analyses

#### Tobacco

Tobacco use was associated with incident mood disorders (K= 43; RR:1.39, 95%CI:1.30–1.47; I^2=^61.2%; *τ*^2=^0.014; PI: 1.08–1.77; **Figure 1**)^13, 47, 49, 50, 52, 53, 55–64, 68, 71–73, 75–78, 80, 81, 85, 87, 88, 91, 92, 100, 102, 103, 105–110, 114, 115, 118^. Exclusion of outliers,^49, 68, 73^ produced similar results (K=40; RR:1.38, 95%CI:1.31–1.45, I^2=^28.5%). Pooled unadjusted results yielded a larger point estimate (K=41; RR:1.47, 95%CI:1.34–1.60; I^2=^68.6%; *τ*^2=^0.06; PI: 0.92–2.33; **eSupplement**).^47, 48, 50–56, 58, 68–74, 77, 80, 81, 83, 85, 86, 88, 90, 92, 93, 97, 98, 102, 103, 105–108, 110, 114, 115, 118, 119^

**Figure 1.**
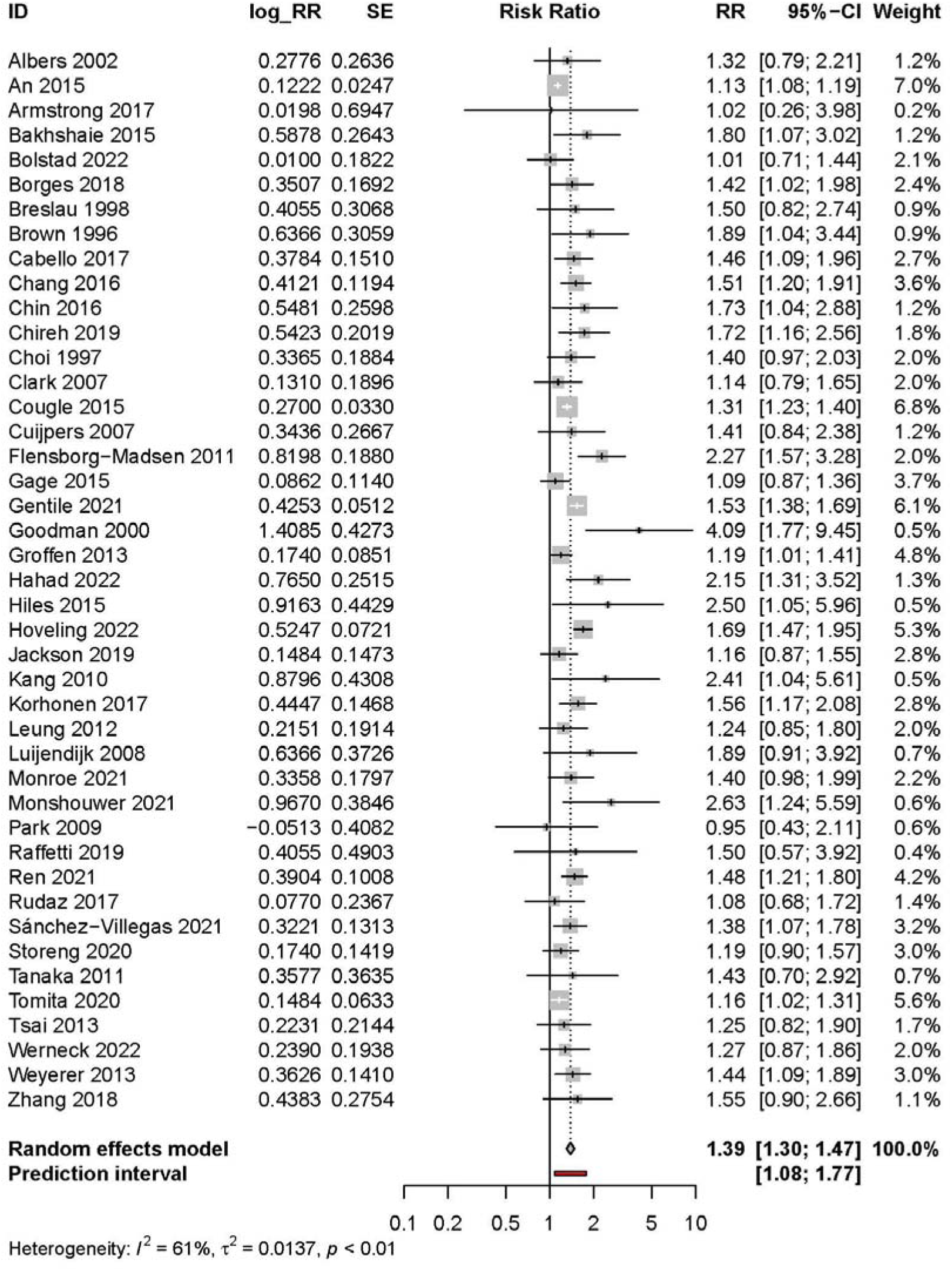
Meta-analysis of the association of tobacco use and mood disorders

Tobacco use was not associated with incident anxiety disorders (K=7; RR:1.21, 95%CI:0.87-1.68; I^2^=82.2%; *τ*^2^=0.143; PI: 0.42–3.50; **Figure 2**).^63, 64, 71, 76, 91, 92, 107^ Pooled unadjusted studies yielded a larger point estimate (K=8; RR:1.60, 95%CI:1.10–2.32; I^2^=71.7%; *τ*^2^=0.204; PI: 0.48–5.30; **eSupplement**).^53, 71, 79, 91, 92, 96, 107, 120^

**Figure 2.**
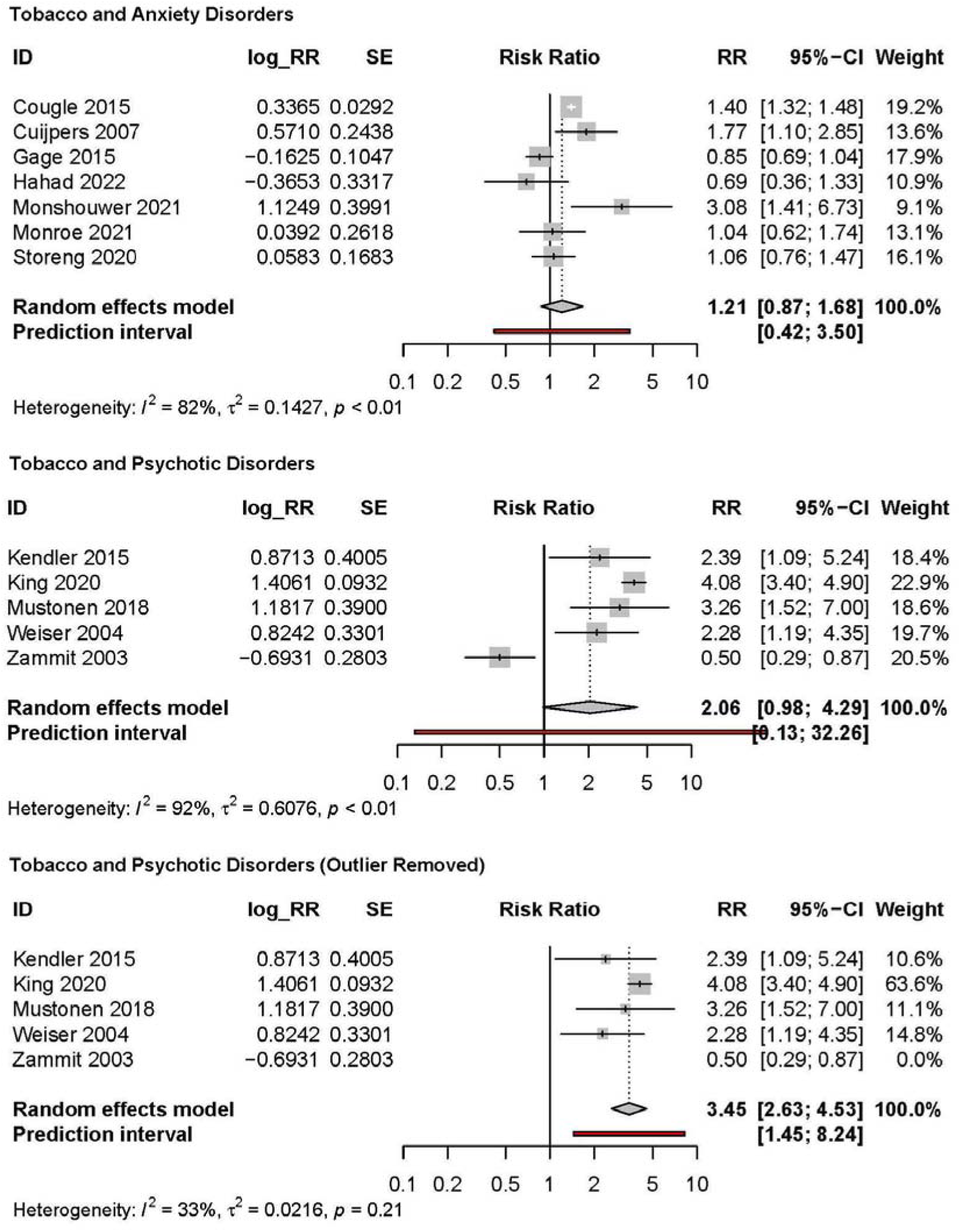
Meta-analyses of adjusted associations of tobacco use and anxiety and psychotic disorders

Tobacco use was not associated with incident psychotic disorders (K=5; RR:2.06, 95%CI:0.98-4.29; I^2^=92.3%; *τ*^2^=0.608; PI=0.13–32.26).^82, 84, 94, 113, 117^ Exclusion of one outlier,^117^ yielded a larger pooled estimate (RR:3.45, 95%CI:2.63–4.53, I^2^=32.9). As outlier identification was exploratory, pooled results with and without the outlier excluded are presented (**Figure 2**). Pooled unadjusted studies yielded a larger estimate (K=5; RR:3.12, 95%CI:1.67–5.81; I^2^=83%; **eSupplement**).^82, 84, 99, 113, 117^

### Cannabis

Cannabis use was not associated with incident mood disorders (K=7; RR:1.31, 95%CI:0.92-1.86; I^2^=77.0%; *τ*^2^=0.164; PI: 0.42-4.09; **Figure 3**).^65, 66, 71, 89, 96, 104, 111^ Pooled unadjusted studies yielded a larger estimate (K=7; RR:1.47 95%CI:1.19–1.81; I^2^=72.4%; **eSupplement**).^65, 66, 71, 89, 96, 101, 111^

**Figure 3.**
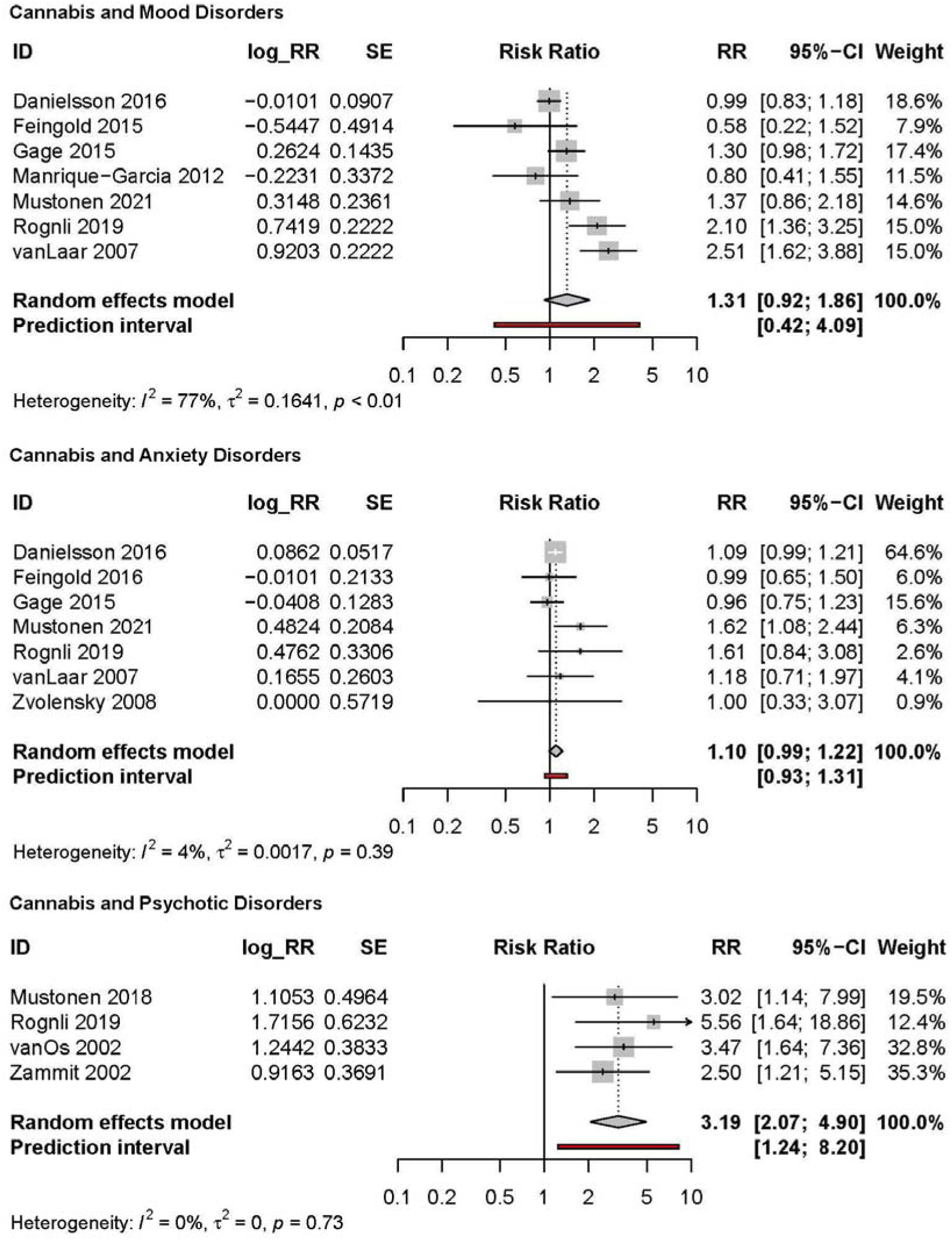
Meta-analyses of adjusted associations of cannabis use and mood, anxiety and psychotic disorders

Cannabis use was not associated with incident anxiety disorders (K=7; RR:1.10, 95%CI:0.99-1.22; I^2^=4.4%; *τ*^2^=0.002; PI: 0.93–1.31; **Figure 3**).^65, 67, 71, 96, 104, 111, 120^ Pooled unadjusted studies yielded a larger estimate (K=6; RR:1.51 95%CI:1.20–1.89; I^2^=74.3%; **eSupplement**).^65, 67, 71, 96, 111, 120^

Cannabis use was associated with incident psychotic disorders (K=4; RR:3.19, 95%CI:2.07–4.90; I^2^=0%; *τ*^2^=0.00; PI: 1.24–8.20; **Figure 3**).^95, 104, 112, 116^ Pooled unadjusted studies yielded a larger estimate (K=3; RR:4.68 95%CI:3.30–6.64; I^2^=0.0%; **eSupplement**).^95, 112, 117^

### Quality Assessment and Meta-Biases

Across studies included in the primary meta-analyses, roughly one quarter of studies (27%) were judged as ‘high’ quality (i.e., lower risk of bias) in the quality assessment (**eSupplement**), with an overall mean score of 7.35 (SD 1.01). The proportion of high quality studies differed by analysis (**eSupplement**). Many studies (58%) were marked down due to high attrition or insufficient information about loss to follow-up (e.g., differential attrition), and 41% of studies were marked down for ‘comparability’ (i.e., confounding bias). Using the confounder matrix, most studies had multiple confounding constructs rated as inadequately adjusted for (**eSupplement**), particularly other psychological factors (e.g., loneliness) and psychiatric comorbidity. Median E-values for study effect estimates and CIs have been presented in Table 1.

**Table 1.** E-value and confounder matrix summary

| Analysis Details |  |  |  | Median E-Value <sup>a</sup> |  | Summary Matrix (%) <sup>b</sup> |  |  |  |  |  |
| --- | --- | --- | --- | --- | --- | --- | --- | --- | --- | --- | --- |
| Exposure | Outcome | K | RR≤1 | PE | CI | SU | PC | SES | SD | PF | LF |
| Tobacco | Mood | 43 | 2% | 2.21 | 1.16 | 47 | 7 | 47 | 95 | 26 | 44 |
| Tobacco | Anxiety | 7 | 29% | 2.15 | 1.00 | 43 | 14 | 57 | 100 | 43 | 57 |
| Tobacco | Psychotic | 5 | 20% | 4.21 | 1.67 | 40 | 40 | 80 | 60 | 40 | 0 |
| Cannabis | Mood | 7 | 43% | 2.08 | 1.00 | 86 | 43 | 29 | 100 | 57 | 0 |
| Cannabis | Anxiety | 7 | 43% | 1.40 | 1.00 | 71 | 43 | 29 | 86 | 43 | 0 |
| Cannabis | Psychotic | 4 | 0% | 5.95 | 1.20 | 50 | 50 | 0 | 75 | 25 | 0 |

Abbreviation: RR = risk ratio; PE = pooled estimate; CI = confidence interval; SU = substance use; PC = psychiatric comorbidity; SES = socioeconomic status; SD = socio-demographics; PF = psychological factors; LF = lifestyle factors.
<sup>a</sup> For observed estimates below the null (RR<1) the inverse of the observed RR was taken prior to applying the formula and the upper limit (UL) of the 95%CI was considered. Where the UL of the 95%CI was >=1, E-values = 1, where the UL of the 95%CI was <1 then $UL^* = 1/UL$ and $E\text{-value} = UL + \sqrt{UL^* \times (UL - 1)}$ .
<sup>b</sup> Numbers denote the percentages of studies in the primary meta-analyses that were judged as adequately adjusting for the different constructs (i.e. SU, PC, SES, SD, PF, LF).

Evidence for small-study effects was present in all analyses, except cannabis and mood disorders, with Doi plots and LFK indices suggesting minor or major asymmetry (**eSupplement**).

### Subgroup and Sensitivity Analyses

Subgroup and sensitivity analyses were only performed for tobacco use and mood disorders due to low study numbers (K <10) in other meta-analyses. Results were examined across different age groups, follow-up length, sample size, study quality, confounding adjustment and exposure/outcome types. No analyses supported evidence of subgroup effects (**eSupplement**).

## Discussion

To our knowledge, this is the first systematic review and meta-analysis on the association of tobacco use, cannabis use and incident mental ill-health that has undertaken a comprehensive assessment of the influence of confounding bias, employing novel methods (i.e., confounder matrix, E-values) to support this assessment. We found evidence for positive associations of tobacco and incident mood and psychotic disorders, and of cannabis and incident psychotic disorders. Our review includes the first meta-analysis of the longitudinal association between tobacco use and anxiety disorders, and addresses limitations of previous reviews which have considered evidence for both substances in relation to psychological outcomes.

Accurately understanding the causal pathways between substance use and subsequent mental illness is crucial for informing the implementation of effective evidence-based public health policies.^121^ Results from this review are based on observational evidence and cannot in isolation be considered proof of causality. However, current tobacco policy and clinical implications regarding the association and between smoking and various physical conditions (e.g., lung cancer) is considered strong evidence of causality due to consistency in direction and strength of the effect estimate in combination with other criteria (e.g. dose-response). This study adds to a wider, growing body of evidence that these substances have a causal role in development of psychotic disorders, and tobacco use in mood disorders.^122, 123^ As such, policy makers should consider the level of evidence as adequate to inform public health campaigns (e.g. warning labels) about the potential harms of smoking for mental health.^76^ Nonetheless, there are still essential areas for future research to inform, including accurately identifying size of causal effects and possible biological mechanisms.^123^

While there was evidence for an association between tobacco and both mood and psychotic disorders, we did not find compelling evidence to suggest tobacco use is associated with incident anxiety disorders. Previous narrative syntheses found mixed evidence regarding the association between tobacco use and later anxiety.^7,12^ The effect size observed in the analysis of tobacco use and subsequent mood disorders is consistent with previous meta-analyses.^2, 3, 6, 8^ Although there was considerable methodological heterogeneity present across these studies, subgroup analyses of studies using different exposure and outcome definitions demonstrated similar associations. Importantly, lack of evidence for differences between subgroups does not automatically imply the effect is equivalent across subgroups, and specific subgroup analyses may have been affected by other sources of heterogeneity.

Comparatively fewer studies examined the relationship between cannabis use and incident mood disorders. Our analyses of cannabis use for subsequent mood and anxiety disorders did not support evidence of an increased risk in the cannabis use versus non-use groups. Several previous meta-analyses have reported mixed evidence regarding the association between cannabis use and elevated anxiety symptoms or disorder,^124^ and there are multiple meta-analyses of prospective studies which report a modest association between cannabis use and depressive symptoms or disorder.^124^ Three previous meta-analyses of prospective studies adjusting for baseline depression, found modest associations (OR range: 1.17-1.37)^4, 5, 8^ between cannabis use and subsequent depression. It’s possible that focusing on incident outcomes (vs. statistical adjustment) could explain the discrepancy in findings but may also relate to other differences in review criteria and content (e.g., adolescents only, number of studies). Recent reviews focusing on studies of cannabis frequency and potency suggest that more frequent use,^24^ and use of more potent forms,^125^ poses greater risk. However, due to limited study numbers and measurements, it was not feasible to investigate these potential moderators.

In line with other meta-analyses, this review reported evidence of a strong association between tobacco use, cannabis use and psychotic disorders.^10, 18, 19, 21, 22, 24^ Considerable uncertainty regarding the size of the association was indicated by confidence and prediction intervals. ‘Noisy’ effect estimates are common in the case of rare outcomes, due to lower statistical power. Pooling such effects in a meta-analysis can help yield a more precise estimate of the association of substance use with psychotic disorder, but this review included few studies. This is likely related to our exclusion of traditional case-control designs, which are well suited to the study of rare outcomes but are at increased risk of bias from retrospective recall and reverse causality.^126^ Lack of prospective research in this area has been previously highlighted.^127, 128^

We did not identify any eligible studies of cannabis-tobacco co-use. Assuming causality, dual use may place consumers at a higher risk of developing a mental health disorder than the independent use of either substance. There is a selection of cross-sectional research which indicates people who co-use have a higher prevalence of mental health disorders,^129, 130^ and levels of psychological distress.^131^ Some longitudinal evidence suggests co-use is associated with greater mental health symptoms,^132^ but prospective evidence in general population samples is lacking.

Analyses of small-study effects suggested possible risk of publication bias, with evidence of asymmetry for most meta-analyses. As such, pooled estimates may misrepresent the ‘true’ association. However, asymmetry can be driven by multiple factors (e.g., methodological heterogeneity) and may not represent publication bias.^133^ Furthermore, in the case of small study numbers (K<10), Doi plots and LFK index have advantages over traditional funnel plots in detecting asymmetry but may still misrepresent asymmetry.^44^

E-value and confounder matrix assessment suggested that many of the studies are at risk of confounding bias. Studies often inadequately adjusted for key confounding variables (e.g., ACEs). Previous reviews of these exposures have demonstrated moderate-strong associations with risk of substance use and various mental health outcomes (e.g., ACEs: OR_Smoking_ 2.82, OR_Depression,_ 4.40).^134^ Notably, subgroup analyses stratified by study quality and confounding adjustment suggested minimal impact on the tobacco and mood disorders pooled estimate. Combined with our focus on incidence, this implies stronger evidence of a causal effect of tobacco smoking for mood disorders. However, none of the extracted effect estimates adjusted for genetic vulnerability (e.g., polygenic risk) which alternative study designs (e.g., familial-based designs) suggest may play a substantial role in the observed associations.^135–137^ E-values must be interpreted considering some assumptions and limitations.^40, 138^ Importantly, adjustment for some covariates (e.g., SES) likely reduces bias from some unmeasured confounding (e.g., ACEs) due to the associations between these constructs. Nonetheless, the smaller E-values observed for some significant associations (i.e., tobacco/mood) would suggest that pooled estimates likely overestimate the size of effect. Furthermore, many studies were limited by inadequate description of attrition and few studies reported on individual-level missing data or used methods to account for this (e.g., multiple imputation). This directly contradicts recommendations by relevant reporting guidelines (e.g., STROBE)^139^ and hinders assessment of selection bias. It’s critical that future studies aiming to explore causal associations, provide more detailed descriptions of participant attrition and missing data and apply appropriate methods to reduce bias.^140^ Finally, although we focused on incidence, this does not exclude risk of bias from reverse causation as both psychotic and non-psychotic mental disorders do not have discrete onsets and subthreshold or prodromal symptoms at baseline may remain unaccounted for.^141^ As such, to support the identification of the size of causal effects, there is the need for further research focusing on addressing and exploring the biases that arise in traditional observational epidemiological studies.

Mendelian randomisation (MR) is one such method. MR uses genetic variation as an instrumental variable for an exposure to estimate causal effects that are more robust to reverse causality and confounding bias.^142^ A systematic review of MR studies investigating the causal relationships between substance use and mental health found evidence to support a bi-directional, increasing relationship between smoking and symptoms of depression, bipolar disorder and schizophrenia.^143^ Evidence regarding cannabis use and mental health was less conclusive, which may relate to lack of available frequency instruments.^143, 144^ Still, MR is “*far from a silver bullet*”^145^ and there are important limitations to be addressed through more advanced methods (e.g., multivariable MR), additional sensitivity tests (e.g., residual population stratification) and incorporation into planned triangulation frameworks,^27^ including triangulation with carefully planned longitudinal cohort analyses.^27, 143^ Widespread adoption of DAGs when selecting secondary data sources may yield useful insights as to whether research questions are feasibly explored within certain datasets.^40^ In combination with the need for well-controlled prospective longitudinal studies, more evidence using alternative study designs is required, as meta-analysis of the same study design may serve to amplify inherent biases.

### Limitations

Several important limitations need to be considered. All studies used self-report to define exposure status. This is not unusual in cohort studies but will result in measurement error that can bias effect estimates in the case of both differential and non-differential misclassification. Similarly, we included studies which used symptom-based scales, self-reported diagnosis and resource access (e.g., medication) which will introduce further measurement error. Most studies were based in high-income countries, and we restricted the review to include studies conducted in a broadly general population samples which reduces generalisability. The number of studies included in most meta-analyses was small and also prevented planned explorations of heterogeneity, which is highly recommended for syntheses of non-randomised studies.^146^ Finally, through analysing overarching diagnostic groups (e.g., mood disorders), relevant differences for individual disorders may be overlooked (e.g., bipolar disorder), which will be important to consider in exploring possible causal mechanisms (e.g., neuroadaptations in nicotinic pathways).^123^

## Conclusions

This systematic review and meta-analysis presents evidence for a longitudinal, positive association between both substances and incident psychotic disorders and tobacco use and mood disorders. In contrast to previous meta-analyses, there was no evidence to support an association between cannabis use and common mental health disorders. Existing evidence across all outcomes was limited by inadequate adjustment for potential confounders. Future research should prioritise methods allowing for stronger causal inference, such as Mendelian randomization and evidence triangulation.

## Supporting information

eSupplement

## Data Availability

All data for meta-analyses are available online at https://github.com/chloeeburke/tobcanmeta.

## Acknowledgements

**Author Contributions**: (1) Concept and design: CB, TF, HS, RW, GT; (2) Acquisition, analysis, or interpretation of data: CB, AB, JL, KS, RL, TF, HS, RW, GT; (3) Drafting of the manuscript: CB, AB, JL, KS, RL, TF, HS, RW, GT; (4) Statistical analysis: CB, GT, TF, HS, RW; (5) Obtained funding: CB; (6) Administrative, technical, or material support: CB, AB, JL, KS, RL; (7) Supervision: TF, HS, RW, GT

**Conflict of Interest Disclosures**: GT has previously received funding from Grand (Pfizer) for work not related to this project. CB, HS and RW have done paid consultancy work for Action on Smoking and Health (ASH) for work related to this project. The remaining authors have no conflicts of interest to declare.

**Funding/Support**: This study was supported by a Society for the Study of Addiction (SSA) PhD studentship awarded to CB.

**Role of Funder/Sponsor**: The funding organization had no role in the design and conduct of the study; collection, management, analysis, and interpretation of the data; preparation, review, or approval of the manuscript; and decision to submit the manuscript for publication.

## Notes

### Clinical Protocols

https://osf.io/5t2pu/

### Funding Statement

This work is primarily supported by a Society for the Study of Addiction PhD studentship awarded to CB.

## References

1. UNODC. *World Drug Report* 2020. United Nations Publication; 2021.

2. Chaiton MO, Cohen JE, O’Loughlin J, Rehm J. A systematic review of longitudinal studies on the association between depression and smoking in adolescents. BMC Public Health. 2009;9:356. doi:10.1186/1471-2458-9-356

3. Chaplin AB, Daniels NF, Ples D, et al. Longitudinal association between cardiovascular risk factors and depression in young people: a systematic review and meta-analysis of cohort studies. Psychol Med. Published online June 25, 2021:1–11. doi:10.1017/S0033291721002488

4. Lev-Ran S, Roerecke M, Le Foll B, George TP, McKenzie K, Rehm J. The association between cannabis use and depression: a systematic review and meta-analysis of longitudinal studies. Psychol Med. 2014;44(4):797–810. doi:10.1017/S0033291713001438

5. Gobbi G, Atkin T, Zytynski T, et al. Association of Cannabis Use in Adolescence and Risk of Depression, Anxiety, and Suicidality in Young Adulthood: A Systematic Review and Meta-analysis. JAMA Psychiatry. 2019;76(4):426–434. doi:10.1001/jamapsychiatry.2018.4500

6. Luger TM, Suls J, Vander Weg MW. How robust is the association between smoking and depression in adults? A meta-analysis using linear mixed-effects models. Addict Behav. 2014;39(10):1418–1429. doi:10.1016/j.addbeh.2014.05.011

7. Stevenson J, Miller CL, Martin K, Mohammadi L, Lawn S. Investigating the reciprocal temporal relationships between tobacco consumption and psychological disorders for youth: an international review. BMJ Open. 2022;12(6):e055499. doi:10.1136/bmjopen-2021-055499

8. Esmaeelzadeh S, Moraros J, Thorpe L, Bird Y. Examining the Association and Directionality between Mental Health Disorders and Substance Use among Adolescents and Young Adults in the U.S. and Canada—A Systematic Review and Meta-Analysis. J Clin Med. 2018;7(12):543. doi:10.3390/jcm7120543

9. Garey L, Olofsson H, Garza T, Rogers AH, Kauffman BY, Zvolensky MJ. Directional Effects of Anxiety and Depressive Disorders with Substance Use: a Review of Recent Prospective Research. Curr Addict Rep. 2020;7(3):344–355. doi:10.1007/s40429-020-00321-z

10. Moore THM, Zammit S, Lingford-Hughes A, et al. Cannabis use and risk of psychotic or affective mental health outcomes: a systematic review. Lancet Lond Engl. 2007;370(9584):319–328. doi:10.1016/S0140-6736(07)61162-3

11. Farooqui M, Shoaib S, Afaq H, et al. Bidirectionality of Smoking and Depression in Adolescents: a Systemic Review. Trends Psychiatry Psychother. Published online June 23, 2022. doi:10.47626/2237-6089-2021-0429

12. Fluharty M, Taylor AE, Grabski M, Munafò MR. The Association of Cigarette Smoking With Depression and Anxiety: A Systematic Review. Nicotine Tob Res Off J Soc Res Nicotine Tob. 2017;19(1):3–13. doi:10.1093/ntr/ntw140

13. Armstrong NM, Meoni LA, Carlson MC, et al. Cardiovascular risk factors and risk of incident depression throughout adulthood among men: The Johns Hopkins Precursors Study. J Affect Disord. 2017;214:60–66. doi:10.1016/j.jad.2017.03.004

14. Twomey CD. Association of cannabis use with the development of elevated anxiety symptoms in the general population: a meta-analysis. J Epidemiol Community Health. 2017;71(8):811–816. doi:10.1136/jech-2016-208145

15. Zimmermann M, Chong AK, Vechiu C, Papa A. Modifiable risk and protective factors for anxiety disorders among adults: A systematic review. Psychiatry Res. 2020;285:112705. doi:10.1016/j.psychres.2019.112705

16. Kedzior KK, Laeber LT. A positive association between anxiety disorders and cannabis use or cannabis use disorders in the general population- a meta-analysis of 31 studies. BMC Psychiatry. 2014;14(1):136. doi:10.1186/1471-244X-14-136

17. Godin SL, Shehata S. Adolescent cannabis use and later development of schizophrenia: An updated systematic review of longitudinal studies. J Clin Psychol. 2022;78(7):1331–1340. doi:10.1002/jclp.23312

18. Gurillo P, Jauhar S, Murray RM, MacCabe JH. Does tobacco use cause psychosis? Systematic review and meta-analysis. Lancet Psychiatry. 2015;2(8):718–725. doi:10.1016/S2215-0366(15)00152-2

19. Hunter A, Murray R, Asher L, Leonardi-Bee J. The Effects of Tobacco Smoking, and Prenatal Tobacco Smoke Exposure, on Risk of Schizophrenia: A Systematic Review and Meta-Analysis. Nicotine Tob Res. 2020;22(1):3–10. doi:10.1093/ntr/nty160

20. Le Bec PY, Fatséas M, Denis C, Lavie E, Auriacombe M. [Cannabis and psychosis: search of a causal link through a critical and systematic review]. L’Encephale. 2009;35(4):377–385. doi:10.1016/j.encep.2008.02.012

21. Marconi A, Di Forti M, Lewis CM, Murray RM, Vassos E. Meta-analysis of the Association Between the Level of Cannabis Use and Risk of Psychosis. Schizophr Bull. 2016;42(5):1262–1269. doi:10.1093/schbul/sbw003

22. Myles N, Newall HD, Curtis J, Nielssen O, Shiers D, Large M. Tobacco Use Before, At, and After First-Episode Psychosis: A Systematic Meta-Analysis. J Clin Psychiatry. 2012;73(4):21015. doi:10.4088/JCP.11r07222

23. Patel S, Khan S, M S, Hamid P. The Association Between Cannabis Use and Schizophrenia: Causative or Curative? A Systematic Review. Cureus. 2020;12(7). doi:10.7759/cureus.9309

24. Robinson T, Ali MU, Easterbrook B, Hall W, Jutras-Aswad D, Fischer B. Risk-thresholds for the association between frequency of cannabis use and the development of psychosis: a systematic review and meta-analysis. Psychol Med. Published online March 24, 2022:1–11. doi:10.1017/S0033291722000502

25. Semple DM, McIntosh AM, Lawrie SM. Cannabis as a risk factor for psychosis: systematic review. J Psychopharmacol (Oxf). 2005;19(2):187–194. doi:10.1177/0269881105049040

26. Arango C, Dragioti E, Solmi M, et al. Risk and protective factors for mental disorders beyond genetics: an evidence-based atlas. World Psychiatry. 2021;20(3):417–436. doi:10.1002/wps.20894

27. Hammerton G, Munafò MR. Causal inference with observational data: the need for triangulation of evidence. Psychol Med. 2021;51(4):563–578. doi:10.1017/S0033291720005127

28. Fewell Z, Davey Smith G, Sterne JAC. The impact of residual and unmeasured confounding in epidemiologic studies: a simulation study. Am J Epidemiol. 2007;166(6):646–655. doi:10.1093/aje/kwm165

29. Agrawal A, Budney AJ, Lynskey MT. The co-occurring use and misuse of cannabis and tobacco: a review. Addiction. 2012;107(7):1221–1233. doi:https://doi.org/10.1111/j.1360-0443.2012.03837.x

30. Gravely S, Driezen P, Smith DM, et al. International differences in patterns of cannabis use among adult cigarette smokers: Findings from the 2018 ITC Four Country Smoking and Vaping Survey. Int J Drug Policy. 2020;79:102754. doi:10.1016/j.drugpo.2020.102754

31. Hindocha C, McClure EA. Unknown population-level harms of cannabis and tobacco co-use: if you don’t measure it, you can’t manage it. Addiction. 2021;116(7):1622–1630. doi:10.1111/add.15290

32. Gage SH, Munafò MR. Smoking as a causal risk factor for schizophrenia. Lancet Psychiatry. 2015;2(9):778–779. doi:10.1016/S2215-0366(15)00333-8

33. Fergusson DM, Hall W, Boden JM, Horwood LJ. Rethinking cigarette smoking, cannabis use, and psychosis. Published online 2015. doi:10.1016/S2215-0366(15)00208-4

34. Peters EN, Budney AJ, Carroll KM. Clinical correlates of co-occurring cannabis and tobacco use: a systematic review. Addict Abingdon Engl. 2012;107(8):1404–1417. doi:10.1111/j.1360-0443.2012.03843.x

35. Ramo DE, Liu H, Prochaska JJ. Tobacco and marijuana use among adolescents and young adults: A systematic review of their co-use. Clin Psychol Rev. 2012;32(2):105–121. doi:10.1016/j.cpr.2011.12.002

36. Sabe M, Zhao N, Kaiser S. Cannabis, nicotine and the negative symptoms of schizophrenia: Systematic review and meta-analysis of observational studies. Neurosci Biobehav Rev. 2020;116:415–425. doi:10.1016/j.neubiorev.2020.07.007

37. Stroup DF, Berlin JA, Morton SC, et al. Meta-analysis of Observational Studies in EpidemiologyA Proposal for Reporting. JAMA. 2000;283(15):2008–2012. doi:10.1001/jama.283.15.2008

38. Wells G, Shea B, O’Connell D, et al. The Nwecastle-Ottawa Scale (NOS) for assessing the quality of nonrandomised studies in meta-analyses. Published 2013. Accessed February 12, 2021. http://www.ohri.ca/programs/clinical_epidemiology/oxford.asp

39. VanderWeele TJ, Ding P. Sensitivity Analysis in Observational Research: Introducing the E-Value. Ann Intern Med. 2017;167(4):268–274. doi:10.7326/M16-2607

40. VanderWeele TJ, Ding P, Mathur M. Technical Considerations in the Use of the E-Value. J Causal Inference. 2019;7(2). doi:10.1515/jci-2018-0007

41. Petersen JM, Barrett M, Ahrens KA, et al. The confounder matrix: A tool to assess confounding bias in systematic reviews of observational studies of etiology. Res Synth Methods. 2022;13(2):242–254. doi:10.1002/jrsm.1544

42. Higgins J, Thomas J, Chandler J, et al. Cochrane Handbook for Systematic Reviews of Interventions. Published 2020. Accessed February 12, 2021. www.training.cochrane.org/handbook

43. IntHout J, Ioannidis JPA, Rovers MM, Goeman JJ. Plea for routinely presenting prediction intervals in meta-analysis. BMJ Open. 2016;6(7):e010247. doi:10.1136/bmjopen-2015-010247

44. Furuya-Kanamori L, Barendregt JJ, Doi SAR. A new improved graphical and quantitative method for detecting bias in meta-analysis. Int J Evid Based Healthc. 2018;16(4):195–203. doi:10.1097/XEB.0000000000000141

45. Schwarzer G, Carpenter JR, Rücker G. metasens: Statistical Methods for Sensitivity Analysis in Meta-Analysis. Published online January 16, 2021. Accessed February 12, 2021. https://CRAN.R-project.org/package=metasens

46. Barrett M, Petersen JM, Trinquart L. metaconfoundr: Visualize “Confounder” Control in Meta-Analyses. Published online August 6, 2022. Accessed September 18, 2022. https://CRAN.R-project.org/package=metaconfoundr

47. Albers AB, Biener L. The Role of Smoking and Rebelliousness in the Development of Depressive Symptoms among a Cohort of Massachusetts Adolescents. Prev Med. 2002;34(6):625–631. doi:10.1006/pmed.2002.1029

48. Almeida OP, Hankey GJ, Yeap BB, Golledge J, McCaul K, Flicker L. A risk table to assist health practitioners assess and prevent the onset of depression in later life. Prev Med. 2013;57(6):878–882. doi:10.1016/j.ypmed.2013.09.021

49. An R, Xiang X. Smoking, heavy drinking, and depression among U.S. middle-aged and older adults. Prev Med. 2015;81:295–302. doi:10.1016/j.ypmed.2015.09.026

50. Bakhshaie J, Zvolensky MJ, Goodwin RD. Cigarette smoking and the onset and persistence of depression among adults in the United States: 1994–2005. Compr Psychiatry. 2015;60:142–148. doi:10.1016/j.comppsych.2014.10.012

51. Beutel ME, Brähler E, Wiltink J, et al. New onset of depression in aging women and men: contributions of social, psychological, behavioral, and somatic predictors in the community. Psychol Med. 2019;49(7):1148–1155. doi:10.1017/S0033291718001848

52. Bolstad I, Alakokkare AE, Bramness JG, et al. The relationships between use of alcohol, tobacco and coffee in adolescence and mood disorders in adulthood. Acta Psychiatr Scand. 2022;146(6):594–603. doi:10.1111/acps.13506

53. Borges G, Benjet C, Orozco R, Medina-Mora ME. A longitudinal study of reciprocal risk between mental and substance use disorders among Mexican youth. J Psychiatr Res. 2018;105:45–53. doi:10.1016/j.jpsychires.2018.08.014

54. Bots S, Tijhuis M, Giampaoli S, Kromhout D, Nissinen A. Lifestyle- and diet-related factors in late-life depression—a 5-year follow-up of elderly European men: the FINE study. Int J Geriatr Psychiatry. 2008;23(5):478–484. doi:10.1002/gps.1919

55. Breslau N, Peterson EL, Schultz LR, Chilcoat HD, Andreski P. Major Depression and Stages of Smoking: A Longitudinal Investigation. Arch Gen Psychiatry. 1998;55(2):161–166. doi:10.1001/archpsyc.55.2.161

56. Brown RA, Lewinsohn PM, Seeley JR, Wagner EF. Cigarette Smoking, Major Depression, and Other Psychiatric Disorders among Adolescents. J Am Acad Child Adolesc Psychiatry. 1996;35(12):1602–1610. doi:10.1097/00004583-199612000-00011

57. Cabello M, Miret M, Caballero FF, et al. The role of unhealthy lifestyles in the incidence and persistence of depression: a longitudinal general population study in four emerging countries. Glob Health. 2017;13(1):18. doi:10.1186/s12992-017-0237-5

58. Chang SC, Pan A, Kawachi I, Okereke OI. Risk factors for late-life depression: A prospective cohort study among older women. Prev Med. 2016;91:144–151. doi:10.1016/j.ypmed.2016.08.014

59. Chin WY, Wan EYF, Choi EPH, Chan KTY, Lam CLK. The 12-Month Incidence and Predictors of PHQ-9–Screened Depressive Symptoms in Chinese Primary Care Patients. Ann Fam Med. 2016;14(1):47–53. doi:10.1370/afm.1854

60. Chireh B, D’Arcy C. Shared and unique risk factors for depression and diabetes mellitus in a longitudinal study, implications for prevention: an analysis of a longitudinal population sample aged ⩾45 years. Ther Adv Endocrinol Metab. 2019;10:2042018819865828. doi:10.1177/2042018819865828

61. Choi WS, Patten CA, Gillin JC, Kaplan RM, Pierce JP. Cigarette smoking predicts development of depressive symptoms among U.S. Adolescents1,2. Ann Behav Med. 1997;19(1):42–50. doi:10.1007/BF02883426

62. Clark C, Haines MM, Head J, et al. Psychological symptoms and physical health and health behaviours in adolescents: a prospective 2-year study in East London. Addiction. 2007;102(1):126–135. doi:10.1111/j.1360-0443.2006.01621.x

63. Cougle JR, Hakes JK, Macatee RJ, Chavarria J, Zvolensky MJ. Quality of life and risk of psychiatric disorders among regular users of alcohol, nicotine, and cannabis: An analysis of the National Epidemiological Survey on Alcohol and Related Conditions (NESARC). J Psychiatr Res. 2015;66–67:135–141. doi:10.1016/j.jpsychires.2015.05.004

64. Cuijpers P, Smit F, Ten Have M, De Graaf R. Smoking is associated with first-ever incidence of mental disorders: a prospective population-based study. Addiction. 2007;102(8):1303–1309. doi:10.1111/j.1360-0443.2007.01885.x

65. Danielsson AK, Lundin A, Agardh E, Allebeck P, Forsell Y. Cannabis use, depression and anxiety: A 3-year prospective population-based study. J Affect Disord. 2016;193:103–108. doi:10.1016/j.jad.2015.12.045

66. Feingold D, Weiser M, Rehm J, Lev-Ran S. The association between cannabis use and mood disorders: A longitudinal study. J Affect Disord. 2015;172:211–218. doi:10.1016/j.jad.2014.10.006

67. Feingold D, Weiser M, Rehm J, Lev-Ran S. The association between cannabis use and anxiety disorders: Results from a population-based representative sample. Eur Neuropsychopharmacol. 2016;26(3):493–505. doi:10.1016/j.euroneuro.2015.12.037

68. Flensborg-Madsen T, Bay von Scholten M, Flachs EM, Mortensen EL, Prescott E, Tolstrup JS. Tobacco smoking as a risk factor for depression. A 26-year population-based follow-up study. J Psychiatr Res. 2011;45(2):143–149. doi:10.1016/j.jpsychires.2010.06.006

69. Fonseca LB, Pereira LP, Rodrigues PRM, et al. Incidence of depressive symptoms and its association with sociodemographic factors and lifestyle-related behaviors among Brazilian university students. Psychol Health Med. 2022;27(6):1311–1325. doi:10.1080/13548506.2021.1874432

70. Ford DE, Mead LA, Chang PP, Cooper-Patrick L, Wang NY, Klag MJ. Depression is a risk factor for coronary artery disease in men: The precursors study. Arch Intern Med. 1998;158(13):1422–1426. doi:10.1001/archinte.158.13.1422

71. Gage SH, Hickman M, Heron J, et al. Associations of Cannabis and Cigarette Use with Depression and Anxiety at Age 18: Findings from the Avon Longitudinal Study of Parents and Children. PLOS ONE. 2015;10(4):e0122896. doi:10.1371/journal.pone.0122896

72. Gentile A, Bianco A, Nordstrӧm A, Nordstrӧm P. Use of alcohol, drugs, inhalants, and smoking tobacco and the long-term risk of depression in men: A nationwide Swedish cohort study from 1969–2017. Drug Alcohol Depend. 2021;221:108553. doi:10.1016/j.drugalcdep.2021.108553

73. Goodman E, Capitman J. Depressive Symptoms and Cigarette Smoking Among Teens. Pediatrics. 2000;106(4):748–755. doi:10.1542/peds.106.4.748

74. Goodwin RD, Prescott M, Tamburrino M, Calabrese JR, Liberzon I, Galea S. Smoking is a predictor of depression onset among National Guard soldiers. Psychiatry Res. 2013;206(2):321–323. doi:10.1016/j.psychres.2012.11.025

75. Groffen DAI, Koster A, Bosma H, et al. Unhealthy Lifestyles Do Not Mediate the Relationship Between Socioeconomic Status and Incident Depressive Symptoms: The Health ABC study. Am J Geriatr Psychiatry. 2013;21(7):664–674. doi:10.1016/j.jagp.2013.01.004

76. Hahad O, Beutel M, Gilan DA, et al. The association of smoking and smoking cessation with prevalent and incident symptoms of depression, anxiety, and sleep disturbance in the general population. J Affect Disord. 2022;313:100–109. doi:10.1016/j.jad.2022.06.083

77. Hiles SA, Baker AL, de Malmanche T, McEvoy M, Boyle M, Attia J. Unhealthy lifestyle may increase later depression via inflammation in older women but not men. J Psychiatr Res. 2015;63:65–74. doi:10.1016/j.jpsychires.2015.02.010

78. Hoveling LA, Liefbroer AC, Schweren LJS, Bültmann U, Smidt N. Socioeconomic differences in major depressive disorder onset among adults are partially explained by lifestyle factors: A longitudinal analysis of the Lifelines Cohort Study. J Affect Disord. 2022;314:309–317. doi:10.1016/j.jad.2022.06.018

79. Isensee B, Wittchen HU, Stein MB, Höfler M, Lieb R. Smoking Increases the Risk of Panic: Findings From a Prospective Community Study. Arch Gen Psychiatry. 2003;60(7):692–700. doi:10.1001/archpsyc.60.7.692

80. Jackson SE, Brown J, Ussher M, Shahab L, Steptoe A, Smith L. Combined health risks of cigarette smoking and low levels of physical activity: a prospective cohort study in England with 12-year follow-up. BMJ Open. 2019;9(11):e032852. doi:10.1136/bmjopen-2019-032852

81. Kang E, Lee J. A longitudinal study on the causal association between smoking and depression. J Prev Med Public Health Yebang Uihakhoe Chi. 2010;43(3):193–204. doi:10.3961/jpmph.2010.43.3.193

82. Kendler KS, Lönn SL, Sundquist J, Sundquist K. Smoking and Schizophrenia in Population Cohorts of Swedish Women and Men: A Prospective Co-Relative Control Study. Am J Psychiatry. 2015;172(11):1092–1100. doi:10.1176/appi.ajp.2015.15010126

83. Kim GE, Kim M ho, Lim WJ, Kim SI. The effects of smoking habit change on the risk of depression–Analysis of data from the Korean National Health Insurance Service. J Affect Disord. 2022;302:293–301. doi:10.1016/j.jad.2022.01.095

84. King M, Jones R, Petersen I, Hamilton F, Nazareth I. Cigarette smoking as a risk factor for schizophrenia or all non-affective psychoses. Psychol Med. 2021;51(8):1373–1381. doi:10.1017/S0033291720000136

85. Korhonen T, Ranjit A, Tuulio-Henriksson A, Kaprio J. Smoking status as a predictor of antidepressant medication use. J Affect Disord. 2017;207:221–227. doi:10.1016/j.jad.2016.09.035

86. Lam TH, Stewart SM, Ho SY, et al. Depressive symptoms and smoking among Hong Kong Chinese adolescents. Addiction. 2005;100(7):1003–1011. doi:10.1111/j.1360-0443.2005.01092.x

87. Leung J, Gartner C, Hall W, Lucke J, Dobson A. A longitudinal study of the bi-directional relationship between tobacco smoking and psychological distress in a community sample of young Australian women. Psychol Med. 2012;42(6):1273–1282. doi:10.1017/S0033291711002261

88. Luijendijk HJ, Stricker BH, Hofman A, Witteman JCM, Tiemeier H. Cerebrovascular risk factors and incident depression in community-dwelling elderly. Acta Psychiatr Scand. 2008;118(2):139–148. doi:10.1111/j.1600-0447.2008.01189.x

89. Manrique-Garcia E, Zammit S, Dalman C, Hemmingsson T, Allebeck P. Cannabis use and depression: a longitudinal study of a national cohort of Swedish conscripts. BMC Psychiatry. 2012;12(1):112. doi:10.1186/1471-244X-12-112

90. Meng X, Brunet A, Turecki G, Liu A, D’Arcy C, Caron J. Risk factor modifications and depression incidence: a 4-year longitudinal Canadian cohort of the Montreal Catchment Area Study. BMJ Open. 2017;7(6):e015156. doi:10.1136/bmjopen-2016-015156

91. Monroe DC, McDowell CP, Kenny RA, Herring MP. Dynamic associations between anxiety, depression, and tobacco use in older adults: Results from The Irish Longitudinal Study on Ageing. J Psychiatr Res. 2021;139:99–105. doi:10.1016/j.jpsychires.2021.05.017

92. Monshouwer K, ten Have M, de Graaf R, Blankers M, van Laar M. Tobacco Smoking and the Association With First Incidence of Mood, Anxiety, and Substance Use Disorders: A 3-Year Prospective Population-Based Study. Clin Psychol Sci. 2021;9(3):403–412. doi:10.1177/2167702620959287

93. Murphy JM, Horton NJ, Monson RR, Laird NM, Sobol AM, Leighton AH. Cigarette Smoking in Relation to Depression: Historical Trends From the Stirling County Study. Am J Psychiatry. 2003;160(9):1663–1669. doi:10.1176/appi.ajp.160.9.1663

94. Mustonen A, Ahokas T, Nordström T, et al. Smokin‘ hot: adolescent smoking and the risk of psychosis. Acta Psychiatr Scand. 2018;138(1):5–14. doi:10.1111/acps.12863

95. Mustonen A, Niemelä S, Nordström T, et al. Adolescent cannabis use, baseline prodromal symptoms and the risk of psychosis. Br J Psychiatry. 2018;212(4):227–233. doi:10.1192/bjp.2017.52

96. Mustonen A, Hielscher E, Miettunen J, et al. Adolescent cannabis use, depression and anxiety disorders in the Northern Finland Birth Cohort 1986. BJPsych Open. 2021;7(4):e137. doi:10.1192/bjo.2021.967

97. Najafipour H, Shahrokhabadi MS, Banivaheb G, Sabahi A, Shadkam M, Mirzazadeh A. Trends in the prevalence and incidence of anxiety and depressive symptoms in Iran: findings from KERCADRS. Fam Med Community Health. 2021;9(3):e000937. doi:10.1136/fmch-2021-000937

98. do Nascimento KKF, Pereira KS, Firmo JOA, Lima-Costa MF, Diniz BS, Castro-Costa E. Predictors of incidence of clinically significant depressive symptoms in the elderly: 10-year follow-up study of the Bambui cohort study of aging. Int J Geriatr Psychiatry. 2015;30(12):1171–1176. doi:10.1002/gps.4271

99. Self-reported premorbid health in 15 individuals who later developed schizophrenia compared with healthy controls: Prospective data from the Young-HUNT1 Survey (The HUNT Study). Psykologisk.no. Published November 28, 2018. Accessed March 14, 2023. https://psykologisk.no/sp/2018/11/e8/

100. Park S. The Causal Association Between Smoking and Depression Among South Korean Adolescents. J Addict Nurs. 2009;20(2):93–103. doi:10.1080/10884600902850111

101. Paton S, Kessler R, Kandel D. Depressive Mood and Adolescent Illicit Drug Use: A Longitudinal Analysis. J Genet Psychol. 1977;131(2):267–289. doi:10.1080/00221325.1977.10533299

102. Raffetti E, Donato F, Forsell Y, Galanti MR. Longitudinal association between tobacco use and the onset of depressive symptoms among Swedish adolescents: the Kupol cohort study. Eur Child Adolesc Psychiatry. 2019;28(5):695–704. doi:10.1007/s00787-018-1237-6

103. Ren X, Wang S, He Y, et al. Chronic Lung Diseases and the Risk of Depressive Symptoms Based on the China Health and Retirement Longitudinal Study: A Prospective Cohort Study. Front Psychol. 2021;12. Accessed March 14, 2023. https://www.frontiersin.org/articles/10.3389/fpsyg.2021.585597

104. Rognli EB, Bramness JG, von Soest T. Cannabis use in early adulthood is prospectively associated with prescriptions of antipsychotics, mood stabilizers, and antidepressants. Acta Psychiatr Scand. 2020;141(2):149–156. doi:10.1111/acps.13104

105. Rudaz DA, Vandeleur CL, Gebreab SZ, et al. Partially distinct combinations of psychological, metabolic and inflammatory risk factors are prospectively associated with the onset of the subtypes of Major Depressive Disorder in midlife. J Affect Disord. 2017;222:195–203. doi:10.1016/j.jad.2017.07.016

106. Sánchez-Villegas A, Gea A, Lahortiga-Ramos F, Martínez-González J, Molero P, Martínez-González MÁ. Bidirectional association between tobacco use and depression risk in the SUN cohort study. Adicciones. 2021;0(0):1725. doi:10.20882/adicciones.1725

107. Storeng SH, Sund ER, Krokstad S. Prevalence, clustering and combined effects of lifestyle behaviours and their association with health after retirement age in a prospective cohort study, the Nord-Trøndelag Health Study, Norway. BMC Public Health. 2020;20(1):900. doi:10.1186/s12889-020-08993-y

108. Tanaka H, Sasazawa Y, Suzuki S, Nakazawa M, Koyama H. Health status and lifestyle factors as predictors of depression in middle-aged and elderly Japanese adults: a seven-year follow-up of the Komo-Ise cohort study. BMC Psychiatry. 2011;11(1):20. doi:10.1186/1471-244X-11-20

109. Tomita A, Manuel JI. Evidence on the Association Between Cigarette Smoking and Incident Depression From the South African National Income Dynamics Study 2008–2015: Mental Health Implications for a Resource-Limited Setting. Nicotine Tob Res. 2020;22(1):118–123. doi:10.1093/ntr/nty163

110. Tsai AC, Chi SH, Wang JY. Cross-sectional and longitudinal associations of lifestyle factors with depressive symptoms in ≥53-year old Taiwanese — Results of an 8-year cohort study. Prev Med. 2013;57(2):92–97. doi:10.1016/j.ypmed.2013.04.021

111. Van Laar M, Van Dorsselaer S, Monshouwer K, De Graaf R. Does cannabis use predict the first incidence of mood and anxiety disorders in the adult population? Addiction. 2007;102(8):1251–1260. doi:10.1111/j.1360-0443.2007.01875.x

112. van Os J, Bak M, Hanssen M, Bijl RV, de Graaf R, Verdoux H. Cannabis Use and Psychosis: A Longitudinal Population-based Study. Am J Epidemiol. 2002;156(4):319–327. doi:10.1093/aje/kwf043

113. Weiser M, Reichenberg A, Grotto I, et al. Higher Rates of Cigarette Smoking in Male Adolescents Before the Onset of Schizophrenia: A Historical-Prospective Cohort Study. Am J Psychiatry. 2004;161(7):1219–1223. doi:10.1176/appi.ajp.161.7.1219

114. Werneck AO, Vancampfort D, Stubbs B, et al. Prospective associations between multiple lifestyle behaviors and depressive symptoms. J Affect Disord. 2022;301:233–239. doi:10.1016/j.jad.2021.12.131

115. Weyerer S, Eifflaender-Gorfer S, Wiese B, et al. Incidence and predictors of depression in non-demented primary care attenders aged 75 years and older: results from a 3-year follow-up study. Age Ageing. 2013;42(2):173–180. doi:10.1093/ageing/afs184

116. Zammit S, Allebeck P, Andreasson S, Lundberg I, Lewis G. Self reported cannabis use as a risk factor for schizophrenia in Swedish conscripts of 1969: historical cohort study. BMJ. 2002;325(7374):1199. doi:10.1136/bmj.325.7374.1199

117. Zammit S, Allebeck P, Dalman C, Lundberg I, Hemmingsson T, Lewis G. Investigating the Association Between Cigarette Smoking and Schizophrenia in a Cohort Study. Am J Psychiatry. 2003;160(12):2216–2221. doi:10.1176/appi.ajp.160.12.2216

118. Zhang XC, Woud ML, Becker ES, Margraf J. Do health-related factors predict major depression? A longitudinal epidemiologic study. Clin Psychol Psychother. 2018;25(3):378–387. doi:10.1002/cpp.2171

119. Zimmerman JA, Mast BT, Miles T, Markides KS. Vascular risk and depression in the Hispanic Established Population for the Epidemiologic Study of the Elderly (EPESE). Int J Geriatr Psychiatry. 2009;24(4):409–416. doi:10.1002/gps.2136

120. Zvolensky MJ, Lewinsohn P, Bernstein A, et al. Prospective associations between cannabis use, abuse, and dependence and panic attacks and disorder. J Psychiatr Res. 2008;42(12):1017–1023. doi:10.1016/j.jpsychires.2007.10.012

121. Taylor GMJ, Treur JL. An application of the stress-diathesis model: A review about the association between smoking tobacco, smoking cessation, and mental health. Int J Clin Health Psychol. 2023;23(1):100335. doi:10.1016/j.ijchp.2022.100335

122. Munafò MR. Growing Evidence for a Causal Role for Smoking in Mental Health. Nicotine Tob Res. 2022;24(5):631–632. doi:10.1093/ntr/ntac027

123. Firth J, Wootton RE, Sawyer C, Taylor GM. Clearing the air: clarifying the causal role of smoking in mental illness. World Psychiatry. 2023;22(1):151–152. doi:10.1002/wps.21023

124. Hall W, Leung J, Lynskey M. The Effects of Cannabis Use on the Development of Adolescents and Young Adults. Annu Rev Dev Psychol. 2020;2(1):461–483. doi:10.1146/annurev-devpsych-040320-084904

125. Petrilli K, Ofori S, Hines L, Taylor G, Adams S, Freeman TP. Association of cannabis potency with mental ill health and addiction: a systematic review. Lancet Psychiatry. 2022;9(9):736–750. doi:10.1016/S2215-0366(22)00161-4

126. Rothman K, Lash T, VanderWeele T, Haneuse S. Modern Epidemiology. Vol 63. Fourth Edi. Wolters Kluwer; 2021.

127. Sideli L, Quigley H, La Cascia C, Murray RM. Cannabis Use and the Risk for Psychosis and Affective Disorders. J Dual Diagn. 2020;16(1):22–42. doi:10.1080/15504263.2019.1674991

128. Quigley H, MacCabe JH. The relationship between nicotine and psychosis. Ther Adv Psychopharmacol. 2019;9:2045125319859969. doi:10.1177/2045125319859969

129. Hindocha C, Brose LS, Walsh H, Cheeseman H. Cannabis use and co-use in tobacco smokers and non-smokers: prevalence and associations with mental health in a cross-sectional, nationally representative sample of adults in Great Britain, 2020. Addiction. 2021;116(8):2209–2219. doi:10.1111/add.15381

130. Peters EN, Schwartz RP, Wang S, O’Grady KE, Blanco C. Psychiatric, Psychosocial, and Physical Health Correlates of Co-Occurring Cannabis Use Disorders and Nicotine Dependence. Drug Alcohol Depend. 2014;0. doi:10.1016/j.drugalcdep.2013.10.003

131. Wang N, Yao T, Sung HY, Max W. The Association of Cannabis Use and Cigarette Smoking with Psychological Distress Among Adults in California. Subst Use Misuse. 2022;57(2):193–201. doi:10.1080/10826084.2021.1995758

132. Tucker JS, Pedersen ER, Seelam R, Dunbar MS, Shih RA, D’Amico EJ. Types of Cannabis and Tobacco/Nicotine Co-Use and Associated Outcomes in Young Adulthood. Psychol Addict Behav J Soc Psychol Addict Behav. 2019;33(4):401–411. doi:10.1037/adb0000464

133. Sterne JAC, Harbord RM. Funnel Plots in Meta-analysis. Stata J. 2004;4(2):127–141. doi:10.1177/1536867X0400400204

134. Hughes K, Bellis MA, Hardcastle KA, et al. The effect of multiple adverse childhood experiences on health: a systematic review and meta-analysis. Lancet Public Health. 2017;2(8):e356–e366. doi:10.1016/S2468-2667(17)30118-4

135. Ranjit A, Korhonen T, Buchwald J, et al. Testing the reciprocal association between smoking and depressive symptoms from adolescence to adulthood: A longitudinal twin study. Drug Alcohol Depend. 2019;200:64–70. doi:10.1016/j.drugalcdep.2019.03.012

136. Barkhuizen W, Taylor MJ, Freeman D, Ronald A. A Twin Study on the Association Between Psychotic Experiences and Tobacco Use During Adolescence. J Am Acad Child Adolesc Psychiatry. 2019;58(2):267–276.e8. doi:10.1016/j.jaac.2018.06.037

137. Schaefer JD, Hamdi NR, Malone SM, et al. Associations between adolescent cannabis use and young-adult functioning in three longitudinal twin studies. Proc Natl Acad Sci. 2021;118(14):e2013180118. doi:10.1073/pnas.2013180118

138. VanderWeele TJ. Are Greenland, Ioannidis and Poole opposed to the Cornfield conditions? A defence of the E-value. Int J Epidemiol. 2022;51(2):364–371. doi:10.1093/ije/dyab218

139. Vandenbroucke JP, von Elm E, Altman DG, et al. Strengthening the Reporting of Observational Studies in Epidemiology (STROBE): Explanation and Elaboration. PLoS Med. 2007;4(10):e297. doi:10.1371/journal.pmed.0040297

140. VanderWeele TJ. Can Sophisticated Study Designs With Regression Analyses of Observational Data Provide Causal Inferences? JAMA Psychiatry. 2021;78(3):244–246. doi:10.1001/jamapsychiatry.2020.2588

141. Patten SB. Cannabis and non-psychotic mental disorders. Curr Opin Psychol. 2021;38:61-66. doi:10.1016/j.copsyc.2020.09.006

142. Davies NM, Holmes MV, Smith GD. Reading Mendelian randomisation studies: a guide, glossary, and checklist for clinicians. BMJ. 2018;362:k601. doi:10.1136/bmj.k601

143. Treur JL, Munafò MR, Logtenberg E, Wiers RW, Verweij KJH. Using Mendelian randomization analysis to better understand the relationship between mental health and substance use: a systematic review. Psychol Med. 2021;51(10):1593–1624. doi:10.1017/S003329172100180X

144. Hines LA, Treur JL, Jones HJ, Sallis HM, Munafò MR. Using genetic information to inform policy on cannabis. Lancet Psychiatry. 2020;7(12):1002–1003. doi:10.1016/S2215-0366(20)30377-1

145. Wootton RE, Jones HJ, Sallis HM. Mendelian randomisation for psychiatry: how does it work, and what can it tell us? Mol Psychiatry. Published online June 4, 2021:1–5. doi:10.1038/s41380-021-01173-3

146. Egger M, Higgins JPT, Smith GD. Systematic Reviews in Health Research: Meta-Analysis in Context. John Wiley & Sons; 2022.

