## Supplementary material for "Associations of cannabis use, tobacco use and incident anxiety, mood, and psychotic disorders: a systematic review and meta-analysis": eSupplement

### Supplemental Online Content

**eMethods 1.** MOOSE checklist

**MOOSE Checklist for Meta-analyses of Observational Studies**[1]

| Item No | Recommendation | Reported |
|---|---|---|
| Reporting of background should include | | |
| 1 | Problem definition | Introduction |
| 2 | Hypothesis statement | Introduction |
| 3 | Description of study outcome(s) | Eligibility Criteria |
| 4 | Type of exposure or intervention used | Eligibility Criteria |
| 5 | Type of study designs used | Eligibility Criteria |
| 6 | Study population | Eligibility Criteria |
| Reporting of search strategy should include | | |
| 7 | Qualifications of searchers (eg, librarians and investigators) | eMethods |
| 8 | Search strategy, including time period included in the synthesis and key words | eMethods |
| 9 | Effort to include all available studies, including contact with authors | eMethods |
| 10 | Databases and registries searched | eMethods |
| 11 | Search software used, name and version, including special features used (eg, explosion) | eMethods |
| 12 | Use of hand searching (eg, reference lists of obtained articles) | eMethods |
| 13 | List of citations located and those excluded, including justification | eTable1 |
| 14 | Method of addressing articles published in languages other than English | eMethods |
| 15 | Method of handling abstracts and unpublished studies | eMethods |
| 16 | Description of any contact with authors | eMethods |
| Reporting of methods should include | | |
| 17 | Description of relevance or appropriateness of studies assembled for assessing the hypothesis to be tested | Eligibility Criteria |
| 18 | Rationale for the selection and coding of data (eg, sound clinical principles or convenience) | Eligibility Criteria |
| 19 | Documentation of how data were classified and coded (eg, multiple raters, blinding and interrater reliability) | Search and Selection; Data Extraction |
| 20 | Assessment of confounding (eg, comparability of cases and controls in studies where appropriate) | eMethods |
| 21 | Assessment of study quality, including blinding of quality assessors, stratification or regression on possible predictors of study results | Data Extraction; eMethods |
| 22 | Assessment of heterogeneity | Statistical Analysis |
| 23 | Description of statistical methods (eg, complete description of fixed or random effects models, justification of whether the chosen models account for predictors of study results, dose-response models, or cumulative meta-analysis) in sufficient detail to be replicated | Statistical Analysis, eMethods |

| | | |
|---|---|---|
| 24 | Provision of appropriate tables and graphics | Results; Supplement |
| Reporting of results should include | | |
| 25 | Graphic summarizing individual study estimates and overall estimate | Figure 1-3 |
| 26 | Table giving descriptive information for each study included | eTable 2-7 |
| 27 | Results of sensitivity testing (eg, subgroup analysis) | eTable |
| 28 | Indication of statistical uncertainty of findings | Results (CIs and PIs) |
| Reporting of discussion should include | | |
| 29 | Quantitative assessment of bias (eg, publication bias) | Meta-Biases; eFigure 10-15; Discussion |
| 30 | Justification for exclusion (eg, exclusion of non-English language citations) | Eligibility Criteria; Discussion |
| 31 | Assessment of quality of included studies | eTable 8 + 9 |
| Reporting of conclusions should include | | |
| 32 | Consideration of alternative explanations for observed results | Discussion |
| 33 | Generalization of the conclusions (ie, appropriate for the data presented and within the domain of the literature review) | Discussion |
| 34 | Guidelines for future research | Discussion |
| 35 | Disclosure of funding source | Acknowledgements |

**eMethods 2.** Documentation of deviations from the pre-registered protocol

Summary of protocol deviations informed by suggested structure[2] for reporting available online at: https://osf.io/q8stz/

**Deviation (1):** Eligibility criteria – population characteristic

| | |
|---|---|
| Description: | Change in inclusion criteria made during title/abstract screening to exclude studies where participants were selected based on health status (e.g. pregnant women, asthmatic) or other clinically meaningful characteristics (e.g. incarcerated persons). |
| Justification: | This amendment was made to reduce the substantial heterogeneity in populations, and was informed by recommendations from a previous review in this area which cites highly specific samples as a barrier to interpretation of evidence synthesis[3] |
| Impact: | **Major**; reduced the total number of included studies and applicability of the review to other populations i.e. does the association of smoking with subsequent mental illness differ in different populations? Furthermore, evidence of positive association across heterogeneous populations would have added further evidence to assessment of causality. However, also enabled a more targeted assessment of confounding bias in 'general population' studies. |

**Deviation (2):** Search strategy – citation searching tool

| | |
|---|---|
| Description: | A citation chasing tool not pre-specified in the protocol was used for forward and backward citation searching (https://estech.shinyapps.io/citationchaser/). |
| Justification: | The tool was not publicly available at the time of writing the pre-registration. |
| Impact: | **Minor**; may have yielded a more comprehensive search of records than achieved through standard manual approach, although also possible that some records missed due to software reliance on Lens.org database to compile all reference and/or citing records. These supplementary references were screened by one reviewer due to resource availability, which may have increased possible biases in screening. |

**Deviation (3):** Data synthesis – meta-analysis method

| | |
|---|---|
| Description: | Change in pre-planned random-effects meta-analysis method from using Mantel-Haenszel (MH) method, to generic inverse variance method (GIV). |
| Justification: | MH method was originally selected due to recommendation by Cochrane Handbook as optimal approach for binary outcome data with few events.[4] However, it was discovered after protocol registration that this method is only applicable to raw data, rather than pre-calculated estimates - which was the target of the primary meta-analysis of adjusted effect estimates. |
| Impact: | **Minor**; still random-effects (i.e. assumption true effect varies from study to study) and GIV is a commonly implemented approach for random-effects meta-analysis. Under this method, the weight given to each study is the inverse of the variance of the effect estimate, which may have made the pooled estimate more precise. However, importantly, the synthesis of pre-calculated effect estimates would not have been possible using the MH method. |

**Deviation (4):** Data synthesis – unadjusted/minimally-adjusted

| | |
|---|---|
| Description: | Change from pooling unadjusted and adjusted estimates separately, to pooling 'minimally-adjusted' (i.e. age/sex, none) and adjusted estimates separately. |
| Justification: | Numerous studies didn't provide unadjusted estimates but estimates adjusted for age and sex. Some studies provided only estimates adjusted for age and sex as their maximally adjusted estimate. Given the aims of the review, we chose to treat these as 'minimally adjusted' results and these were pooled alongside crude estimates. |
| Impact: | **Minor**; including the estimates adjusted for age and/or sex in the unadjusted meta-analyses may have slightly attenuated the difference between adjusted vs. unadjusted meta-analyses. However, there was still an observed difference and a more in-depth assessment of confounding bias was also conducted using a subgroup analyses informed by the confounder matrix judgements. |

**Deviation (5):** Data synthesis – confounder matrix

| | |
|---|---|
| Description: | A confounding assessment tool, the 'confounder matrix'[5] that was not pre-specified in the protocol was used to inform the assessment of control for confounding across the studies included in the primary meta-analyses. |
| Justification: | The tool was not publicly available at the time of writing the pre-registration. |
| Impact: | **Minor**; in order to interpret the E-values, an assessment of important confounders controlled for was necessary and recommended by concept developers.[6] The use of a transparent, visual approach was selected as review team felt it aided interpretability. |

**Deviation (6):** Data synthesis – summary estimate (approximate RR vs. OR)

| | |
|---|---|
| Description: | A change in summary estimate for the meta-analyses was made from the odds ratio (OR) to the risk ratio (RR). |
| Justification: | Given that E-values are assessed on the RR scale, and estimates required transformation if outcome prevalence >15%, we selected the square-root transformation available for OR à RR and HR à RR to increase consistency across the review.[7,8] Alternative methods required information frequently not provided by studies. [8,9] This approach has been employed in other published reviews.[10] For outcomes where prevalence was rare (e.g. psychotic disorder), estimates were pooled together without conversion. This is line with approaches employed in other reviews.[11] |
| Impact: | **Major**; as a result, the pooled estimate reflects an 'approximate' RR. The review team felt this was the optimal approach for interpretability of the review. Risk ratios are also recommended as a preferred summary estimate by Cochrane Handbook, due to increased interpretability of the estimate.[4] |

**Deviation (7):** Data synthesis - subgroup analyses

| | |
|---|---|
| Description: | An additional subgroup analysis using judgements from the confounder matrix was added, instead of the planned meta-regression using 'number of covariates'. |
| Justification: | Number of covariates was planned as a meta-regression to explore impact of control for confounding on the pooled estimate. However, this was felt to be less informative than information on 'important' confounders adjusted for from the confounder matrix assessment. |
| Impact: | **Minor**; a quantitative exploration of bias due to inadequate confounding control was still performed. |

**Deviation (8):** Data synthesis – sensitivity analyses

| | |
|---|---|
| Description: | A planned sensitivity analysis *excluding* studies not rated as 'high quality' was instead performed as a subgroup analysis. A planned sensitivity analysis *excluding* studies not using a validated outcome measure was not performed. |
| Justification: | A small number of studies were rated as high quality; therefore, it was deemed a better use of available information to compare estimates across 'low', 'moderate' and 'high' quality studies. There were few studies using non-validated outcome measures, and therefore a planned subgroup analysis by outcome type was deemed satisfactory explanation of the role of outcome measurement in pooled estimate. |
| Impact: | **Minor**; a quantitative exploration of the possible influence of study quality and outcome measure on pooled estimate was still performed. |

**Deviation (9):** Risk of bias in individual studies – additional modifications

| | |
|---|---|
| Description: | Change from total score cut-off to distinguish quality of studies (i.e. low, moderate, high) to using a weighted approach to key items on the Newcastle Ottawa Scale. For details see **eMethods 5**. |
| Justification: | Total score approach has been criticised for not providing a nuanced assessment of possible sources of bias (e.g. a study could score >=7 with limited adjustment for confounding and substantial, or unclear, loss to follow-up). A weighted approach allows key sources of bias (i.e. confounding, attrition) to be preferentially considered in overall judgement of quality, and has been employed in other systematic reviews which assess risk factors for incident mental health conditions.[12] |
| Impact: | **Moderate**; some studies that would have been rated as 'high' based on original cut-off (i.e. >=7) were instead rated as 'moderate' quality due to lack of information about attrition. However, we have also presented total scores in order to enable readers to have access to the overall 'score'. |

**Deviation (10):** E-values for individual studies versus overall synthesis

| | |
|---|---|
| Description: | We planned to calculate E-values for individual studies, and for the overall meta-analyses using the extension of E-values to the random-effects meta-analysis but did not perform the latter. |
| Justification: | Due to feasibility, it was decided that the E-values performed for individual studies in combination with the confounder matrix assessment was sufficient exploration of confounding bias. |
| Impact: | **Minor**; an assessment of confounding bias using E-values was still performed and impact considered on the overall synthesis. Other measures of uncertainty (e.g. 95CIs) were also considered in interpretation of results. |

**eMethods 3**. Search strategy supplemental information

The search strategy for all databases have been provided below. The search terms and strategy were developed by the lead author (CB), reviewed and agreed amongst the review team and reviewed by a scientific librarian. Searches were initially performed in March 2021, and updated again in January 2022 and November 2022. Final searches were run on 18th November 2022.

**Databases:**

CINAHL (via EBSCOhost), Embase (via embase.com), MEDLINE (via PubMed), PsycINFO (via APA PsycNET). Searched from inception to present (i.e. point at which search performed). Base strategy was created in PubMed, which was adapted as required for the syntax and subject headings of the other databases used in the search. The following vocabulary databases were used to find equivalent terms: APA Thesaurus, CINAHL Headings and Emtree. For studies published in languages other than English, Google Translate was used to adequately translate the title, abstract and full text as required.

**Grey literature:**

Unpublished dissertations/theses were searched via the database ProQuest Dissertation & Theses.

**Other sources:**

- The shiny app '*citationchaser*' (https://estech.shinyapps.io/citationchaser/)[13] was used to perform forward and backward citation searching of: (1) included studies, and (2) identified relevant reviews.
- Authors of published conference abstracts without accompanying full-texts were contacted where possible to ascertain study eligibility and acquire relevant results.
- A defined list of 10 experts in the field were successfully contacted via e-mail for unpublished findings or 'file drawer' data that might fulfil inclusion criteria. The response rate was 100%. A list of contacted persons is available on request.

**Search field:**

Text-words searched in Title/Abstract throughout all databases, except where otherwise specified.

**Limits:**

None applied (e.g. language, publication status) – not exclusion criteria for the review.

**De-duplication:**

Applied in Endnote and Covidence.

**Full-text screening:**

Studies were excluded following a hierarchy, as presented in **eTable 1**. As such, studies excluded on a higher-level criteria (e.g. not longitudinal, not binary) do not necessarily meet remaining criteria. Please note, exclusion on 'not longitudinal' refers to both cross-sectional studies, in addition to longitudinal studies with no eligible prospective association (i.e. temporal separation between exposure and outcome could not be established due to data collection methods or analysis).

**Missing data and non-retrievable full-texts:**

All authors of studies excluded on 'FT not retrieved' or 'Missing data' were contacted where a corresponding e-mail address could be located. Reasons for non-retrieval include (1) author non-response, (2) no e-mail address located, and (3) authors no longer having access to the data.

PubMed Search Strategy

All MeSH Terms exploded unless otherwise specified by ‘MeSH Terms:noexp’

| ID | Searches |
|---|---|
| #1 | "cannabis"[MeSH Terms] OR "marijuana abuse"[MeSH Terms] OR "marijuana smoking"[MeSH Terms] OR cannabis[Title/Abstract] OR marijuana[Title/Abstract] OR marihuana[Title/Abstract] |
| #2 | "tobacco use"[MeSH Terms] OR "tobacco products"[MeSH Terms] OR "tobacco smoking"[MeSH Terms] OR "tobacco use disorder"[MeSH Terms] OR cigar*[Title/Abstract] OR smok*[Title/Abstract] OR nicotin*[Title/Abstract] OR tobacco[Title/Abstract] |
| #3 | #1 OR #2 |
| #4 | "schizophrenia spectrum and other psychotic disorders"[MeSH Terms] OR schizo*[Title/Abstract] OR psychosis[Title/Abstract] OR psychotic[Title/Abstract] |
| #5 | "mood disorders"[MeSH Terms] OR "bipolar and related disorders"[MeSH Terms] OR “depression”[MeSH Terms] OR depress*[Title/Abstract] OR bipolar[Title/Abstract] OR dysthymia[Title/Abstract] OR "mood disorder*"[Title/Abstract] |
| #6 | "anxiety disorders"[MeSH Terms] OR "Anxiety"[MeSH Terms:noexp] OR phobia*[Title/Abstract] OR panic[Title/Abstract] OR anxi*[Title/Abstract] |
| #7 | "Mental Disorders"[MeSH Terms:noexp] OR "mental illness*"[Title/Abstract] OR "mental disorder*"[Title/Abstract] OR "psychiatric diagnos*"[Title/Abstract] OR "psychiatric disorder*"[Title/Abstract] |
| #8 | #4 OR #5 OR #6 OR #7 |
| #9 | "Epidemiologic Studies"[MeSH Terms:noexp] OR "cohort studies"[MeSH Terms] OR "case control studies"[MeSH Terms] OR "case control stud*"[Title/Abstract] OR "cohort stud*"[Title/Abstract] OR retrospective[Title/Abstract] OR prospective[Title/Abstract] OR longitudinal[Title/Abstract] OR inciden*[Title/Abstract] OR "follow up"[Title] |
| #10 | #3 AND #8 AND #9 |

Embase

Limits applied: MEDLINE limiter

| ID | Searches |
|---|---|
| #1 | 'cannabis'/exp/mj OR 'cannabis use'/exp/mj OR 'cannabis addiction'/exp/mj OR cannabis:ab,ti OR marijuana:ab,ti OR marihuana:ab,ti |
| #2 | 'tobacco'/exp/mj OR 'tobacco use'/exp/mj OR 'tobacco dependence'/exp/mj OR cigar*:ab,ti OR smok*:ab,ti OR nicotin*:ab,ti OR tobacco:ab,ti |
| #3 | 'psychosis'/exp/mj OR schizo*:ab,ti OR psychotic:ab,ti OR psychosis:ab,ti |
| #4 | 'mood disorder'/exp/mj OR 'bipolar disorder'/exp/mj OR depress*:ab,ti OR 'mood disorder*':ab,ti OR bipolar:ab,ti OR dysthymia:ab,ti |
| #5 | 'anxiety disorder'/exp/mj OR phobia*:ab,ti OR panic:ab,ti OR anxi*:ab,ti |
| #6 | 'mental disease'/mj OR 'mental illness*':ab,ti OR 'mental disorder*':ab,ti OR 'psychiatric disorder*':ab,ti OR 'psychiatric diagnos*':ab,ti |
| #7 | 'epidemiology'/mj OR 'cohort analysis'/mj OR 'case control study'/exp/mj OR 'cohort stud*':ab,ti OR 'case control stud*':ab,ti OR retrospective:ab,ti OR prospective:ab,ti OR longitudinal:ab,ti OR inciden*:ab,ti OR 'follow up':ti |
| #8 | #1 OR #2 |
| #9 | #3 OR #4 OR #5 OR #6 |

| | |
|---|---|
| #10 | #7 AND #8 AND #9 |
| #11 | #7 AND #8 AND #9 NOT [medline]/lim |

<u>CINAHL Search Strategy</u>

| ID | Searches |
|---|---|
| S1 | MH ( (MH "Cannabis+") ) OR TI ( cannabis OR marijuana OR marihuana ) OR AB ( cannabis OR marijuana OR marihuana) |
| S2 | MH ( ( MH "Tobacco") OR (MH "Smoking") OR (MH "Tobacco Products") ) OR TI ( cigar* OR smok* OR nicotin* OR tobacco ) OR AB ( cigar* OR smok* OR nicotin* OR tobacco ) |
| S3 | MH ( (MH "Bipolar Disorder+") OR (MH "Affective Disorders+") ) OR TI ( "mood disorder*" OR depress* OR bipolar OR dysthymia ) OR AB ( "mood disorder*" OR depress* OR bipolar OR dysthymia ) |
| S4 | MH (MH "Anxiety Disorders+") OR TI ( phobia* OR panic OR anxi* ) OR AB ( phobia* OR panic OR anxi* ) |
| S5 | MH (MH "Psychotic Disorders+") OR TI ( psychosis OR schizo* OR psychotic) OR AB ( psychosis OR schizo* OR psychotic) |
| S6 | MH (MH "Mental Disorders") OR TI ( "mental illness*" OR "mental disorder*" OR "psychiatric disorder*" OR "psychiatric diagnos*” ) OR AB ( "mental illness*" OR "mental disorder*" OR "psychiatric disorder*" OR "psychiatric diagnos*” ) |
| S7 | MH ( (MH "Case Control Studies+") OR (MH "Prospective Studies") ) OR TI ( “cohort stud*” OR “case control stud*” OR retrospective OR prospective OR longitudinal OR inciden* OR “follow up” ) OR AB ( “cohort stud*” OR “case control stud*” OR retrospective OR prospective OR longitudinal OR inciden* ) |
| S8 | S1 OR S2 |
| S9 | OR/S3 – S6 |
| S10 | S7 AND S8 AND S9 |

(NB: ‘+’ indicates that subject heading exploded in search)

<u>PsycINFO</u>

| ID | Searches |
|---|---|
| #1 | Index Terms: {Cannabis} OR {Hashish} OR {Marijuana} OR {Cannabis Use Disorder} OR Title: cannabis OR Title: marijuana OR Title: marihuana OR Abstract: cannabis OR Abstract: marijuana OR Abstract: marihuana |
| #2 | Index Terms: {Tobacco Smoking} OR {Smokeless Tobacco} OR {Tobacco Use Disorder} OR Title: cigar* OR Title: smok* OR Title: nicotin* OR Title: tobacco OR Abstract: cigar* OR Abstract: smok* OR Abstract: nicotin* OR Abstract: tobacco |
| #3 | Index Terms: {Psychosis} OR {Schizophrenia} OR {Acute Psychosis} OR {Affective Psychosis} OR {Serious Mental Illness} OR Title: psychosis OR Title: schizo* OR Title: psychotic OR Abstract: psychosis OR Abstract: schizo* OR Abstract: psychotic |
| #4 | Index Terms: {Seasonal Affective Disorder} OR {Major Depression} OR {Disruptive Mood Dysregulation Disorder} OR {Affective Disorders} OR {Mania} OR {Cyclothymic Disorder} OR {Dysthymic Disorder} OR {Bipolar II Disorder} OR {Bipolar I Disorder} OR {Bipolar Disorder} OR Title: “mood disorder*” OR depress* OR bipolar OR dysthymia OR Abstract: “mood disorder*” OR depress* OR bipolar OR dysthymia |
| #5 | Index Terms: {Anxiety Disorders} OR {Generalized Anxiety Disorder} OR {Panic |

| | |
|---|---|
| | Attack} OR {Panic Disorder} OR {Phobias} OR Title: phobia* OR Title: panic OR Title: anxi* OR Abstract: phobia* OR Abstract: panic OR Abstract: anxi* |
| #6 | Index Terms: {Mental Disorders} OR Title: "mental illness*" OR "mental disorder*" OR "psychiatric disorder*" OR "psychiatric diagnos*" OR Abstract: "mental illness*" OR "mental disorder*" OR "psychiatric disorder*" OR "psychiatric diagnos*" |
| #7 | Index Terms: {Epidemiology} OR {Longitudinal Studies} OR {Prospective Studies} OR {Followup Studies} OR {Retrospective Studies} OR Title: “cohort stud*” OR “case control stud*” OR retrospective OR prospective OR longitudinal OR inciden* OR "follow up" OR Abstract: “cohort stud*” OR “case control stud*” OR retrospective OR prospective OR longitudinal OR inciden* |
| #8 | #1 OR #2 |
| #9 | OR/#3-6 |
| #10 | #7 AND #8 AND #9 |

<u>ProQuest Dissertation + Theses</u>

| | |
|---|---|
| S1 | diskw.Exact("Cannabis") OR ti(cannabis OR marijuana OR marihuana) OR ab(cannabis OR marijuana OR marihuana) |
| S2 | diskw.Exact("Tobacco") OR ti(cigar* OR smok* OR nicotin* OR tobacco) OR ab(cigar* OR smok* OR nicotin* OR tobacco) |
| S3 | diskw.Exact("Psychotic") OR ti(psychosis OR schizophren* OR schizoaff* OR schizoty* OR psychotic) OR ab(psychosis OR schizophren* OR schizoaff* OR schizoty* OR psychotic) |
| S4 | diskw.Exact("Depression" OR "Bipolar disorder") OR ti(“mood disorder*” OR depress* OR bipolar OR dysthymia) OR ab(“mood disorder*” OR depress* OR bipolar OR dysthymia) |
| S5 | diskw.Exact("Anxiety disorders") OR ti(anxi* OR phobia* OR panic) OR ab(anxi* OR phobia* OR panic OR) |
| S6 | diskw.Exact("Mental illness") OR ti(“mental illness*” OR “mental disorder*” OR “psychiatric disorder*” OR “psychiatric diagnos*”) OR ab (“mental illness*” OR “mental disorder*” OR “psychiatric disorder*” OR “psychiatric diagnos*”) |
| S7 | diskw.Exact("Epidemiologic Studies" OR "Cohort study" OR "Case-control study") OR ti("cohort studies" OR "cohort study" OR "case control studies" OR "case control study" OR retrospective OR prospective OR longitudinal OR inciden* OR "follow up") OR ab("cohort studies" OR "cohort study" OR "case control studies" OR "case control study" OR retrospective OR prospective OR longitudinal OR inciden*) |
| S8 | S1 OR S2 |
| S9 | OR/S3-S6 |
| S10 | S7 AND S8 AND S9 |

**eMethods 4.** PECOS inclusion/exclusion criteria

| | |
|---|---|
| **Population** | • Broadly representative of general population<br>• Can be selected on basic demographics (e.g. age, sex, ethnicity, occupation), but not eligible if comprised of persons with particular health characteristics (e.g. chronic condition) or other clinically relevant factors (e.g. pregnancy, incarcerated adults, 'high-risk') |
| **Exposure** | • Any measure of tobacco product use (e.g. current smoking, ever smoking, cigarettes per day); excluding products not containing tobacco leaf (e.g. e-cigarettes)<br>• Any measure of recreational cannabis use (e.g. current use, ever use, lifetime frequency)<br>• Any measure of cannabis-tobacco co-use (e.g. concurrent use, co-administration) |
| **Comparator** | • Eligible comparators are: (1) 'non-exposed' (e.g. never use, non-use, past year non-use, non-regular use); (2) other eligible exposures (e.g. *exp* = co-use, *ref* = tobacco use)<br>• Comparators of only lower frequency use groups (e.g. >=20CPD vs >=10CPD) or absence of problematic use (e.g. CUD vs no CUD) are not eligible |
| **Outcome** | • Can be any mood, anxiety or psychotic disorder except for substance-induced disorders (e.g. cannabis-induced psychotic disorder) and outcomes that *only* measure OCD or PTSD<br>• Can be composite measures (e.g. "any mood disorder") or specific conditions (e.g. depressive symptoms), but not eligible if combined across disorder groups (e.g. "any mental illness")<br>• Must be a binary measure of an *incident* condition (i.e. analysis/study excluded participants with a current/lifetime history of condition)<br>• No limit on outcome measurement type, examples may include: self-rated scales, registry codes, interviews, self-reported diagnoses<br>• Must have raw data necessary for meta-analysis or pre-calculated effect estimate (e.g. OR, RR, HR, IRR) |
| **Study Design** | • Longitudinal design (cohort or nested case-control) and must collect and assess the exposure/outcome across multiple timepoints (i.e. retrospective recall not eligible)<br>• Studies without original data (e.g. systematic reviews, commentaries) are excluded, and case-reports, case-series, interventional studies, qualitative studies, animal studies and in-vitro studies are excluded. |
| **Other** | • No limits on article publication status, year of publication or language. |

**eMethods 5.** Modified Newcastle Ottawa Scale (NOS)

Quality of included studies was evaluated using an adapted version of the Newcastle Ottawa Scale (NOS) for case-control and cohort studies (Wells et al., 2013)[14]. The adapted scale can be found in Appendix A of the online protocol (https://osf.io/5t2pu). The original scare can be found online (https://www.ohri.ca/programs/clinical_epidemiology/oxford.asp). Assessments were performed by two independent reviewers, and any disagreements were discussed between reviewers with a third reviewer consulted if a consensus could not be reached.

The NOS evaluates the quality of individual studies across selection of participants, comparability and outcome assessments. In **eTable 8**, scores on each section have been broken down by assessment area i.e. selection (S1, S2, S3, S4), comparability (C1, C2, C3) and outcome (O1, O2, O3). The NOS assigns a maximum of 9 points using a star-based system. Whilst reviews commonly employ standard cut-offs (e.g. ≥7) to denote 'low' vs 'high' risk of bias, this approach has been criticised for not providing a nuanced assessment of possible sources of bias. For example, a study could score ≥7 with limited adjustment for confounding and substantial, or unclear, loss to follow-up.

As such, in this review we employed a weighted approach in which potential risk of bias was awarded based on satisfactory approaches to key sources of bias. To be rated as 'high quality', a study must score 2 stars on items relating to confounding (5) and 1 star on items relating to attrition (8), and only score <1 star on one other item. To be rated as 'moderate quality' a study must score ≥1 on items relating to either confounding (5) or attrition (8) and may score <1 star on more than one other item. To be rated as 'low quality' a study must score <1 star on items related to confounding (5) and attrition (8). This is similar to approaches taken in other systematic reviews which assess risk factors for incident mental health conditions (e.g. Richardson et al., 2015)[12]. Subgroup analyses were performed based on study quality. The 'total score' and number of points allocated per domain (i.e. selection, comparability, exposure/outcome) have also been reported.

**eMethods 6.** Directed Acyclic Graph (DAG)

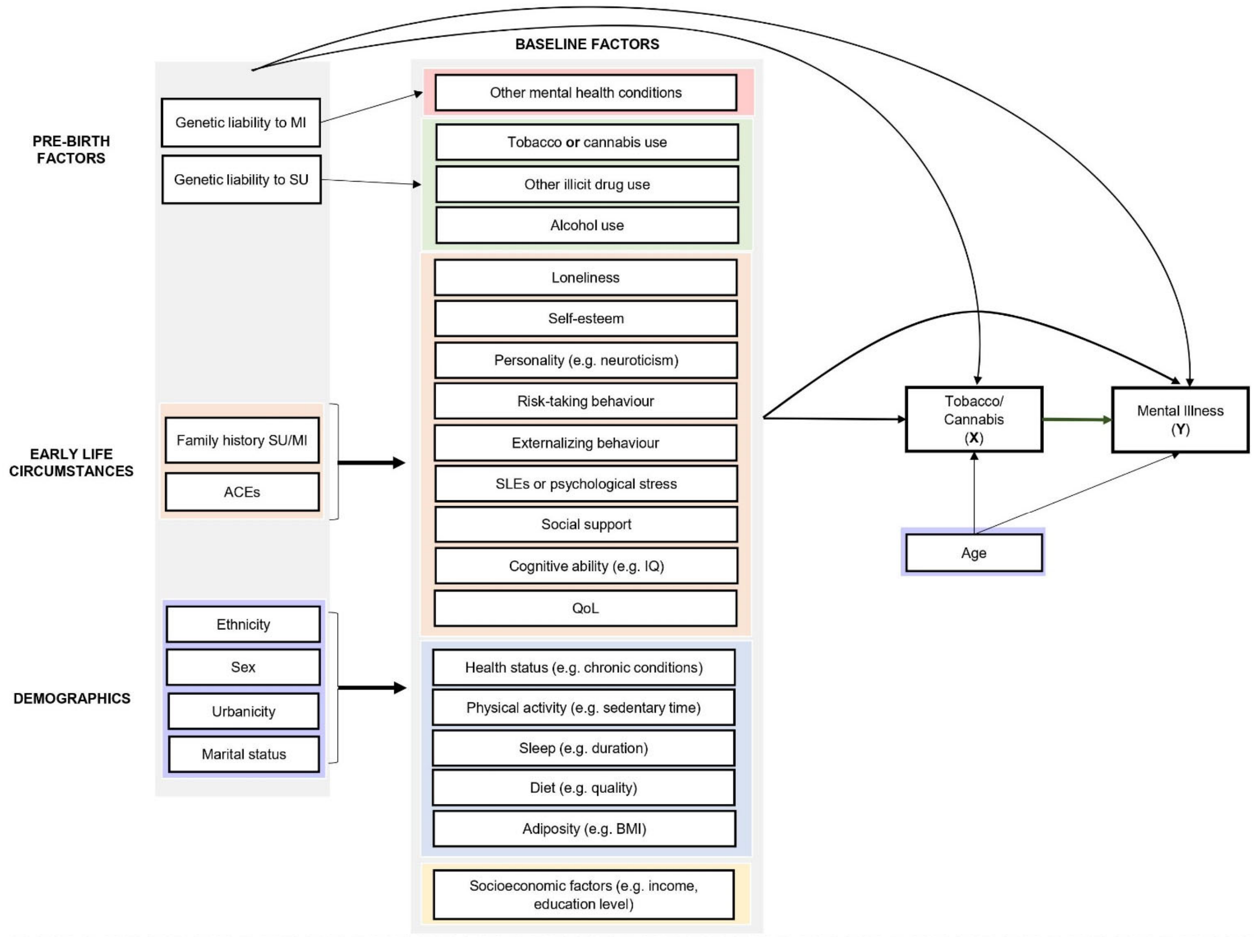

Exposures = X; Outcomes = Y; Green arrow = focal relationship; Bolded arrows represent direction of effect for group contained within bracket. Inward and outward arrows from grey shaded areas pertain to all boxes with the shaded area.
Abbreviations: SES = socioeconomic status; SU = substance use; MI = mental illness; ACEs = adverse childhood experiences; SLEs = stressful life events; QoL = quality of life.
Confounding groups: Other substance use (**Green**); Psychiatric comorbidity (**Pink**); Socioeconomic factors (**Yellow**); Psychological factors (**Orange**); Other lifestyle factors (**Blue**); Demographics (**Purple**).

**eMethods 7.** Confounder matrix judgement criteria

| Construct | Variables Controlled | | |
|---|---|---|---|
| | Adequate | Some Concerns | Inadequate |
| **Other substance use** [a] | Alcohol use + tobacco/cannabis use + other illicit drug use | Some but not all of the variables (e.g., alcohol use only) | None |
| **Psychiatric comorbidity** | Composite indicator of any other mental health condition **or** adjustment for other key groups | Adjustment for one other diagnostic group (e.g., anxiety disorders) | None |
| **Socioeconomic position** [b] | SES index or social class or two of: education, income (e.g. personal, household), unemployment, occupation | Only education, income, employment status or occupation | None |
| **Sociodemographic factors** [b] | Age, sex and one of: race/ethnicity, marital/partner status or urbanicity | Some but not all of the variables (e.g. age + urbanicity but not sex) | Only age or sex |
| **Psychological factors** | Two of: stressful life events, ACEs, loneliness, personality, self-esteem, social support, risk-taking externalizing behaviours, family history of mental illness, IQ/cognitive ability, quality of life | Adjustment for only one psychological factor (e.g., ACEs) | None |
| **Lifestyle factors** | Two of: physical activity, diet, adiposity, sleep, health status | Adjustment for only one lifestyle factor (e.g. diet) | None |

[a] For middle/older adult populations, adequate adjustment was alcohol-use only given the low prevalence of cannabis use and other controlled substances (e.g. cocaine, heroin) in these groups.

[b] For adolescent populations, variables measuring family-level factors (e.g. family structure, parental income, parental education) were considered for assessment.

**eMethods 8.** Statistical approach

Included studies presented varied effect estimates. For studies providing only raw data, an unadjusted risk ratio (RR) was calculated using the number of positive and negative events in the exposed group and reference group (see **Formula 1**). The same approach was applied when studies did not present crude effect estimates, to facilitate inclusion in both adjusted and unadjusted meta-analyses. For studies which fulfilled the 'rare event' assumption (i.e. outcome prevalence <15%) the odds ratio (OR) and Hazard Ratio (HR) were considered to approximate the RR.[7,8] Each study was checked for reported outcome prevalence, rather than assuming rare events. Where the outcome prevalence was >15%, the OR and HR were approximately transformed using the square-root transformation.[8] See **Formula 2** (OR → RR) and **Formula 3** (HR → RR).[8] The Incident Rate Ratio (IRR) was approximated as an HR, and treated as described (i.e. >15% converted to approximate RR using square-root transformation).

Studies which only presented effect measures stratified by population characteristics (e.g. age, sex, ethnicity) were combined using random-effects meta-analysis and included as one estimate. For studies which presented frequency of use estimates (e.g. 1-9 CPD, 19-20 CPD, ≥20CPD), only the estimate for the highest use category was used in the main meta-analysis unless there were justified reasons to extract an alternative estimate (e.g. alternative exposure level subject to multiple imputation or buffer period to reduce risk of prodromal symptoms). Adjusted and unadjusted effect estimates were extracted and pooled separately. Studies which provided estimates that were adjusted only for age and/or sex (i.e. restriction, stratification, statistical) were not included in the primary meta-analysis as they were deemed insufficiently adjusted. These were instead included in the unadjusted meta-analysis as 'minimally adjusted' estimates.

Outliers were excluded as a exploratory sensitivity analysis, defined as all studies for which **upper bound of the 95% confidence interval of the study is lower than the lower bound of the pooled effect confidence interval** (i.e., extremely small effects) **and for which the lower bound of the 95% confidence interval of the study is higher than the higher bound of the pooled effect confidence interval** (i.e., extremely large effects).

**Formula 1:**

$$RR_i = \frac{a_i \,/\, n_{1i}}{c_i \,/\, n_{2i}}$$

**Formula 2:**

$$\{\frac{\mathrm{OR}(u + \mathrm{OR} - \mathrm{ORu})}{1 - w + \mathrm{ORw}}\}^{1/2}$$

**Formula 3:**

$$\{\frac{1-(1-w)^{\phi}}{1-(1-u)^{1/\phi}}\frac{u}{w}\}^{1/2}$$

**eMethods 9.** References

1. Stroup DF, Berlin JA, Morton SC, et al. Meta-analysis of Observational Studies in EpidemiologyA Proposal for Reporting. *JAMA*. 2000;283(15):2008-2012. doi:10.1001/jama.283.15.2008

2. Moreau D, Gamble B. Conducting a meta-analysis in the age of open science: Tools, tips, and practical recommendations. *Psychological Methods*. 2022;27:426-432. doi:10.1037/met0000351

3. Fluharty M, Taylor AE, Grabski M, Munafò MR. The Association of Cigarette Smoking With Depression and Anxiety: A Systematic Review. *Nicotine Tob Res*. 2017;19(1):3-13. doi:10.1093/ntr/ntw140

4. Higgins J, Thomas J, Chandler J, et al. Cochrane Handbook for Systematic Reviews of Interventions. Published 2020. Accessed February 12, 2021. www.training.cochrane.org/handbook

5. Petersen JM, Barrett M, Ahrens KA, et al. The confounder matrix: A tool to assess confounding bias in systematic reviews of observational studies of etiology. *Research Synthesis Methods*. 2022;13(2):242-254. doi:10.1002/jrsm.1544

6. VanderWeele TJ, Ding P. Sensitivity Analysis in Observational Research: Introducing the E-Value. *Ann Intern Med*. 2017;167(4):268-274. doi:10.7326/M16-2607

7. VanderWeele T. On a square-root transformation of the odds ratio for a common outcome. *Epidemiology*. 2017;28(6):e58-e60. doi:10.1097/EDE.0000000000000733

8. VanderWeele TJ. Optimal approximate conversions of odds ratios and hazard ratios to risk ratios. *Biometrics*. 2020;76(3):746-752. doi:10.1111/biom.13197

9. Zhang J, Yu KF. What's the Relative Risk?A Method of Correcting the Odds Ratio in Cohort Studies of Common Outcomes. *JAMA*. 1998;280(19):1690-1691. doi:10.1001/jama.280.19.1690

10. Ahn N, Nolde M, Krause E, et al. Do proton pump inhibitors increase the risk of dementia? A systematic review, meta-analysis and bias analysis. *British Journal of Clinical Pharmacology*. 2023;89(2):602-616. doi:10.1111/bcp.15583

11. Hunter A, Murray R, Asher L, Leonardi-Bee J. The Effects of Tobacco Smoking, and Prenatal Tobacco Smoke Exposure, on Risk of Schizophrenia: A Systematic Review and Meta-Analysis. *Nicotine & Tobacco Research*. 2020;22(1):3-10. doi:10.1093/ntr/nty160

12. Richardson R, Westley T, Gariépy G, Austin N, Nandi A. Neighborhood socioeconomic conditions and depression: a systematic review and meta-analysis. *Soc Psychiatry Psychiatr Epidemiol*. 2015;50(11):1641-1656. doi:10.1007/s00127-015-1092-4

13. Haddaway, N R, Grainger, M. J., Gray, C. T. citationchaser: An R package and Shiny app for forward and backward citations chasing in academic searching. Published online February 16, 2021. doi:10.5281/ZENODO.4543513

14. Wells G, Shea B, O'Connell D, et al. The Nwecastle-Ottawa Scale (NOS) for assessing the quality of nonrandomised studies in meta-analyses. Published 2013. Accessed February 12, 2021. http://www.ohri.ca/programs/clinical_epidemiology/oxford.asp

**eTable 1.** Studies excluded at full-text screening

| First Author/Year | DOI/URL/PMID | Exclusion reason |
|---|---|---|
| Barata 2018 | 10.1183/13993003.congress-2018.PA1256 | FT not retrieved |
| Bourque 2016 | 10.1016/j.jaac.2016.09.406 | FT not retrieved |
| Cao 2014 | 10.1111/jgs.13075 | FT not retrieved |
| Cheng 2014 | N/A | FT not retrieved |
| Delgado 1989 | https://www.researchgate.net/publication/291745248_Cannabis_consumption_Incidence_and_psychological_effects | FT not retrieved |
| Duroseau 2020 | 10.1016/j.jadohealth.2019.11.259 | FT not retrieved |
| Marfatia 2009 | 10.1002/pds.1806 | FT not retrieved |
| Martinez-Hernaez 2015 | 10.1016/S0924-9338(15)30153-X | FT not retrieved |
| Michal 2015 | N/A | FT not retrieved |
| Prom-Wormley 2012 | 10.1007/s10519-012-9566-6 | FT not retrieved |
| Regeer 2009 | 10.1016/S0924-9338(09)70293-7 | FT not retrieved |
| Stuchlik 2015 | 10.1161/circ.132.suppl_3.18557 | FT not retrieved |
| Bakhshaie 2014 | 10.1016/j.comppsych.2014.10.012 | Duplicate |
| Fritz 2016 | https://scholar.colorado.edu/concern/undergraduate_honors_theses/8910jv21p | Duplicate |
| Hill 2017 | 10.1001/jamapsychiatry.2015.3229 | Duplicate |
| Addington 2005 | 10.1136/ebmh.8.3.87. | Ineligible design |
| Bulacia 2016 | 28898309 | Ineligible design |
| Beutel 2020 | 10.13109/zptm.2020.66.4.355 | Ineligible design |
| Costello 2016 | 10.1007/s00127-015-1168-1 | Ineligible design |
| Fergusson 2015 | 10.1007/s00127-015-1070-x | Ineligible design |
| Kirkbride 2011 | 10.1136/ebmh.14.3.70 | Ineligible design |
| Kuepper 2011 | 10.1016/j.schres.2011.06.012 | Ineligible design |
| Lewinsohn 1998 | 10.1016/s0272-7358(98)00010-5 | Ineligible design |
| Nishida 2007 | 17561673 | Ineligible design |
| Noda 2016 | 10.11560/jahp.28.special_issue_129 | Ineligible design |
| SeguíDíaz 2017 | 10.1016/j.semerg.2016.05.005 | Ineligible design |
| Skosnik 2007 | 10.1136/ebmh.10.2.61 | Ineligible design |
| Abuladze 2020 | 10.1177/2050312120974167 | Not longitudinal |
| Agrawal 2007 | 10.1111/j.1360-0443.2006.01630.x | Not longitudinal |
| Agrawal 2017 | 10.1016/s2215-0366(17)30280-8 | Not longitudinal |
| Baggio 2014 | 10.1515/ijamh-2013-0305 | Not longitudinal |
| Bandiera 2015 | 10.1093/ntr/ntu209 | Not longitudinal |
| Barkhuizen 2020 | 10.1016/j.jaac.2018.06.037 | Not longitudinal |
| Barrera 2016 | 28898304 | Not longitudinal |
| Baskak 2012 | 10.1016/j.psychres.2012.01.027 | Not longitudinal |
| Bhandari 2020 | 10.1007/s12126-020-09371-0 | Not longitudinal |
| Buckner 2012 | 10.1016/j.drugalcdep.2011.12.023 | Not longitudinal |
| Campbell 2020 | 10.1080/02791072.2020.1747665 | Not longitudinal |
| Carceller-Maicas 2014 | 10.20882/adicciones.127 | Not longitudinal |
| Catalao 2022 | 10.1136/bmjopen-2021-059257 | Not longitudinal |
| Chabrol 2004 | 10.1016/S0013-7006(04)95424-3 | Not longitudinal |
| Chaiton 2015 | 10.1016/j.addbeh.2014.11.026 | Not longitudinal |

| | | |
|---|---|---|
| Chen 2002 | 10.1007/s00127-002-0541-z | Not longitudinal |
| Chéron-Launay 2011 | 10.1016/j.addbeh.2011.02.013 | Not longitudinal |
| Cheung 2010 | 10.3109/00952991003713784 | Not longitudinal |
| Cougle 2010 | 10.1093/ntr/ntq006 | Not longitudinal |
| Cruz-Barreda 2022 | 10.1016/j.ypmed.2022.107156 | Not longitudinal |
| Cunningham 2022 | 10.1016/j.amepre.2021.06.024 | Not longitudinal |
| Davis 2019 | 10.1016/j.drugalcdep.2019.107696 | Not longitudinal |
| Degenhardt 2001 | 10.1080/09652140120080732 | Not longitudinal |
| Degenhardt 2003 | 10.1016/s0376-8716(03)00064-4 | Not longitudinal |
| DeSousa 2013 | 10.1016/j.schres.2013.10.037 | Not longitudinal |
| Dominguez 2010 | 10.1176/appi.ajp.2010.09060883 | Not longitudinal |
| Emerson 2018 | 10.1016/j.mhp.2018.09.002 | Not longitudinal |
| Espada 2011 | https://www.redalyc.org/pdf/560/56017110002.pdf | Not longitudinal |
| Ferdinand 2005 | 10.1111/j.1360-0443.2005.01070.x | Not longitudinal |
| Ferdinand 2005 | 10.1016/j.schres.2005.07.027 | Not longitudinal |
| Fernandez-Pujals 2015 | 10.1371/journal.pone.0142197 | Not longitudinal |
| Field 2001 | 11817630 | Not longitudinal |
| Geus 2004 | https://www.researchgate.net/publication/289162839_Psychotic_disorders_and_use_of_cannabis_Retrospective_study | Not longitudinal |
| Gonçalves-Pinho 2019 | 10.1002/mpr.1813 | Not longitudinal |
| Goodwin 2012 | 10.1111/j.1521-0391.2012.00263.x | Not longitudinal |
| Green 2000 | 10.2307/2676359 | Not longitudinal |
| Haavisto 2004 | 10.1016/j.jad.2004.06.008 | Not longitudinal |
| Han 2019 | 10.1016/j.jad.2019.07.003 | Not longitudinal |
| Harder 2008 | 10.1093/aje/kwn184 | Not longitudinal |
| Hearld 2015 | 10.1016/j.jad.2014.11.041 | Not longitudinal |
| Hong 2020 | 10.1007/s00127-020-01945-2 | Not longitudinal |
| Hooker 2022 | 10.1016/j.jpsychores.2022.110920 | Not longitudinal |
| Jitnarin 2015 | 10.1093/ntr/ntu131 | Not longitudinal |
| Kawasaki 2015 | 10.1539/sangyoeisei.b14011 | Not longitudinal |
| Keskitalo-Vuokko 2016 | 10.1017/thg.2016.36 | Not longitudinal |
| Kim 2021 | 10.3390/ijerph18189887 | Not longitudinal |
| Kim 2022 | 10.1186/s12877-021-02729-2 | Not longitudinal |
| Kiviruusu 2022 | 10.1080/08039488.2021.2019912 | Not longitudinal |
| Liu 2022 | 10.1080/08039488.2021.2019912 | Not longitudinal |
| Livne 2021 | 10.1176/appi.ajp.2021.21010073 | Not longitudinal |
| Lyons 2008 | 10.1080/14622200701705332 | Not longitudinal |
| Martínez-Ortega 2013 | 10.1016/j.jpsychires.2013.03.012 | Not longitudinal |
| Martín-Merino 2010 | 10.4088/PCC.08m00764blu | Not longitudinal |
| Martín-Merino 2010 | 10.1093/fampra/cmp071 | Not longitudinal |
| Massak 2008 | 10.1080/14622200802163449 | Not longitudinal |
| McCaffery 2008 | 10.1037/0278-6133.27.3(suppl.).s207 | Not longitudinal |
| McGrath 2010 | 10.1001/archgenpsychiatry.2010.6 | Not longitudinal |
| McGrath 2016 | 10.1177/0004867415587341 | Not longitudinal |
| Miettunen 2008 | 10.1192/bjp.bp.107.045740 | Not longitudinal |
| Mykletun 2007 | 10.1016/j.eurpsy.2007.10.005 | Not longitudinal |

| | | |
|---|---|---|
| Nesvåg 2017 | 10.1093/schbul/sbw101 | Not longitudinal |
| Noh 2014 | 10.4306/pi.2014.11.3.272 | Not longitudinal |
| Park 2013 | 10.1097/jan.0b013e3182a4cad3 | Not longitudinal |
| Pedersen 2009 | 10.4045/tidsskr.09.34699 | Not longitudinal |
| Punjani 2018 | 10.1177/1557988318799022 | Not longitudinal |
| Purborini 2021 | 10.1016/j.jfma.2021.01.016 | Not longitudinal |
| Quadros 2020 | 10.1590/0034-7167-2018-0162 | Not longitudinal |
| Rabiee 2020 | 10.1016/j.drugalcdep.2020.108332 | Not longitudinal |
| Radhakrishnan 2019 | 10.1017/s0033291718002635 | Not longitudinal |
| Riala 2005 | http://jultika.oulu.fi/Record/isbn951-42-7495-4 | Not longitudinal |
| Romans 1993 | 10.3109/00048679309075795 | Not longitudinal |
| Rougemont-Bücking 2019 | 10.1080/02791072.2019.1571258 | Not longitudinal |
| Santangelo 2018 | 10.4081/mi.2018.7649 | Not longitudinal |
| Scholes-Balog 2013 | 10.1016/j.adolescence.2013.03.001 | Not longitudinal |
| Shevlin 2017 | 10.1108/dat-03-2017-0014 | Not longitudinal |
| Sideli 2015 | 10.1111/eip.12285 | Not longitudinal |
| Slomp 2019 | 10.2147/NDT.S217069 | Not longitudinal |
| Son 1997 | 10.1093/oxfordjournals.aje.a009081 | Not longitudinal |
| Sourander 2004 | 10.1097/01.chi.0000134493.88549.e2 | Not longitudinal |
| Stefanis 1976 | 10.1111/j.1749-6632.1976.tb49885.x | Not longitudinal |
| Van der Pol 2013 | 10.1111/add.12196 | Not longitudinal |
| Yunis 2003 | 10.2224/sbp.2003.31.5.461 | Not longitudinal |
| Zablocki 1991 | 10.2307/2136800 | Not longitudinal |
| Zvolensky 2008 | 10.1016/j.jpsychires.2006.09.012 | Not longitudinal |
| Andreas 2021 | 10.1177/00048674211025711 | No X/Y association |
| Benjet 2015 | 10.1007/s00787-015-0721-5 | No X/Y association |
| Bierhoff 2019 | 10.1080/10826084.2019.1581220 | No X/Y association |
| Boffin 2012 | 10.1093/fampra/cms024 | No X/Y association |
| Bogren 2009 | https://lup.lub.lu.se/search/publication/1484236 | No X/Y association |
| Bratlien 2014 | 10.1016/j.psychres.2013.12.048 | No X/Y association |
| Brook 2010 | 10.1093/ntr/ntq027 | No X/Y association |
| Cartier 2017 | 10.4158/ep161456.Or | No X/Y association |
| Conde-Sala 2018 | 10.1016/j.jad.2018.10.358 | No X/Y association |
| Crane 2015 | 10.1016/j.addbeh.2015.05.014 | No X/Y association |
| Crespo 2022 | 10.1016/j.ypmed.2021.106932 | No X/Y association |
| deGraaf 2002 | 10.1034/j.1600-0447.2002.01397.x | No X/Y association |
| deGraaf 2013 | 10.1016/j.jad.2013.01.009 | No X/Y association |
| Filatova 2016 | 10.1017/s2045796016000123 | No X/Y association |
| Forsell 1999 | https://pubmed.ncbi.nlm.nih.gov/10389040/ | No X/Y association |
| Grant 2008 | 10.1038/mp.2008.41 | No X/Y association |
| Harris 2005 | 10.1093/ageing/afi216 | No X/Y association |
| Hofstra 2002 | 10.1097/00004583-200202000-00012 | No X/Y association |
| Holley 2006 | 10.1097/01.jgp.0000192504.48810.cb | No X/Y association |
| Kang 2015 | 10.1017/s1041610215001301 | No X/Y association |
| Kim 2006 | 10.1192/bjp.bp.105.015032 | No X/Y association |
| Köhler 2007 | 10.1007/s00127-007-0171-6 | No X/Y association |
| Lyness 2009 | 10.1176/appi.ajp.2009.08101489 | No X/Y association |

| | | |
|---|---|---|
| Mazza 2008 | 10.1177/0272431608324193 | No X/Y association |
| Misawa 2018 | 10.1080/13607863.2018.1496225 | No X/Y association |
| Mojtabai 2004 | 10.1017/s0033291703001764 | No X/Y association |
| Moutinho 2019 | 10.1016/j.psychres.2019.02.041 | No X/Y association |
| Ogasawara 2011 | 10.1016/j.jad.2010.06.015 | No X/Y association |
| Phifer 1986 | 10.1037/0021-843x.95.3.282 | No X/Y association |
| Reinherz 1993 | 10.1097/00004583-199311000-00007 | No X/Y association |
| Shedler 1990 | 10.1037//0003-066x.45.5.612 | No X/Y association |
| Smit 2004 | 10.1016/j.jad.2003.08.007 | No X/Y association |
| Stek 2006 | 10.1192/bjp.188.1.65 | No X/Y association |
| Uemura 2017 | 10.1002/gps.4776 | No X/Y association |
| vanGool 2003 | 10.1093/ageing/32.1.81 | No X/Y association |
| Weinstein 2013 | 10.1037/a0031488 | No X/Y association |
| Welzel 2021 | 10.3390/ijerph182312786 | No X/Y association |
| Whitbeck 2009 | 10.1016/j.addbeh.2008.12.009 | No X/Y association |
| Yu 2016 | 10.1016/j.addbeh.2016.02.007 | No X/Y association |
| Aneshensel 1983 | 10.1037//0021-843x.92.2.134 | Not binary |
| Anglin 2012 | 10.1016/j.schres.2012.01.019 | Not binary |
| Anstey 2009 | 10.1097/PSY.0b013e3181beab60 | Not binary |
| Arria 2016 | 10.1016/j.drugalcdep.2015.12.009 | Not binary |
| Assari 2018 | 10.3389/fpsyg.2018.02135 | Not binary |
| Audrain-McGovern 2009 | 10.1111/j.1360-0443.2009.02617.x | Not binary |
| Baggio 2014 | 10.1111/add.12490 | Not binary |
| Bares 2014 | 10.1080/15504263.2014.961852 | Not binary |
| Beal 2014 | 10.1007/s11121-013-0402-x | Not binary |
| Bilsky 2021 | 10.1016/j.addbeh.2021.106981 | Not binary |
| Block 1991 | 10.1037//0022-3514.60.5.726 | Not binary |
| Bourque 2018 | 10.1001/jamapsychiatry.2018.1330 | Not binary |
| Brook 2006 | 10.2466/pr0.99.2.421-438 | Not binary |
| Brook 2013 | 10.2466/15.13.PR0.113x26z6 | Not binary |
| Caldeira 2012 | 10.1016/j.drugalcdep.2012.02.022 | Not binary |
| Campbell 2020 | 10.1186/s40695-020-00050-3 | Not binary |
| Capaldi 2022 | 10.1177/11782218221096154 | Not binary |
| Chang 2018 | 10.1007/s10802-018-0414-x | Not binary |
| DeBoer 2021 | 10.1017/S0033291721002968 | Not binary |
| Duncan 2021 | 10.1007/s00127-020-01900-1 | Not binary |
| Duperrouzel 2018 | 10.1016/j.addbeh.2017.11.005 | Not binary |
| Fergusson 2015 | 10.1016/s2215-0366(15)00208-4 | Not binary |
| Fleming 2008 | 10.1037/0893-164x.22.2.186 | Not binary |
| Frost 1999 | 10.1037/h0080411 | Not binary |
| Griffith-Lendering 2011 | 10.1016/j.drugalcdep.2010.11.024 | Not binary |
| Griffith-Lendering 2013 | 10.1111/add.12050 | Not binary |
| Grunberg 2015 | 10.1037/adb0000109 | Not binary |
| Gunn 2020 | 10.1037/pha0000357 | Not binary |
| Guttmannova 2017 | 10.1016/j.drugalcdep.2017.06.016 | Not binary |
| Hoffmann 2018 | 10.4236/health.2018.108080 | Not binary |
| Hooshmand 2012 | 10.1016/j.jadohealth.2011.05.016 | Not binary |

| | | |
|---|---|---|
| Horwood 2012 | 10.1016/j.drugalcdep.2012.06.002 | Not binary |
| Hossain 2019 | 10.1016/j.jad.2019.06.001 | Not binary |
| Leadbeater 2019 | 10.1111/add.14459 | Not binary |
| Lee 2021 | https://www.proquest.com/docview/2454357025?pq-origsite=gscholar&fromopenview=true | Not binary |
| Lin 2014 | 10.1016/j.ridd.2014.07.018 | Not binary |
| London-Nadeau 2021 | 10.1037/abn0000542 | Not binary |
| Magee 2020 | 10.1016/j.addbeh.2020.106641 | Not binary |
| Marsden 2019 | 10.1016/j.addbeh.2019.106078 | Not binary |
| McIntyre 2020 | https://www.proquest.com/openview/bb5b57a9f10550e75cd5ff6f0f0172c3/1?pq-origsite=gscholar&cbl=18750&diss=y | Not binary |
| Meier 2020 | 10.1007/s10802-020-00641-8 | Not binary |
| Melamed 2020 | 10.1111/sjop.12699 | Not binary |
| Mino 2001 | 10.1006/pmed.2000.0803 | Not binary |
| Moylan 2013 | 10.1371/journal.pone.0063252 | Not binary |
| Munafò 2008 | 10.1111/j.1360-0443.2007.02052.x | Not binary |
| Nault-Briere 2011 | https://papyrus.bib.umontreal.ca/xmlui/handle/1866/6150 | Not binary |
| Needham 2007 | 10.1016/j.socscimed.2007.04.037 | Not binary |
| Newcomb 1993 | 10.1037/1064-1297.1.1-4.215 | Not binary |
| Newcomb 1999 | 10.1002/(SICI)1520-6629(199907)27:4<405::AID-JCOP4>3.0.CO;2-2 | Not binary |
| Ostrowsky 2006 | https://psycnet.apa.org/record/2007-99015-033 | Not binary |
| Otten 2013 | 10.1111/j.1369-1600.2011.00380.x | Not binary |
| Otten 2017 | 10.1111/adb.12372 | Not binary |
| Pahl 2011 | 10.1017/s0033291710002345 | Not binary |
| Patten 2010 | 10.1177/070674371005501006 | Not binary |
| Pedersen 2009 | 10.1111/j.1360-0443.2008.02395.x | Not binary |
| Piumatti 2018 | 10.1016/j.psychres.2017.12.009 | Not binary |
| Poole 2018 | 10.1007/s12529-017-9703-y | Not binary |
| Ranjit 2019 | 10.1007/s11121-019-01020-6 | Not binary |
| Ranjit 2019 | 10.1016/j.drugalcdep.2019.03.012 | Not binary |
| Repetto 2005 | 10.1037/0278-6133.24.2.209 | Not binary |
| Repetto 2008 | 10.1111/j.1532-7795.2008.00566.x | Not binary |
| Sarris 2020 | 10.1186/s12916-020-01813-5 | Not binary |
| Schaefer 2021 | 10.1037/abn0000701 | Not binary |
| Scholes-Balog 2015 | 10.1177/0272431614540526 | Not binary |
| Scholes-Balog 2016 | 10.1016/j.addbeh.2015.09.008 | Not binary |
| Schuler 2015 | 10.1016/j.drugalcdep.2015.10.005 | Not binary |
| Shanahan 2021 | 10.1016/j.drugalcdep.2021.109063 | Not binary |
| Thompson 2018 | 10.1037/cbs0000090 | Not binary |
| Thompson 2021 | 10.3390/ijerph18073652 | Not binary |
| Tjora 2014 | 10.1111/add.12522 | Not binary |
| Tubman 1990 | 10.1016/s0899-3289(10)80004-5 | Not binary |
| Tucker 2019 | 10.1016/j.drugalcdep.2019.06.004 | Not binary |
| Tully 2010 | 10.1017/s0954579410000490 | Not binary |

| | | |
|---|---|---|
| Vaillant 1990 | 10.1176/ajp.147.1.31 | Not binary |
| vanGastel 2014 | 10.1016/j.schres.2014.04.023 | Not binary |
| Velten 2018 | 10.1186/s12889-018-5526-2 | Not binary |
| Vermeulen 2019 | 10.1016/s2215-0366(18)30424-3 | Not binary |
| Walsh 2013 | 10.1007/s13142-012-0189-5 | Not binary |
| Wang 1996 | 10.2466/pr0.1996.79.1.127 | Not binary |
| Washburn 2014 | 10.1017/s0954579414000686 | Not binary |
| Wilhoit 2013 | https://www.proquest.com/openview/5f9534622a345e41197b365d9beb6daa/1?pq-origsite=gscholar&cbl=18750 | Not binary |
| Wilkinson 2016 | 10.1016/j.jadohealth.2016.07.010 | Not binary |
| Wilkinson 2016 | 10.1016/j.addbeh.2016.03.036 | Not binary |
| Williams 2021 | 10.3390/ijerph181910468 | Not binary |
| Williams 2022 | 10.1097/CXA.0000000000000144 | Not binary |
| Womack 2016 | 10.15288/jsad.2016.77.287 | Not binary |
| Wymbs 2014 | 10.15288/jsad.2014.75.269 | Not binary |
| Xu 2010 | 10.1080/07399332.2010.486096 | Not binary |
| Almeida 2006 | 10.1097/01.Jgp.0000192486.20308.42 | Not incident |
| Ajdacic-Gross 2009 | 10.1111/j.1360-0443.2009.02640.x | Not incident |
| Amialchuk 2022 | 10.1002/hec.4469 | Not incident |
| Anderson 2022 | 10.1177/13591053211072685 | Not incident |
| Andréasson 1987 | 10.1016/s0140-6736(87)92620-1 | Not incident |
| Andréasson 1989 | 10.1111/j.1600-0447.1989.tb10296.x | Not incident |
| Angst 1996 | 10.1192/S0007125000298383 | Not incident |
| Anstey 2007 | 10.1097/JGP.0b013e31802e21d8 | Not incident |
| Arseneault 2002 | 10.1136/bmj.325.7374.1212 | Not incident |
| Bagade 2022 | 10.1038/s41598-022-15064-2 | Not incident |
| Bechtold 2015 | 10.1037/adb0000103 | Not incident |
| Bechtold 2016 | 10.1176/appi.ajp.2016.15070878 | Not incident |
| Boden 2010 | 10.1192/bjp.bp.109.065912 | Not incident |
| Boden 2020 | 10.1111/add.14814 | Not incident |
| Bolanis 2020 | 10.1016/j.jad.2020.05.136 | Not incident |
| Breslau 1999 | 10.1001/archpsyc.56.12.1141 | Not incident |
| Brook 2002 | 10.1001/archpsyc.59.11.1039 | Not incident |
| Brook 2004 | 10.2466/pr0.95.1.159-166 | Not incident |
| Brook 2014 | 10.2105/AJPH.2014.301880 | Not incident |
| Brook 2016 | 10.1016/j.addbeh.2016.06.003 | Not incident |
| Bruin 2018 | 10.1002/gps.4889 | Not incident |
| Cardno 2021 | 10.1176/appi.prcp.20200010 | Not incident |
| Caspi 2005 | 10.1016/j.biopsych.2005.01.026 | Not incident |
| Chan 2021 | 10.1111/dar.13239 | Not incident |
| Chang 2022 | 10.1007/s11469-022-00912-z | Not incident |
| Cho 2018 | 10.1093/ntr/ntx270 | Not incident |
| Copeland 2021 | 10.1016/j.jaac.2021.07.824 | Not incident |
| Costello 1999 | 10.1207/S15374424jccp280302 | Not incident |
| Degenhardt 2010 | 10.1192/bjp.bp.108.056952 | Not incident |
| Degenhardt 2013 | 10.1111/j.1360-0443.2012.04015.x | Not incident |
| Du 2022 | 10.3389/fpubh.2022.913636 | Not incident |

| | | |
|---|---|---|
| Dugan 2015 | 10.1249/MSS.0000000000000407 | Not incident |
| Duncan 2005 | 10.1093/aje/kwi219 | Not incident |
| Feng 2018 | 10.1016/S0140-6736(18)32688-6 | Not incident |
| Fergusson 1996 | 10.1007/bf01441571 | Not incident |
| Fergusson 1997 | 10.1111/j.1360-0443.1997.tb03198.x | Not incident |
| Fergusson 2002 | 10.1046/j.1360-0443.2002.00103.x | Not incident |
| Fergusson 2003 | 10.1017/s0033291703008596 | Not incident |
| Fergusson 2003 | 10.1017/s0033291702006402 | Not incident |
| Fergusson 2005 | 10.1111/j.1360-0443.2005.01001.x | Not incident |
| Fergusson 2011 | 10.1007/s00127-010-0268-1 | Not incident |
| Franco 2019 | 10.1016/j.drugalcdep.2019.03.003 | Not incident |
| Frederick 1988 | 10.1016/0091-7435(88)90061-8 | Not incident |
| Georgiades 2007 | 10.1111/j.1469-7610.2007.01740.x | Not incident |
| Green 2012 | 10.1016/j.addbeh.2012.06.008 | Not incident |
| Green 2017 | 10.1080/00952990.2016.1258706 | Not incident |
| Gueltzow 2021 | 10.4054/mpidr-wp-2021-017 | Not incident |
| Guo 2021 | 10.1186/s12911-021-01674-9 | Not incident |
| Hamdi 2016 | https://conservancy.umn.edu/handle/11299/201036 | Not incident |
| Hamer 2008 | 10.1001/archinte.168.22.2474 | Not incident |
| Handley 2019 | 10.1007/s00127-018-1591-1 | Not incident |
| Harder 2006 | 10.1111/j.1360-0443.2006.01545.x | Not incident |
| Hashmi 2022 | 10.1371/journal.pone.0267191 | Not incident |
| Hayatbakhsh 2007 | 10.1097/chi.0b013e31802dc54d | Not incident |
| Hengartner 2020 | 10.1016/j.jad.2020.03.126 | Not incident |
| Henquet 2005 | 10.1136/bmj.38267.664086.63 | Not incident |
| Hozawa 2006 | 10.1016/j.ypmed.2005.12.008 | Not incident |
| Jackson 2004 | https://digitalcommons.library.tmc.edu/dissertations/AAI3155309/ | Not incident |
| John 2004 | 10.1016/j.drugalcdep.2004.06.004 | Not incident |
| Johnson 2000 | 10.1001/jama.284.18.2348 | Not incident |
| Johnson 2004 | 10.1080/14622200412331324901 | Not incident |
| Johnson 2006 | 10.1080/14622200600576644 | Not incident |
| Jones 2018 | 10.1001/jamapsychiatry.2017.4271 | Not incident |
| Kendler 1993 | 10.1001/archpsyc.1993.01820130038007 | Not incident |
| Keyes 2011 | 10.1016/j.socscimed.2010.12.005 | Not incident |
| Klungsøyr 2006 | 10.1093/aje/kwj058 | Not incident |
| Konings 2012 | 10.1017/S0033291711000973 | Not incident |
| Korhonen 2007 | 10.1017/s0033291706009639 | Not incident |
| Korhonen 2011 | 10.1093/ntr/ntq251 | Not incident |
| Korn 2018 | 10.1016/j.drugpo.2018.05.003 | Not incident |
| Kounali 2014 | 10.1017/s0033291714000026 | Not incident |
| Lavallee 2021 | 10.1016/j.abrep.2021.100347 | Not incident |
| Lee 2017 | 10.1080/10550887.2017.1303958 | Not incident |
| Lee 2019 | 10.1080/08897077.2019.1572047 | Not incident |
| Lee 2019 | 10.1080/09687637.2019.1698518 | Not incident |
| Leventhal 2008 | 10.1080/00952990802013367 | Not incident |
| Lien 2009 | 10.1016/j.jadohealth.2009.04.011 | Not incident |

| | | |
|---|---|---|
| Lyness 2000 | 10.1176/appi.ajp.157.9.1499 | Not incident |
| Manrique-Garcia 2012 | 10.1017/s0033291711002078 | Not incident |
| Marmorstein 2011 | 10.1016/j.addbeh.2011.02.006 | Not incident |
| Marwaha 2018 | 10.1093/schbul/sbx158 | Not incident |
| Mather 2015 | 10.1097/JOM.0000000000000504 | Not incident |
| McGee 2000 | 10.1046/j.1360-0443.2000.9544912.x | Not incident |
| Nelson 2018 | 10.1177/2333721418766127 | Not incident |
| Paffenbarger 1994 | 10.1111/j.1600-0447.1994.tb05796.x | Not incident |
| Pahwa 2012 | 22762903 | Not incident |
| Pasco 2008 | 10.1192/bjp.bp.107.046706 | Not incident |
| Patrick 2020 | 10.1016/j.drugalcdep.2020.108018 | Not incident |
| Patton 2002 | 10.1136/bmj.325.7374.1195 | Not incident |
| Patton 2006 | 10.1016/j.jadohealth.2005.11.027 | Not incident |
| Rasic 2013 | 10.1016/j.drugalcdep.2012.09.009 | Not incident |
| Rössler 2007 | 10.1016/j.schres.2007.01.002 | Not incident |
| Rössler 2012 | 10.1111/j.1360-0443.2011.03760.x | Not incident |
| Ruggles 2017 | 10.1007/s10461-016-1492-9 | Not incident |
| Schaefer 2021 | 10.1073/pnas.2013180118 | Not incident |
| Schoeler 2018 | 10.1017/s0033291717003658 | Not incident |
| Shidhaye 2010 | 10.1093/ije/dyq179 | Not incident |
| Silberg 2003 | 10.1111/1469-7610.00153 | Not incident |
| Silins 2014 | 10.1016/s2215-0366(14)70307-4 | Not incident |
| Steuber 2006 | 10.1016/j.addbeh.2005.04.010 | Not incident |
| Strong 2014 | 10.1007/s11524-013-9849-0 | Not incident |
| Tait 2012 | 10.1017/s1041610212000087 | Not incident |
| Talati 2016 | 10.1038/mp.2015.224 | Not incident |
| vanGool 2007 | 10.2105/ajph.2004.053199 | Not incident |
| Welham 2009 | 10.1017/s0033291708003760 | Not incident |
| Weller 1985 | 10.1176/ajp.142.7.848 | Not incident |
| Werneck 2022 | 10.1016/j.maturitas.2021.09.010 | Not incident |
| Windle 2001 | 10.1037/0022-006X.69.2.215 | Not incident |
| Windle 2004 | 10.1017/s0954579404040118 | Not incident |
| Woo 2002 | 10.1159/000058356 | Not incident |
| Woodruff 2010 | 10.1093/ntr/ntq007 | Not incident |
| Woodward 2016 | 10.1007/s10519-016-9812-4 | Not incident |
| Yoo 2016 | 10.1136/bmjopen-2015-008570 | Not incident |
| Bildt 2002 | 10.1007/s00420-001-0299-8 | Ineligible outcome |
| Bovasso 2001 | 10.1176/appi.ajp.158.12.2033 | Ineligible outcome |
| Cougnard 2007 | 10.1017/s0033291706009731 | Ineligible outcome |
| Federman 1997 | 10.1016/s0376-8716(96)01317-8 | Ineligible outcome |
| Gage 2014 | 10.1017/s0033291714000531 | Ineligible outcome |
| Goodman 2010 | 10.1111/j.1360-0443.2010.02981.x | Ineligible outcome |
| Gubata 2013 | 10.1016/j.jpsychores.2013.04.003 | Ineligible outcome |
| Hamer 2010 | 10.1001/archgenpsychiatry.2010.76 | Ineligible outcome |
| Henquet 2006 | 10.1016/j.jad.2006.05.002 | Ineligible outcome |
| Kuepper 2011 | 10.1017/s0033291711000511 | Ineligible outcome |
| Kuepper 2011 | 10.1136/bmj.d738 | Ineligible outcome |

| | | |
|---|---|---|
| Markota 2018 | 10.1016/j.jaac.2018.09.025 | Ineligible outcome |
| Miettunen 2014 | 10.1017/s0033291713002328 | Ineligible outcome |
| Patel 2006 | 10.1192/bjp.bp.106.022558 | Ineligible outcome |
| Samuelsson 2013 | 10.1186/1471-2458-13-621 | Ineligible outcome |
| Shang 2020 | 10.3390/cancers12061700 | Ineligible outcome |
| Smith 2014 | 10.1097/adm.0000000000000050 | Ineligible outcome |
| Suwazono 2003 | 10.1093/occmed/kqg102 | Ineligible outcome |
| Tien 1990 | 10.1097/00005053-199017880-00001 | Ineligible outcome |
| Tijssen 2010 | 10.1111/j.1600-0447.2010.01539.x | Ineligible outcome |
| vanOs 2021 | 10.1093/schbul/sbab019 | Ineligible outcome |
| Wang 2022 | 10.1016/j.jpsychires.2022.10.040 | Ineligible outcome |
| Wang 2022 | 10.1016/j.jad.2022.04.064 | Ineligible outcome |
| Wiles 2006 | 10.1192/bjp.bp.105.012179 | Ineligible outcome |
| Wu 1999 | 10.2105/ajph.89.12.1837 | Ineligible outcome |
| Zammit 2011 | 10.1192/bjp.bp.111.091421 | Ineligible outcome |
| Brown 1996 | 10.1097/00005768-199602000-00012 | Ineligible exposure |
| Frisher 2005 | 10.1136/jech.2004.030833 | Ineligible exposure |
| Fukunaga 2019 | 10.2188/jea.JE20190018 | Ineligible exposure |
| Bach 2021 | 10.1016/j.psychres.2021.114109 | Ineligible comparator |
| Breslau 1993 | 10.1001/archpsyc.1993.01820130033006 | Ineligible comparator |
| Chou 2011 | 10.4088/JCP.09m05618gry | Ineligible comparator |
| Giordano 2015 | 10.1017/s0033291714001524 | Ineligible comparator |
| Griesler 2008 | 10.1097/CHI.0b013e318185d2ad | Ineligible comparator |
| Griesler 2011 | 10.1111/j.1360-0443.2011.03403.x | Ineligible comparator |
| Keerthy 2021 | 10.1017/S003329172100386X | Ineligible comparator |
| Kinnunen 2006 | 10.2190/g652-t403-73h7-2x28 | Ineligible comparator |
| Nay 2013 | 10.1016/j.psychres.2013.03.006 | Ineligible comparator |
| Nielsen 2017 | 10.1017/s0033291717000162 | Ineligible comparator |
| Pacek 2013 | 10.1016/j.jad.2012.11.059 | Ineligible comparator |
| Sullivan 2018 | 10.1001/jamanetworkopen.2018.5174 | Ineligible comparator |
| Martín-Santos 2010 | 10.1111/j.1369-1600.2009.00180.x | No OR/HR/RR/IRR |
| Wade 2005 | 10.3138/cjccj.47.4.619 | No OR/HR/RR/IRR |
| Kirli 2016 | 10.1016/j.eurpsy.2016.01.526 | Population |
| Larson 2009 | 10.7205/milmed-d-02-0308 | Population |
| Galambos 2004 | 10.1080/01650250344000235 | Missing data |
| Green 1992 | 10.1111/j.1600-0447.1992.tb03254.x | Missing data |
| Kivelä 1996 | 10.1002/(SICI)1099-1166(199610)11:10<871::AID-GPS396>3.0.CO;2-6 | Missing data |
| Lewinsohn 1994 | 10.1037/0021-843X.103.2.302 | Missing data |
| Pedersen 2008 | 10.1111/j.1600-0447.2008.01259.x | Missing data |
| Takeuchi 2004 | 10.1539/joh.46.489 | Missing data |
| Blanco 2016 | 10.1001/jamapsychiatry.2015.3229 | Cohort overlap |
| Chang 2016 | 10.1016/j.jagp.2016.07.008 | Cohort overlap |
| Cheng 2016 | 10.1016/j.jad.2016.02.023 | Cohort overlap |
| Chigogora 2018 | https://discovery.ucl.ac.uk/id/eprint/10046397/ | Cohort overlap |
| Denissoff 2022 | 10.1016/j.schres.2022.06.014 | Cohort overlap |

| | | |
|---|---|---|
| Denissoff 2022 | 10.1111/add.15881 | Cohort overlap |
| Ernst 2021 | 10.1038/s41598-021-81927-9 | Cohort overlap |
| Fann 2022 | 10.3390/ijerph19127300 | Cohort overlap |
| Khaled 2012 | 10.1016/j.jpsychires.2011.11.011 | Cohort overlap |
| McDowell 2018 | 10.1093/ije/dyy141 | Cohort overlap |
| McLachlan 2006 | https://central.bac-lac.gc.ca/.item?id=MR19113&op=pdf&app=Library&oclc_number=298133976 | Cohort overlap |
| Meng 2014 | 10.1016/j.jad.2014.02.007 | Cohort overlap |
| Mo 2007 | https://rc.library.uta.edu/uta-ir/handle/10106/682 | Cohort overlap |
| Mojtabai 2013 | 10.2105/ajph.2012.300911 | Cohort overlap |
| Moon 2010 | 10.1007/s10560-010-0212-y | Cohort overlap |
| Munafò 2016 | 10.1016/j.drugalcdep.2016.04.035 | Cohort overlap |
| Patel 2018 | 10.1097/psy.0000000000000583 | Cohort overlap |
| Sánchez-Villegas 2008 | 10.1157/13117850 | Cohort overlap |
| Tellez 2019 | https://academicworks.cuny.edu/jj_etds/122/ | Cohort overlap |
| Wen 2019 | 10.1136/bmjopen-2019-029529 | Cohort overlap |
| Zammit 2010 | 10.1192/bjp.bp.109.070904 | Cohort overlap |

**eTable 2.** Tobacco and mood disorder study characteristics

| Record; Country | Cohort | Sample size | Follow-up length | Population | Baseline age | Exposure definition | Outcome subtype | Outcome definition | Covariates |
|---|---|---|---|---|---|---|---|---|---|
| **Albers 2002**<br><br>USA | Massachusetts Tobacco Survey of Youth | 503 | 4 years | General | 13.49 years<br><br>Adolescents | Ever smoking vs. never smoking | Depression | Kandel & Davies six-item scale ≥29 | Age, sex, race, parental education, presence of household smoker, baseline depression level and rebelliousness |
| **Almeida 2013**<br><br>Australia | Health in Men Study (HIMS) | 4636 | ~5.7 years | General | 71.5 years (SD 4.1)<br><br>Middle/Older | Current smoking vs non-smoking | Depression | GDS-15 ≥7 | Crude |
| **An 2015**<br><br>USA | Health and Retirement Study (HRS) | 24759 | ≤20 years | General | 60.5 years (95% CI 60.3 - 60.6)<br><br>Middle/Older | Current smoking vs. non-smoking | Depression | CES-D-8 ≥3 | Gender, ethnicity, education level, birth cohort, history of any psychiatric problem, alcohol consumption, age in years, marital status, household net wealth, diagnosis of chronic condition, residential region, BMI |
| **Armstrong 2017**<br><br>USA | Johns Hopkins Precursors Study (JHPS) | 821 | ≤63 years | Medical students | 26.3 years (SD 2.3)<br><br>Young adult | Ever smoking vs. never smoking | Depression | Self-reported depression, antidepressant use or lifetime history of mental health treatment | Sex, race, baseline age, enrollment wave, lack of physical activity, heavy alcohol use |
| **Bakhshaie 2015**<br><br>USA | Midlife Development in the United States Survey | 1406 | 10 years | General | 43.8 years (SD 13.5)<br><br>Middle/Older | Persistent smoking (daily smoking at baseline and follow-up) vs. | Depression | CIDI-SF (DSM-IV) | Age, gender and alcohol/drug use problems |

| | | | | | | | | | |
|---|---|---|---|---|---|---|---|---|---|
| | (MIDUS) | | | | | never smoking | | | |
| **Bolstad 2022**<br><br>Finland | Northern Finland Birth Cohort Study (NFBC86) | 5279 | ≤17years | General | 15-16 years<br><br>Adolescents | Daily smoking vs. no daily smoking | Depression | Major Depression measured by ICD-10 codes recorded in national health care registers | Age, sex, lifetime parental psychiatric disorder, family structure, illicit susbtance use, alcohol use, externalizing and internalizing symptoms |
| **Beutel 2019**<br><br>Germany | Gutenberg Health Study (GHS) | 9943 | 5 years | General | 54.3 years (SD 10.9)<br><br>Middle/Older | Current smoking vs. non-smoking | Depression | PHQ-9 ≥10 | Crude estimate extracted |
| **Borges 2018**<br><br>Mexico | Mexican Adolescent Mental Health Survey (MAMHS) | 928 | 8 years | General | 12-17 years<br><br>Adolescent | Ever smoking vs. never smoking | Any mood disorder | WMH-CIDI | Age/cohort, sex, alcohol use, any other drug use, anxiety disorders, disruptive behaviour |
| **Bots 2008**<br><br>Finland, Italy, Netherlands | The Finland, Italy and Netherlands Elderly (FINE) Study | 526 | 5 years | General | 75.2 years<br><br>Middle/Older | Current smoking vs. non-smoking | Depression | ZSDS-20 ≥48/80 | Age |
| **Breslau 1998**<br><br>USA | Detroit Epidemiologic Study | 843 | 5 years | General | 26 years<br><br>Young adult | Ever daily smoking (≥1 CPD for 1+ month) vs. never daily smoking | Depression | NIMH-DIS (DSM-III-R) | Sex, early conduct problems, history of alcohol dependence |
| **Brown 1996**<br><br>USA | Oregon Adolescent Depression Project | 1471 | ~2 years | General | 16.6 years (SD 1.2)<br><br>Adolescent | Regular smoking (≥3 times per week) vs. non-regular smoking (≤2 times per week) | Depression | K-SADS and LIFE | Age, gender, race, number of biological parents in household, parental education, other psychiatric disorders (i.e. anxiety, disruptive |

| | | | | | | | | | |
|---|---|---|---|---|---|---|---|---|---|
| | | | | | | | | | behaviour disorders, alcohol dependence and other drug dependence) |
| **Cabello 2017**<br><br>Ghana, India, Mexico, Russia | Study on Global AGEing and Adult Health (SAGE) | 5970 | 3-8 years | General | ≥50 years<br><br>Middle/Older | Daily smoking vs. non-smoking | Depression | CIDI | Age, sex, country, education level, employment, household income, presence of physical health condition, BMI, general health status, country, alcohol consumption, physical inactivity |
| **Chang 2016**<br><br>USA | Nurses Health Study (NHS) | 21728 | ≤10 years | Nurses | 71.4 years (SD 4.1)<br><br>Middle/Older | Current smoking (≥ 25 CPD) vs. never smoking | Depression | Self-reported clinician diagnosis, SSRI use or depressive symptoms CES-D-10 ≥10 or GDS-15 ≥6 | Age, sex, education level, ethnicity, social network, social class, regular caregiving to children/grandchildre n, regular caregiving to disabled/ill relatives, BMI, Mediterranean diet score, activity level, daily alcohol consumption, medical comorbidity, daily hours of sleep, sleeping difficulties, bodily pain, physical/functional limitation |
| **Chin 2016**<br><br>China | Chinese Primary Care Patients | 2540 | ≤1 year | General | 49.3 years (SD 7.0)<br><br>Adult | Smoker vs. non-smoker | Depression | PHQ-9 ≥10 | Sex, age, marital status, income, education level, employment status, alcohol consumtpion (y/n), exercise, comorbidities, family |

| | | | | | | | | | |
|---|---|---|---|---|---|---|---|---|---|
| | | | | | | | | | history of mental illness, visits to medical practitioners, area of residence, physician factors (e.g. sex, training) |
| **Chireh 2019**<br><br>Canada | National Population Health Survey (NPHS) | 2,743 | 10 years | General | >45 years<br><br>Middle/Older | Daily smoking vs. no daily smoking | Depression | CIDI-SF (DSM-III-R) | Sex, age, household income, ethnicity, high blood pressure, physical activity, BMI, family stress, traumatic events, chronic disease, heart disease |
| **Choi 1997**<br><br>USA | National Teenage Attitudes and Practices Survey (TAPS) | 6863 | ~4 years | General | 12 - 18 years<br><br>Adolescent | Past-month smoking vs. never smoking | Depression | Kandel & Davies six-item scale ≥29 | Age, sex, household income, perceived school performance, availability of social support, rebelliousness, participation in organised sports, ethnicity, parental educational level |
| **Clark 2007**<br><br>UK | Research with East London Adolescents: Community Health Survey (RELACHS) | 1170 | 2 years | School students | 11 - 14 years<br><br>Adolescent | Ever smoking vs. never smoking | Depression | sMFQ ≥8 | Age, gender, eligibility for free school meals, ethnicity, alcohol and drug-health risk behaviours, general health status, long-standing illness, overweight |
| **Cougle 2015**<br><br>USA | National Epidemiologic Study of Alcohol and Related | 27769 | ~3 years | General | ≥18 years<br><br>Adults | Past-year weekly smoking vs. past-year no weekly smoking | Any mood disorder | AUDADIS-IV (DSM-IV) | Age, income, marital status, gender, ethnicity, education, psychiatric comorbidity (i.e. |

| | | | | | | | | | |
|---|---|---|---|---|---|---|---|---|---|
| | Conditions (NESARC) | | | | | | | | anxiety, disorder, depressive disorder, personality disorder, bipolar disorder), alcohol use and cannabis/nicotine use |
| **Cuijpers 2007**<br><br>Netherlands | Netherlands Mental Health Survey and Incidence Study (NEMESIS) | 2726 | 2 years | General | 18 - 64 years<br><br>Adults | Past year smoking vs. no past year smoking | Any mood disorder | CIDI (DSM-III-R) | Age, gender, education level, employment status, neuroticism, locus of control, presence of somatic illness, parental history of psychopathology, childhood trauma (i.e. emotional neglect, psychological abuse, physical abuse, sexual abuse), lifetime psychiatric condition |
| **Flensborg-Madsen 2011**<br><br>Denmark | Copenhagen City Heart Study (CCHS-I-III)) | 17814 | ≤26 years | General | >20 years<br><br>Adults | Current smoking (max. >20g per/day) vs. never smoking | Depression | ICD code for depression diagnosis recorded in national registers | Age, sex, alcohol use, education, income, number of children living at home, marital status, physical activity level |
| **Fonseca 2021**<br><br>Brazil | Longitudinal Study on the Lifestyle and Health of Unviersity Students (ELESEU) | 1034 | ≤3 years | University students | 16 - 25 years<br><br>Adolescent | Past-month smoking vs. no past-month smoking | Depression | PHQ-9 ≥9 | Sex |
| **Ford 1998**<br><br>USA | John Hopkins Precursors Study | 1190 | ≤37 years | Medical students | 26 years<br><br>Young adult | Smoking vs. non-smoking | Depression | Self-reported diagnosis, treatment or antidepressant | Sex |

| | | | | | | | | | |
|---|---|---|---|---|---|---|---|---|---|
| | (JHPS) | | | | | | | medication | |
| **Gage 2015**<br><br>UK | The Avon Longitudinal Study of Parents and Children (ALSPAC) | 4561 | ~2 years | General | ~16 years<br><br>Adolescent | Frequency of use (max. daily) vs. never smoking | Depression | CIS-R | Family history of depression, gender, urban dwelling, maternal education, childhood borderline personality, childhood IQ, childhood psychotic experiences, conduct disorder group membership, peer problems, victimisation, cannabis use, alcohol, other illicit drugs, depression at age 12 (anxiety model only) |
| **Gentile 2021**<br><br>Sweden | Swedish National Military Conscription Register | 24041 | ≤48 years | Military conscripts | 18.1 years (SD 0.5)<br><br>Young adult | Current smoking (max.>20 CPD) vs. non-smoking | Depression | ICD code for diagnosis or antidepressant treatment | Age, sex, BMI, handgrip strength, verbal comprehension, alcohol consumption, other substance use |
| **Goodman 2000**<br><br>USA | National Longitudinal Study of Adolescent to Adult Health (Add Health) | 8704 | 1 year | School students | 15.48 (SD 1.59)<br><br>Adolescent | Past month smoking vs never smoking | Depression | CES-D-20 ≥22/24; modified 18-item version of CES-D-20 | Age, gender, race, parental education level, baseline depression score, smoking level at follow-up, teenager poor self-rated health, GPA, other drug use, alcohol use, delinquency, lower self-esteem, temperament, parental problematic alcohol use, parental |

| | | | | | | | | | |
|---|---|---|---|---|---|---|---|---|---|
| | | | | | | | | | understanding of teenager and perception of teenagers life, parental relationship with teenager and satisfaction with relationship |
| **Goodwin 2013**<br><br>USA | Ohio Army National Guard Health Initiative (OHARNG MHI) | 1391 | 1 year | National Guard Soldiers | 64.9% were under 35 years<br><br>Adult | Persistent smoking vs. never smoking | Depression | PHQ-9 ≥2 | Age,gender |
| **Groffen 2013**<br><br>USA | Health, Aging and Body Composition Study (Health ABC) | 2694 | ≤9 years | General | 73.6 years (SD 2.87)<br><br>Middle/Older | Current smoking vs. never smoking | Depression | Self-reported use of antidepressant medication and/or CESD-10 ≥11 | Age, sex, ethnicity, recruitment site, marital status and prevalent health conditions (e.g. CVD, diabetes, cancer) |
| **Hahad 2022**<br><br>Germany | Gutenberg Health Study (GHS) | 9937 | ~5 years | General | 54.9 years (SD 11.1)<br><br>Middle/Older | Pack-years of smoking (max. ≥30 years) vs. non-smoking | Depression | PHQ-9 ≥10 | Age, sex, SES, employment, shift work, retirement, relationship status, alcohol use, passive smoking, cardiovascular risk factors (e.g. BMI) and diseases, C-reactive protein |
| **Hiles 2015**<br><br>Australia | Hunter Community Study (HCS) | 1145 | ≤5.5 years | General | 65.6 years (SD 7.1)<br><br>Middle/Older | Current smoking vs. non-smoking | Depression | CES-D-20 ≥16 | Sex, age, inflammatory marker (IL-6), waist-to-hip ratio, steps per day, % energy from saturated fat, alcohol use, number of physical illnesses, |

| | | | | | | | | | |
|---|---|---|---|---|---|---|---|---|---|
| | | | | | | | | | quality of life |
| **Hoveling 2022**<br><br>Netherlands | Lifelines Cohort Study (LCS) | 76045 | ~3.8 years | General | 44.5 years (SD 12.0)<br><br>Adults | Current smoking vs. never smoking | Depression | MINI | Age, sex, socioeconomic position (index of years of education, household income, occupational prestige) |
| **Jackson 2019**<br><br>UK | English Longitudinal Study of Aging (ELSA) | 2019 | ≤12 years | General | 65.9 years (SD 9.34)<br><br>Middle/Older | Current smoking vs. never smoking | Depression | CES-D-8 ≥4 | Age, sex, ethnicity, household wealth, alcohol intake, BMI and physical activity |
| **Kang 2010**<br><br>Korea | Korea Welfare Panel Study (KoWePs) | 10125 | 6 months | Low-income | >20 years; 48.2% 20 – 44 years<br><br>Adult | Past year smoking (max. ≥ two packs/day) vs. past-year non-smoking | Depression | CES-D-11 ≥16 | Alcohol consumption, stressful life events, self-esteem, educational level, religion, marital status, urbanicity, household income, employment status, age, gender, depressive symptoms at baseline, health status |
| **Khaled 2012**<br><br>Canada | National Population Health Survey (NPHS) | 3824 | 12 years | General | NR<br><br>Adult | Current smoker (max. ‘heavy’/>20CPD) vs. never smoker | Depression | CIDI-SF (DSM-III-R) | Crude |
| **Kim 2022**<br><br>Korea | National Health Insurance Service - Health Screening Cohort | 88931 | ~7.7 years | General | >40 years; 38.3% 40 – 49 years<br><br>Middle/Older | Current smoking (max. ≥20 CPD) vs. never smoking | Depression | ICD code for diagnosis and record of at least one prescription for antidepressants | Crude |

| | | | | | | | | | |
|---|---|---|---|---|---|---|---|---|---|
| | (NHIS-<br>HEALS) | | | | | | | | |
| **Korhonen 2017**<br><br>Finland | Finnish Twin Cohort | 8963 | ≤10 years | Twins | 43.7 years (SD 7.70)<br><br>Middle/Older | Lifetime smoking (max. ≥20 CPD) vs. lifetime non-smoking | Depression | ICD code for at least 4 consecutive prescriptions of anti-depressants | Age, sex, social class, binge drinking, presence of somatic disease |
| **Lam 2005**<br><br>China | Hong Kong Secondary Schools | 1409 | 1 year | General | 12.7 years (SD 0.85)<br><br>Adolescent | Ever smoked vs. never smoked | Depression | 13-item custom scale 75th percentile | Age, sex |
| **Leung 2012**<br><br>Australia | Australian Longitudinal Study on Women's Health (ALSWH) | 5740 | ≤13 years | General | 18-23 years<br><br>Young adult | Current smoking (max. >20 CPD) vs lifetime non-smoking | Depression | CES-D-10 ≥10 | Sex, marital status, education level, employment status |
| **Luijendijk 2008**<br><br>Netherlands | The Rotterdam Study | 2801 | ~5 years | General | 71.0 years (SD 6.3)<br><br>Middle/Older | Current smoking vs. never smoking | Any mood disorder | CES-D-20 ³16 and diagnosis via PSE | Age, sex, education, income, disability in daily living, cognitive function, BMI, blood pressure, diabetes, cholesterol, cardiovascular disease, medication |
| **Meng 2017** | National Population Health Survey (NPHS) | 12227 | 16 years | General | >12 years<br><br>Adults | Regular smoking vs. no regular smoking | Depression | CIDI-SF | Crude estimate extracted |
| **Monroe 2021**<br><br>Ireland | The Irish Longitudinal Study on Ageing (TILDA) | 5309 | ≤6 years | General | 64.4 years (SD 9.1)<br><br>Middle/Older | Current smoking vs. never regular smoking | Depression | CIDI-SF | Age, sex, education level, marital status, physical activity, number of chronic conditions, number of cardiovascular conditions, number of physical limitations |

| | | | | | | | | | |
|---|---|---|---|---|---|---|---|---|---|
| **Monshouwer 2021**<br><br>Netherlands | NEMESIS-II | 4204 | ~3 years | General | 18 - 64 years<br><br>Adult | Frequency of use (max. >20 per day) vs no past-month use | Any mood disorder | CIDI (DSM-IV) | Age, gender, employment status, living with partner, education level, psychiatric comorbidity (i.e. anxiety disorder, SUD), physical activity, any somatic disorder, negative life events, childhood abuse |
| **Murphy 2003**<br><br>Canada | Stirling County Study (1970 – 1992) | 396 | 22 years | General | >18 years<br><br>Adult | Current smoking vs. non-smoking | Depression | DPAX-1 and DPAX-2; approximates DSM-III and DSMI-IV criteria via computerised algorithm | Crude |
| **Najafipour 2021**<br><br>Iran | Kerman Coronary Artery Disease Risk Factors Study (KERCADRS) | 2813 | 5 years | General | 46.2 years (SD 15.7)<br><br>Adult | Current smoking vs. non-smoking | Depression | BDI ≥31 | Crude |
| **do Nascimento 2015**<br><br>Brazil | Bambuí Cohort Study of Ageing (BCSA) | 701 | ≤11 years | General | 67.4 years (SD 6.1)<br><br>Middle/Older | Current smoking vs. never smoking | Depression | GHQ-12 ≥5 | Crude |
| **Park 2009**<br><br>Korea | Korea Youth Panel Survey (KYPS) | 1742 | 1 year | School students | 14 - 15 years<br><br>Adolescent | Continued smoking vs. never smoking | Depression | Six depressive symptoms listed in DSM-IV; cut-off: “greater than normal" | Age, gender, intact family, household income, parental education level, relationship with siblings, relationship |

| | | | | | | | | | |
|---|---|---|---|---|---|---|---|---|---|
| | | | | | | | | | with parents, parenting style, parental expectation towards education level, relationship with teachers, loneliness at school, number of friends, friends delinquency, self-esteem, number of stressors, life satisfaction, parental loss, difficulty inhibiting negative affect |
| **Raffetti 2019**<br><br>Sweden | The Kupol Cohort | 2900 | 1 year | School students | 13-14 years<br><br>Adolescent | Current tobacco use vs. no tobacco use | Depression | CES-DC ≥30 | Age, sex, alcohol consumption, parental education level, parent born abroad, depressive symptoms score at baseline |
| **Ren 2021**<br><br>China | China Health and Retirement Longitudinal Study (CHARLS) | 10508 | ~3 years | General | 57.62 years (SD 9.44)<br><br>Middle/Older | Current smoking vs. never smoking | Depression | CES-D-10 ≥10 | Age, educational level, BMI, urban/rural, marital status, alcohol consumption, digestive disease, cardiovascular disease, chronic kidney disease |
| **Rudaz 2017**<br><br>Switzerland | CoLaus/Psy CoLaus | 1524 | ~5.5 years | General | 51.4 years (SD 8.7)<br><br>Middle/Older | Daily smoking vs. no daily smoking | Depression | DIGS (DSM-IV) | Age, sex, SES, family history of depression, lifetime history of subthreshold depressive disorders (i.e. dysthymia, OSDD), alcohol use disorders, drug use |

| | | | | | | | | | |
|---|---|---|---|---|---|---|---|---|---|
| | | | | | | | | | disorders, anxiety disorders, BMI, dyslipidaemia, hypertension, diabetes, inflammatory markers (i.e. IL6, TNF-a, IL-1B, hs-CRP), childhood stressful events, adult life-event impact score, physical inactivity, neuroticism, problem-solving coping, help-seeking behaviours |
| **Sánchez-Villegas 2021**<br><br>Spain | Seguimiento University of Navarra (SUN) Cohort | 15096 | ~11 years | University graduates | 17.6% of sample were ≥50 years<br><br>Adults | Number of pack-years (max. ≥20) vs. never smoked | Depression | Self-reported physician-provided diagnosis of depression | Age, sex, education level, living alone, unemployment, BMI, physical activity, total energy intake, alcohol consumption, adherence to Mediterranean diet, personality traits |
| **Storeng 2020**<br><br>Norway | Nord-Trøndelag Health Studies (HUNT2 + HUNT3) | 4239 | ~11 years | General | 55-64 years<br><br>Middle/Older | Daily smoking (≥1 CPD) vs. non-regular smoking | Depression | HADS-D ≥8 | Age, sex, education level, marital status, presence of chronic illness, physical activity, alcohol consumption, disturbed sleep duration, excessive sitting time, low social participation |
| **Tanaka 2011**<br><br>Japan | The Komo-Ise Study | 8502 | 7 years | General | 40 - 69 years<br><br>Middle/Older | Current smoking vs. never smoking | Depression | DSM-12D ≥5 | Age, sex, rural/urban, education level, occupation, social network |

| | | | | | | | | | |
|---|---|---|---|---|---|---|---|---|---|
| **Tomita 2020**<br><br>South Africa | South African National Income Dynamics Study (SA-NIDS) | 14118 | ≤7 years | General | >15 years; 33.1% 35 – 64 years<br><br>Adult | Current smoking vs. non-smoking | Elevated symptoms | CES-D-10 ≥10 | Race, age, sex, marital status, education level, employment status, household income, rural/urban |
| **Tsai 2013**<br><br>Taiwan | Taiwan Longitudinal Study on Aging (TLSA) | 2145 | 8 years | General | >50 years; 36.5% 53 – 64 years<br><br>Middle/Older | Current smoking vs. never smoking | Depression | CES-D-10 ≥10 | Age, sex, level of education, psychological stress, diabetes, heart disease, IADL status, family support, audio acuity, betel quid chewing, alcohol consumption, tea consumption, physical activity |
| **Werneck 2022**<br><br>Brazil | Routine health evaluations in hospital | 4032 | ~3.1 years | General | 41.2 years (SD 7.9)<br><br>Adult | Current smoking vs. non-smoking | Depression | BDI ≥10 | Age, sex, length of follow-up, number of metabolic risk factors, self-rated health status, HS-CRP, physical activity, alcohol consumption |
| **Weyerer 2013**<br><br>Germany | German Study on Ageing, Cognition, Dementia (AgeCoDe Study) | 2512 | 3 years | General | 79.6 years (SD 3.5)<br><br>Middle/Older | Current smoking vs. non-smoking | Elevated symptoms | GDS ≥6 | Age, sex, living alone, marital status, education level, mobility impairment, vision impairment, hearing impairment, functional impairment, number of somatic comorbidities, mild cognitive impairment, subjective memory impairment, alcohol consumption, apoE4 |

| | | | | | | | | | |
|---|---|---|---|---|---|---|---|---|---|
| | | | | | | | | | status |
| **Zhang 2018**<br><br>Germany | Dresden Predictor Study (DPS) | 1157 | ~1.4 years | General | 21.03 years (SD 1.73)<br><br>Young adult | Current smoking vs. non-smoking | Depression | DIS (DSM-IV) | Sex, BMI, risk level of alcohol consumption, physical activity, good physical health |
| **Zimmerman 2009**<br><br>Mexico | Hispanic Established Population for the Epidemiologic Study of the Elderly (EPESE) | 964 | 2 years | General | 71.7 years (SD 5.6)<br><br>Middle/Older | Lifetime smoking (>100 cigarettes) vs. lifetime non-smoking | Depression | CES-D-20 ≥16 | Crude |

**SD =** standard deviation; **CPD =** cigarettes per day; **GDS** = Geriatric Depression Scale; **CES-D** = Centre for Epidemiologic Studies Depression Scale; **PHQ-9** = Patient Health Questionnaire; **WMH-CIDI** = World Mental Health Survey - Composite International Diagnostic Interview; **ZSDS** = Zung Self-Rating Depression Scale; **NIMH-DIS** = National Institute of Mental Health–Diagnostic Interview Schedule; **DSM-III-R** = Diagnostic and Statistical Manual of Mental Disorders, 3$^{rd}$ ed., Revised; **K-SADS** = Schedule for Affective Disorders and Schizophrenia for School-Age Children; **LIFE** = Longitudinal Interval Follow-up Evaluation; **CIDI** = Composite International Diagnostic Interview; **CIDI-SF** = Composite International Diagnostic Interview – Short Form; **sMFQ** = Short Mood and Feelings Questionnaire; **ICD** = International Classification of Diseases; **CIS-R** = The Clinical Interview Schedule-Revised; **DSM-IV =** Diagnostic and Statistical Manual of Mental Disorders, 4$^{th}$ ed.; **PSE =** Present State Examination; **DPAX-1/2 =** Depression and Anxiety Assessment; **BDI =** Beck Depression Inventory; **GHQ =** General Health Questionnaire; **CES-DC =** Center for Epidemiologic Studies Depression Scale for Children; **DIGS =** Diagnostic Interview for Genetic Studies; **HADS-D =** Hospital Anxiety and Depression Scale – Depression; **DSM-12D =** Diagnostic and Statistical Manual of Mental Disorders 12-Item Scale; **DIS =** Diagnostic Interview for Mental Disorders—Research Version

**eTable 3.** Tobacco and anxiety disorder study characteristics

| Record; Country | Cohort | Sample size | Follow-up length | Population | Baseline age | Exposure definition | Outcome subtype | Outcome definition | Covariates |
|---|---|---|---|---|---|---|---|---|---|
| **Borges 2018**<br><br>Mexico | MAMHS | 553 | 8 years | General | 12 – 17 years<br><br>Adolescent | Ever smoking (<15 years) vs. never smoking | Any anxiety disorder | WMH-CIDI (DSM-IV) | Crude |
| **Cougle 2015**<br><br>USA | NESARC | 28326 | ~3 years | General | ≥18 years<br><br>Adults | Past-year weekly smoking vs. past-year non-regular smoking | Any anxiety disorder | AUDADIS-IV (DSM-IV) | Age, income, marital status, gender, ethnicity, education, psychiatric comorbidity (i.e. depressive disorder, personality disorder, bipolar disorder), alcohol use and cannabis use |
| **Cuijpers 2007**<br><br>Netherlands | NEMESIS | 2726 | 2 years | General | 18 – 64 years<br><br>Adults | Past-year smoking vs past-year non-smoking | Any anxiety disorder | CIDI (DSM-III-R) | Age, gender, education level, employment status, neuroticism, locus of control, presence of somatic illness, parental history of psychopathology, childhood trauma (i.e. emotional neglect, psychological abuse, physical abuse, sexual abuse), lifetime psychiatric condition |
| **Gage 2015**<br><br>UK | ALSPAC | 4561 | ~2 years | General | ~16 years<br><br>Adolescent | Frequency of use (max. daily) vs. never smoking | Any anxiety disorder | CIS-R | Family history of depression, gender, urban dwelling, maternal education, childhoohd borderline personality, childhood IQ, childhood psychotic experiences, conduct disorder group membership, peer problems, victimisation, cannabis use, alcohol, other illicit drugs, depression at age 12 (anxiety model only) |
| **Isensee 2003** | EDSP | 2548 | 3.5 | General | 14-24 years | Tobacco use | GAD | DIA-X/M- | Age, sex |

| | | | | | | | | | |
|---|---|---|---|---|---|---|---|---|---|
| Germany | | | years | | Young adult | (max. non-dependent regular smoking) vs. never use | | CIDI | |
| **Hahad 2022**<br><br>Germany | GHS | 10324 | ~5 years | General | 54.9 years (SD 11.1) | Pack years of smoking (max. ≥30 years) vs. non-smoking | Anxiety | GAD-2 ≥3 | Age, sex, SES, employment, shift work, retirement, relationship status, alcohol use, passive smoking, cardiovascular risk factors (e.g. BMI) and diseases, C-reactive protein |
| **Monshouwer 2021**<br><br>Netherlands | NEMESIS-II | 4182 | ~3 years | General | 18 - 64 years<br><br>Adult | Frequency of use (max. >20 per day) vs no past-month use | Any anxiety disorder | CIDI (DSM-IV) | Age, gender, employment status, living with partner, education level, psychiatric comorbidity (i.e. mood disorder, SUD), physical activity, any somatic disorder, negative life events, childhood abuse |
| **Monroe 2021**<br><br>Ireland | TILDA | 5309 | ≤6 years | General | 64.4 years (SD 9.1)<br><br>Middle/Older | Current smoking vs. never regular smoking | GAD | CIDI-SF | Age, sex, education level, marital status, physical activity, number of chronic conditions, number of cardiovascular conditions, number of physical limitations |
| **Storeng 2020**<br><br>Norway | HUNT2 + HUNT3 | 3728 | ~11 years | General | 55-64 years<br><br>Middle/Older | Daily smoking (≥1 CPD) vs. non-regular smoking | Anxiety | HADS-A ≥8 | Age, sex, education level, marital status, presence of chronic illness, physical activity, alcohol consumption, disturbed sleep duration, excessive sitting time, low social participation |
| **Zvolensky 2008**<br><br>USA | OADP | 889 | NR | School students | 16.6 years (SD 1.2)<br><br>Adolescent | Current smoking vs non-smoking | Panic Disorder | LIFE (DSM-IV) | Crude |

**WMH-CIDI** = World Mental Health Survey - Composite International Diagnostic Interview; **CIS-R** = The Clinical Interview Schedule-Revised; **CIDI** = Composite International Diagnostic Interview; **HADS-A =** Hospital Anxiety and Depression Scale – Anxiety; **GAD-2** = Generalized Anxiety Disorder (2-Item); **LIFE** = Longitudinal Interval Follow-up Evaluation; **AUDADIS-IV** = Alcohol Use Disorder and Associated Disabilities Interview Schedule-IV

**eTable 4.** Tobacco and psychotic disorder study characteristics

| Record; Country | Cohort | Sample size | Follow-up length | Population | Baseline age | Exposure definition | Outcome subtype | Outcome definition | Covariates |
|---|---|---|---|---|---|---|---|---|---|
| **Kendler 2015**<br><br>Sweden | Swedish Conscript Registry (2002 – 2008) | 233879 | ~7.9 years | Military conscripts | 18.3 years (SD 0.6)<br><br>Young adult | Frequency of use (max. >20 CPD) vs. non-smoking | Schizophrenia | ICD code for schizophrenia diagnosis/ hospitalization | Parental education, neighbourhood deprivation, any drug abuse |
| **King 2020**<br><br>UK | The Health Improvement Network (THIN) | 907586 | ~6 years | General | 15 – 30 years<br><br>Young adult | Frequency of use (max. ≥ 20 CPD) vs. non-smoking | Schizophrenia | READ code for any non-affective psychosis | Sex, social deprivation (Townsend deprivation score) |
| **Mustonen 2018b**<br><br>Finland | NFBC-1986 | 4198 | ≤16 years | General | 15-16 years<br><br>Adolescents | Frequency of use (max. ≥10 CPD) vs. non-smoking | Any psychotic disorder | ICD code for any psychotic disorder diagnosis/ hospitalization | Age, cannabis use, alcohol consumption, other substance use, parental substance use, parental psychosis, number of psychotic experiences at baseline |
| **Okkenhaug 2018**<br><br>Norway | Young-HUNT1 | 60 | NR | School students | 16 years (SD 1.54)<br><br>Adolescents | Daily smoking (≥1 CPD) vs. non-daily smoking | Schizophrenia | ICD code for schizophrenia diagnosis/ hospitalization | Crude |
| **Weiser 2004**<br><br>Israel | Israel Defense Forces Medical Corps | 14248 | ~10.2 years | Military conscripts | 16-17 years<br><br>Adolescents | Frequency of use (max. ≥10 CPD) vs. non-smoking | Schizophrenia | ICD code for schizophrenia diagnosis/ hospitalization | Age, sex, other psychiatric disorder at baseline, social functioning, IQ, SES |
| **Zammit 2003**<br><br>Sweden | Swedish Conscript Registry (1969/1970) | 48772 | ≤27 years | Military conscripts | 18-20 years<br><br>Young adult | Frequency of use (max. >20 CPD) vs. non-smoking | Schizophrenia | ICD code for schizophrenia diagnosis/ hospitalization | Age, sex, IQ, cannabis use, poor social integartion, other psychiatric diagnoses, urbanicity, disturbed |

| | | | | | | | | | |
|---|---|---|---|---|---|---|---|---|---|
| | | | | | | | | | behaviour in childhood, family socioeconomic status, father's occupation, family psychiatric history, history of problematic alcohol use |

**eTable 5.** Cannabis and mood disorder study characteristics

| Record; Country | Cohort | Sample size | Follow-up length | Population | Baseline age | Exposure definition | Outcome subtype | Outcome definition | Covariates |
|---|---|---|---|---|---|---|---|---|---|
| **Danielsson 2016**<br><br>Sweden | Mental Health, Work and Relations Study (PART) | 6719 | ~3 years | General | 41.4 years (SD 19.1)<br><br>Adult | Ever use vs. never use | Depression | MDI ≥ 20 | Age, sex, other illicit drug use, alcohol consumption (AUDIT), education, place of upbringing, childhood adverse circumstances (i.e. economic deprivation and serious family tension), ethnicity, depression/anxiety at baseline |
| **Gage 2015**<br><br>UK | ALSPAC | 4561 | ~2 years | General | ~16 years<br><br>Adolescent | Lifetime frequency (cumulative) vs. never use | Depression | CIS-R | Family history of depression, gender, urban dwelling, maternal education, childhoohd borderline personality, childhood IQ, childhood psychotic experiences, conduct disorder group membership, peer problems, victimisation, cannabis use, alcohol, other illicit drugs, depression at age 12 (anxiety model only) |
| **Manrique-Garcia 2012**<br><br>Sweden | Swedish Conscript Registry | 45087 | ≤35 years | Military conscripts | 18-20 years<br><br>Young adult | Lifetime frequency (max. >50) vs. never use | Any mood disorder | ICD code for hospitalization | Age, sex, prior personality disorders, IQ, disturbed behaviour in childhood, social adjustment, risky use of alcohol, smoking, early |

| | | | | | | | | | |
|---|---|---|---|---|---|---|---|---|---|
| | | | | | | | | | adulthood socioeconomic position, use of other illicit substances, urbanicity |
| **Mustonen 2021**<br><br>Finland | NFBC-1986 | 5038 | ≤17 years | General | 15-16 years<br><br>Adolescent | Lifetime frequency (max. ≥5 times) vs. never use | Depression | ICD-10 code for depression drawn from various national registries | Externalizing symptoms, daily smoking, other illicit substance use, alcohol use, family structure, parental psychiatric disorder |
| **Paton 1977**<br><br>USA | New York Schools | 2400 | 6 months | School students | <18 years<br><br>Adolescent | Past-month use vs. past-month non-use | Elevated symptoms | Custom six-item scale (e.g. unhappiness, sadness) adapted from SCL ≥22 | |
| **Rognli 2019**<br><br>Norway | Young in Norway | 2468 | 9 years | General | 28.5 years<br><br>Young adult | Past-year use vs. never use | Depression | ATC-code for prescription of antidepressants | Age, gender, country of birth, parental education, living with biological parents in adolescence, conduct problems, alcohol intoxication episodes, daily smoking, other illicit drug use, mental distress |
| **vanLaar 2007**<br><br>Netherlands | Netherlands Mental Health Survey and Incidence Study (NEMESIS) | 3881 | 3 years | General | 39 years (SD 12.9)<br><br>Adult | Frequency of use (max. almost every day/daily) vs. no lifetime use (<5 times) | Any mood disorder | CIDI (DSM-II-R) | Gender, age, education, urbanicity, employment, partner status, neurotic personality, parental psychiatric history, traumatic events in chidlhood, lifetime alcohol-use disorders or other SUDs, lifetime psychotic symptoms and lifetime anxiety disorders |

**eTable 6.** Cannabis and anxiety disorder study characteristics

| Record; Country | Cohort | Sample size | Follow-up length | Population | Baseline age | Exposure definition | Outcome subtype | Outcome definition | Covariates |
|---|---|---|---|---|---|---|---|---|---|
| **Danielsson 2016**<br><br>Sweden | Mental Health, Work and Relations Study (PART) | 6720 | 3 years | General | 41.4 years (SD 19.1)<br><br>Adult | Ever use vs. never use | Anxiety | > 1.75 average on SCL-10 for Anxiety | Age, sex, other illicit drug use, alcohol consumption (AUDIT), education, place of upbringing, childhood adverse circumstances (i.e. economic deprivation and serious family tension), ethnicity, depression at baseline |
| **Feingold 2016**<br><br>USA | NESARC | 28505 | ~3 years | General | 18+ years<br><br>Adult | Past-year use (max. almost daily/daily) vs. past-year non-use | Any anxiety disorder | AUDADIS-IV (DSM-IV) | Sex, age, educational level, household income, marital status, urbanicity and region, alcohol use disorders and other (non-cannabis) SUDs), other psychiatric disorders at baseline (e.g. MDD). |
| **Gage 2015**<br><br>UK | ALSPAC | 4561 | ~2 years | General | ~16 years<br><br>Adolescent | Lifetime frequency (cumulative) vs. never use | Any anxiety disorder | CIS-R | Family history of depression, gender, urban dwelling, maternal education, childhoohd borderline personality, childhood IQ, childhood psychotic experiences, conduct disorder group membership, peer problems, victimisation, cannabis use, alcohol, other illicit drugs, depression at age 12 (anxiety model only) |
| **Mustonen 2021** | NFBC-1986 | 5038 | ≤17 years | General | 15-16 years | Lifetime frequency | Any anxiety | ICD-10 code for any | Externalizing symptoms, daily smoking, other illicit |

| | | | | | | | | | |
|---|---|---|---|---|---|---|---|---|---|
| Finland | | | | | Adolescent | (max. ≥5 times) vs. never use | disorder | anxiety disorder drawn from various national registries | substance use, alcohol use, family structure, parental psychiatric disorder |
| **Rognli 2019**<br><br>Norway | Young in Norway | 2468 | 9 years | General | 28.5 years<br><br>Young adult | Past-year use vs. never use | Any anxiety disorder | ATC-code for prescription of anxiolytics | Age, gender, country of birth, parental education, living with biological parents in adolescence, conduct problems, alcohol intoxication episodes, daily smoking, other illicit drug use, mental distress |
| **vanLaar 2007**<br><br>Netherlands | Netherlands Mental Health Survey and Incidence Study (NEMESIS) | 3854 | 3 years | General | 39 years (SD 12.9)<br><br>Adult | Frequency of use (max. almost every day/daily) vs. no lifetime use (<5 times) | Any anxiety disorder | CIDI (DSM-II-R) | Gender, age, education, urbanicity, employment, partner status, neurotic personality, parental psychiatric history, traumatic events in childhood, lifetime alcohol-use disorders or other SUDs, lifetime psychotic symptoms and lifetime mood disorders |
| **Zvolensky 2008**<br><br>USA | OADP | 889 | ~60 months | School students | 16.6 years (SD 1.2)<br><br>Adolescent | Lifetime use vs. lifetime non-use | Panic Disorder | LIFE (DSM-IV) | Age, non-cannabis drug dependence, cigarette smoking |

**eTable 7.** Cannabis and psychotic disorder study characteristics

| Record; Country | Cohort | Sample size | Follow-up length | Population | Baseline age | Exposure definition | Outcome subtype | Outcome definition | Covariates |
|---|---|---|---|---|---|---|---|---|---|
| **Mustonen 2018a**<br><br>Finland | NFBC-1986 | 5872 | ≤14 years | General | 15-16 years<br><br>Adolescent | Lifetime frequency (max. ≥5 times) vs. never use | Any psychotic disorder | ICD-10 code for any psychotic disorder drawn from various national registries | Age, prodromal psychosis, other substance use, alcohol use, daily smoking, parental psychosis |
| **Rognli 2019**<br><br>Norway | Young in Norway | 2468 | 9 years | General | 28.5 years<br><br>Young adult | Past-year use vs. never use | Any psychotic disorder | ATC-code for prescription of antipsychotics | Age, gender, country of birth, parental education, living with biological parents in adolescence, conduct problems, alcohol intoxication episodes, daily smoking, other illicit drug use, mental distress |
| **vanOs 2002**<br><br>Netherlands | NEMESIS | 4045 | 3 years | General | 18 – 64 years<br><br>Adult | Lifetime frequency (cumulative) vs. never use | Psychosis | Composite outcome of psychosis requiring treatment, determined via BPRS ≥4, CIDI and SCID (DSM-III-R) | Age, sex, ethnicity, marital status, education level, urbanicity and level of discrimination, other illicit drug use |
| **Zammit 2002**<br><br>Sweden | Swedish Conscript Registry | 40643 | ≤27 years | Military conscripts | 18- 20 years<br><br>Young adult | Lifetime frequency (max. >50 times) vs. never use | Schizophrenia | ICD code for schizophrenia diagnosis/ hospitalization recorded in Swedish National Hospital Discharge Register | Age, sex, other psychiatric diagnoses at conscription, IQ, poor social integration, disturbed behaviour, cigarette smoking and urbanicity |

**eTable 8.** NOS quality assessment

| Study | S1 | S2 | S3 | S4 | C1 | C2 | C3 | O1 | O2 | O3 | Rating | Score |
|---|---|---|---|---|---|---|---|---|---|---|---|---|
| **Albers 2002** | 1 | 1 | 0 | 1 | 0 | 0.5 | 0.5 | 1 | 1 | 0 | Moderate | 6 |
| **An 2015** | 1 | 1 | 0.5 | 1 | 1 | 0.5 | 0.5 | 1 | 1 | 0 | Moderate | 7.5 |
| **Armstrong 2017** | 0 | 1 | 0 | 1 | 1 | 0.5 | 0 | 0 | 1 | 0 | Moderate | 4.5 |
| **Bakhshaie 2015** | 1 | 1 | 0.5 | 1 | 1 | 0.5 | 0 | 1 | 1 | 0 | Moderate | 7 |
| **Bolstad 2022** | 1 | 1 | 1 | 1 | 1 | 0.5 | 0.5 | 1 | 1 | 1 | High | 9 |
| **Borges 2018** | 1 | 1 | 0 | 1 | 1 | 0.5 | 0 | 1 | 1 | 1 | Moderate | 7.5 |
| **Breslau 1998** | 1 | 1 | 0.5 | 1 | 1 | 0 | 0 | 1 | 1 | 1 | Moderate | 7.5 |
| **Brown 1996** | 1 | 1 | 1 | 1 | 1 | 0.5 | 0.5 | 1 | 1 | 0 | Moderate | 8 |
| **Cabello 2017** | 1 | 1 | 1 | 1 | 1 | 0.5 | 0.5 | 1 | 1 | 0 | Moderate | 8 |
| **Chang 2016** | 0 | 1 | 1 | 1 | 1 | 0.5 | 0.5 | 0 | 1 | 0 | Moderate | 8 |
| **Chin 2016** | 0 | 1 | 0 | 1 | 1 | 0.5 | 0.5 | 1 | 0 | 0 | Moderate | 5 |
| **Chireh 2019** | 1 | 1 | 1 | 1 | 0 | 0.5 | 0.5 | 1 | 1 | 0 | Moderate | 7 |
| **Choi 1997** | 1 | 1 | 0.5 | 1 | 0 | 0.5 | 0.5 | 1 | 1 | 0 | Moderate | 6.5 |
| **Clark 2007** | 1 | 1 | 0 | 1 | 1 | 0.5 | 0.5 | 1 | 1 | 1 | High | 8 |
| **Cougle 2015** | 1 | 1 | 0.5 | 1 | 1 | 0.5 | 0.5 | 1 | 1 | 1 | High | 8.5 |
| **Cuijpers 2007** | 1 | 1 | 0.5 | 1 | 0 | 0.5 | 0.5 | 1 | 1 | 1 | Moderate | 7.5 |
| **Danielsson 2016** | 1 | 1 | 0 | 1 | 1 | 0.5 | 0.5 | 1 | 1 | 1 | High | 8 |
| **Feingold 2015** | 1 | 1 | 1 | 1 | 1 | 0.5 | 0.5 | 1 | 1 | 1 | High | 9 |
| **Feingold 2016** | 1 | 1 | 1 | 1 | 1 | 0.5 | 0.5 | 1 | 1 | 1 | High | 9 |
| **Flensborg-Madsen 2011** | 1 | 1 | 1 | 1 | 1 | 0.5 | 0.5 | 1 | 1 | 1 | High | 9 |
| **Gage 2015** | 1 | 1 | 1 | 1 | 1 | 0.5 | 0.5 | 1 | 1 | 1 | High | 9 |
| **Gentile 2021** | 1 | 1 | 1 | 1 | 1 | 0.5 | 0.5 | 1 | 1 | 1 | High | 9 |
| **Goodman 2000** | 1 | 1 | 0.5 | 1 | 1 | 0.5 | 0.5 | 0 | 1 | 0 | Moderate | 6.5 |
| **Groffen 2013** | 1 | 1 | 0.5 | 1 | 0 | 0.5 | 0 | 0 | 1 | 0 | Low | 5 |
| **Hahad 2022** | 1 | 1 | 1 | 1 | 1 | 0.5 | 0.5 | 1 | 1 | 0 | Moderate | 8 |
| **Hiles 2015** | 0 | 1 | 0.5 | 1 | 1 | 0.5 | 0.5 | 1 | 1 | 0 | Moderate | 6.5 |
| **Hoveling 2022** | 1 | 1 | 0.5 | 1 | 0 | 0.5 | 0.5 | 1 | 1 | 0 | Moderate | 6.5 |
| **Jackson 2019** | 1 | 1 | 0.5 | 1 | 1 | 0.5 | 0.5 | 1 | 1 | 0 | Moderate | 7.5 |
| **Kang 2010** | 0 | 1 | 1 | 1 | 1 | 0.5 | 0.5 | 1 | 0 | 0 | Moderate | 6 |

| | | | | | | | | | | | | |
|---|---|---|---|---|---|---|---|---|---|---|---|---|
| **Kendler 2015** | 1 | 1 | 1 | 1 | 1 | 0.5 | 0 | 1 | 1 | 1 | Moderate | 8.5 |
| **King 2020** | 1 | 1 | 1 | 1 | 0 | 0 | 0.5 | 1 | 1 | 1 | Moderate | 7.5 |
| **Korhonen 2017** | 1 | 1 | 1 | 1 | 1 | 0.5 | 0.5 | 1 | 1 | 1 | High | 9 |
| **Leung 2012** | 0 | 1 | 1 | 1 | 0 | 0.5 | 0 | 1 | 1 | 0 | Low | 5.5 |
| **Luijendijk 2008** | 1 | 1 | 1 | 1 | 0 | 0.5 | 0.5 | 1 | 1 | 0 | Moderate | 6.5 |
| **Manrique-Garcia 2012** | 1 | 1 | 1 | 1 | 1 | 0.5 | 0.5 | 1 | 1 | 1 | High | 9 |
| **Monroe 2021** | 1 | 1 | 0.5 | 1 | 0 | 0.5 | 0.5 | 1 | 1 | 0 | Moderate | 6.5 |
| **Monshouwer 2021** | 1 | 1 | 1 | 1 | 1 | 0.5 | 0.5 | 1 | 1 | 0 | Moderate | 8 |
| **Mustonen 2018a** | 1 | 1 | 0 | 1 | 1 | 0 | 0 | 1 | 1 | 0.5 | Moderate | 6.5 |
| **Mustonen 2018b** | 1 | 1 | 1 | 1 | 1 | 0 | 0.5 | 1 | 1 | 0.5 | Moderate | 7.5 |
| **Mustonen 2021** | 1 | 1 | 0 | 1 | 1 | 0 | 0.5 | 1 | 1 | 1 | Moderate | 7.5 |
| **Park 2009** | 1 | 1 | 0 | 1 | 0 | 0.5 | 0.5 | 0 | 1 | 0 | Moderate | 5 |
| **Raffetti 2019** | 0 | 1 | 0.5 | 1 | 1 | 0.5 | 0.5 | 1 | 1 | 0 | Moderate | 6.5 |
| **Ren 2021** | 1 | 1 | 0.5 | 1 | 1 | 0.5 | 0.5 | 1 | 1 | 0 | Moderate | 7.5 |
| **Rognli 2019** | 1 | 1 | 0.5 | 1 | 1 | 0.5 | 0.5 | 1 | 1 | 0.5 | Moderate | 8 |
| **Rudaz 2017** | 1 | 1 | 1 | 1 | 1 | 0.5 | 0.5 | 1 | 1 | 0 | Moderate | 8 |
| **Sánchez-Villegas 2021** | 0 | 1 | 1 | 1 | 1 | 0.5 | 0.5 | 0 | 1 | 0 | Moderate | 6 |
| **Storeng 2020** | 1 | 1 | 1 | 1 | 1 | 0.5 | 0.5 | 1 | 1 | 0 | Moderate | 8 |
| **Tanaka 2011** | 1 | 1 | 0.5 | 1 | 0 | 0.5 | 0.5 | 0 | 1 | 0 | Moderate | 5.5 |
| **Tomita 2020** | 1 | 1 | 0.5 | 1 | 0 | 0.5 | 0 | 1 | 1 | 0 | Low | 6 |
| **Tsai 2013** | 1 | 1 | 0.5 | 1 | 1 | 0.5 | 0.5 | 1 | 1 | 0 | Moderate | 7.5 |
| **van Laar 2007** | 1 | 1 | 0 | 1 | 1 | 0.5 | 0.5 | 1 | 1 | 1 | High | 8 |
| **van Os 2002** | 1 | 1 | 1 | 1 | 1 | 0.5 | 0.5 | 1 | 1 | 1 | High | 9 |
| **Weiser 2004** | 1 | 1 | 1 | 1 | 0 | 0.5 | 0.5 | 1 | 1 | 1 | Moderate | 8 |
| **Werneck 2022** | 0 | 1 | 0.5 | 1 | 1 | 0.5 | 0.5 | 1 | 1 | 0 | Moderate | 6.5 |
| **Weyerer 2013** | 0 | 1 | 0.5 | 1 | 1 | 0.5 | 0.5 | 1 | 1 | 0 | Moderate | 6.5 |
| **Zammit 2002** | 1 | 1 | 1 | 1 | 1 | 0.5 | 0.5 | 1 | 1 | 1 | High | 9 |
| **Zammit 2003** | 1 | 1 | 1 | 1 | 1 | 0.5 | 0.5 | 1 | 1 | 1 | High | 9 |
| **Zhang 2018** | 1 | 1 | 0.5 | 1 | 1 | 0 | 0.5 | 1 | 1 | 0 | Moderate | 7 |
| **Zvolensky 2008** | 1 | 1 | 0 | 1 | 1 | 0 | 0 | 1 | 1 | 0 | Moderate | 6 |

**eTable 9.** Summary of NOS ratings and mean scores by exposure/outcome

| Exposure | Outcome | K | Low (%) | Moderate (%) | High (%) | Mean (SD) |
|---|---|---|---|---|---|---|
| Tobacco | Mood | 43 | 7% | 77% | 16% | 7.01 (1.00) |
| Tobacco | Anxiety | 7 | 0% | 29% | 71% | 7.93 (0.53) |
| Tobacco | Psychosis | 5 | 0% | 80% | 20% | 8.10 (0.52) |
| Cannabis | Mood | 7 | 0% | 29% | 71% | 8.21 (0.67) |
| Cannabis | Anxiety | 7 | 0% | 43% | 57% | 7.92 (0.67) |
| Cannabis | Psychosis | 4 | 0% | 50% | 50% | 8.13 (0.88) |
| | **Overall** | **59** | 5% | 68% | 27% | 7.35 (1.01) |

**eTable 10.** Luis Furuya-Kanamori (LFK) index and asymmetry rating for primary meta-analyses

| Exposure | Outcome | K | LFK Index Score | Asymmetry Rating [a] |
|---|---|---|---|---|
| Tobacco | Mood | 43 | 4.12 | Major |
| Tobacco | Anxiety | 7 | -1.68 | Minor |
| Tobacco | Psychosis | 5 | -3.86 | Major |
| Cannabis | Mood | 7 | -0.59 | No asymmetry |
| Cannabis | Anxiety | 7 | 1.89 | Minor |
| Cannabis | Psychosis | 4 | 1.73 | Minor |

[a] LFK index scores of ±1, between ±1 and ±2, or ±2 indicate ‘no asymmetry’, ‘minor asymmetry’, and ‘major asymmetry’ respectively.

**eTable 11.** Subgroup analyses for tobacco and mood disorders

| | K | RR | 95%CI | $I^2$ | *Q* | $p_{subgroup}$ |
|---|---|---|---|---|---|---|
| **Overall** | 43 | 1.39 | 1.30-1.47 | 61.2% | | |
| **Age Group** | | | | | 5.19 | 0.159 |
| Adolescent | 10 | 1.32 | 1.08-1.61 | 36.3% | | |
| Young adult | 5 | 1.51 | 1.37-1.66 | 0.0% | | |
| Adult | 9 | 1.53 | 1.30-1.81 | 73.1% | | |
| Middle/Older | 19 | 1.34 | 1.24-1.44 | 51.2% | | |
| **Exposure Type** | | | | | 1.23 | 0.267 |
| Status | 25 | 1.34 | 1.24-1.44 | 57.0% | | |
| Heaviness | 18 | 1.44 | 1.30-1.60 | 50.8% | | |
| **Exposure Measure** | | | | | 3.57 | 0.311 |
| Ever use | 5 | 1.35 | 1.10-1.65 | 0.0% | | |
| Current or recent use (e.g. past year) | 20 | 1.35 | 1.23-1.48 | 64.0% | | |
| Regular use (e.g. daily) | 8 | 1.31 | 1.22-1.41 | 1.8% | | |
| Frequency of use (e.g. CPD) | 10 | 1.55 | 1.32-1.82 | 54.3% | | |
| **Outcome Type** | | | | | 0.05 | 0.826 |
| Depression | 38 | 1.38 | 1.29-1.48 | 63.8% | | |
| Mood disorder | 5 | 1.36 | 1.21-1.54 | 9.6% | | |
| **Outcome Measure** | | | | | 1.72 | 0.788 |
| Self-reported | 2 | 1.37 | 1.06-1.76 | 0.0% | | |
| Scale | 20 | 1.32 | 1.20-1.46 | 44.3% | | |
| Interview | 14 | 1.43 | 1.30-1.57 | 41.4% | | |
| Composite | 3 | 1.35 | 1.10-1.66 | 45.3% | | |
| Registry code | 4 | 1.53 | 1.13-2.07 | 68.8% | | |
| **High Quality (NOS)** | | | | | 0.01 | 0.907 |
| No | 36 | 1.38 | 1.29-1.47 | 54.9% | | |
| Yes | 7 | 1.36 | 1.13-1.65 | 72.9% | | |
| **Confounder Matrix (>2)** [a] | | | | | 0.39 | 0.530 |
| Adequate ≤ 2 | 37 | 1.40 | 1.31-1.50 | 43.3% | | |
| Adequate > 2 | 6 | 1.32 | 1.11-1.58 | 61.6% | | |
| **Confounder Matrix (0-4)** [a] | | | | | 4.46 | 0.347 |
| Adequate = 0 | 5 | 1.22 | 1.05-1.42 | 0.0% | | |
| Adequate = 1 | 14 | 1.43 | 1.31-1.56 | 53.4% | | |
| Adequate = 2 | 18 | 1.42 | 1.25-1.62 | 45.6% | | |
| Adequate = 3 | 4 | 1.27 | 1.10-1.48 | 64.1% | | |
| Adequate = 4 | 2 | 1.60 | 0.67-3.81 | 74.3% | | |

[a] Subgroup analyses informed by confounder matrix were considered on adjustment for constructs *over and above* socio-demographics and psychiatric comorbidity due to the high and low, respectively, adjustment for these constructs. Adjustment for >2, represents adjustment for over half of the remaining constructs (i.e. SES, other substance use, psychological factors and lifestyle factors).

**eTable 12.** Meta-regression analyses for tobacco and mood disorders

| Moderator | K | Coefficient | SE | Z value | P value | *95% CI* |
|---|---|---|---|---|---|---|
| Follow-up length | 43 | 0.0000 | 0.0002 | 0.1121 | 0.9108 | -0.0004; 0.0005 |
| Sample size | 43 | 0.0000 | 0.0000 | 1.2354 | 0.2167 | -0.0000; 0.0000 |

**eFigure 1.** PRISMA flow-chart

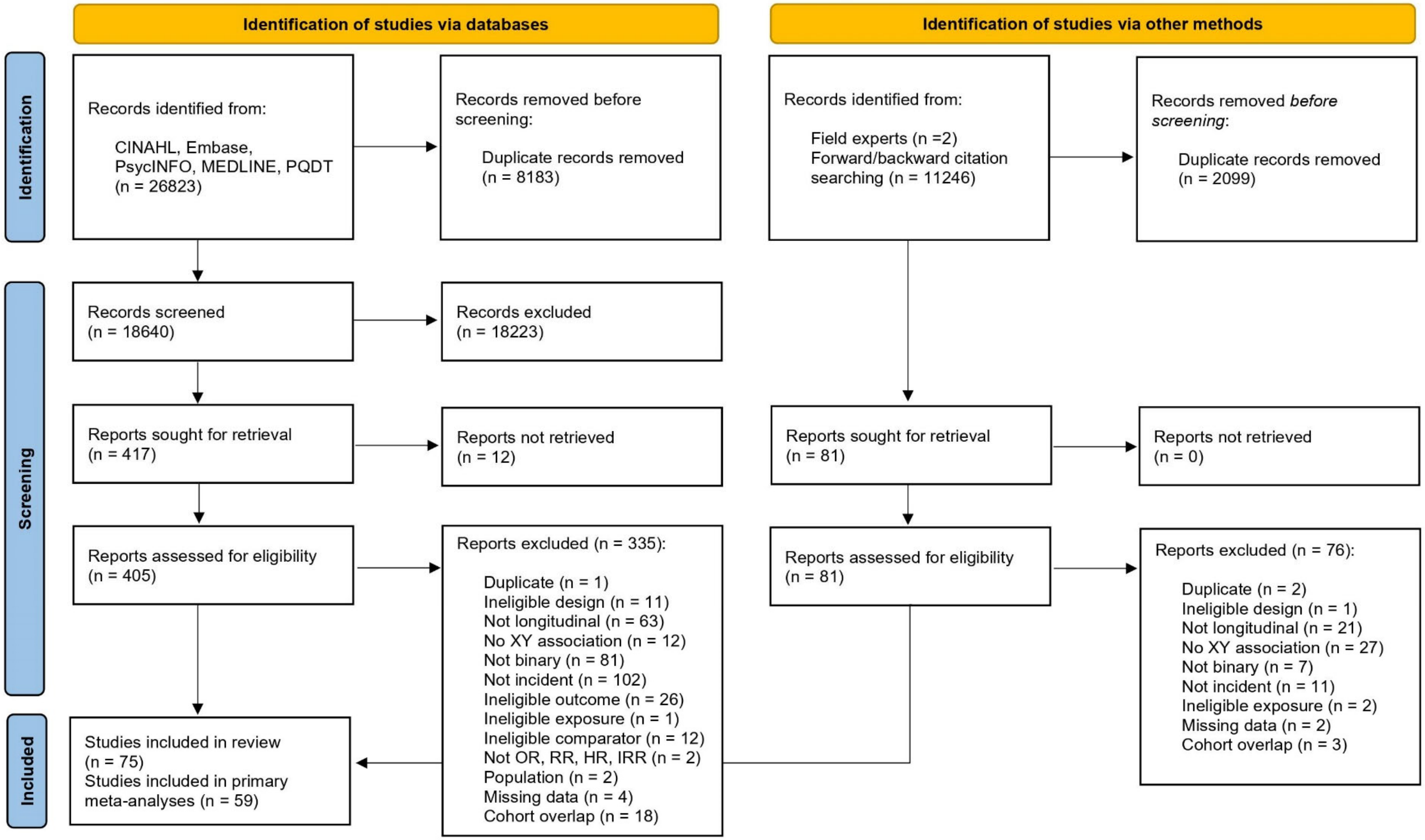

*From:* Page MJ, McKenzie JE, Bossuyt PM, Boutron I, Hoffmann TC, Mulrow CD, et al. The PRISMA 2020 statement: an updated guideline for reporting systematic reviews. BMJ 2021;372:n71. doi: 10.1136/bmj.n71. For more information, visit: http://www.prisma-statement.org/

**eFigure 2.** Unadjusted meta-analysis of tobacco use and mood disorders

| ID | log_RR | SE | RR | 95%−CI | Weight |
|---|---|---|---|---|---|
| Albers 2002 | 0.4637 | 0.2469 | 1.59 | [0.98; 2.58] | 1.9% |
| Almeida 2013 | 0.6471 | 0.1589 | 1.91 | [1.40; 2.61] | 2.8% |
| Bakhshaie 2015 | 0.8755 | 0.2678 | 2.40 | [1.42; 4.06] | 1.7% |
| Beutel 2019 | 0.4318 | 0.1082 | 1.54 | [1.25; 1.90] | 3.4% |
| Bolstad 2022 | 0.2469 | 0.1627 | 1.28 | [0.93; 1.76] | 2.8% |
| Borges 2018 | 0.3577 | 0.1654 | 1.43 | [1.03; 1.98] | 2.7% |
| Bots 2008 | 0.0677 | 0.3893 | 1.07 | [0.50; 2.29] | 1.1% |
| Breslau 1998 | 0.6471 | 0.2849 | 1.91 | [1.09; 3.34] | 1.6% |
| Brown 1996 | 0.8459 | 0.2828 | 2.33 | [1.34; 4.06] | 1.6% |
| Chang 2016 | 0.4121 | 0.1431 | 1.51 | [1.14; 2.00] | 3.0% |
| Flensborg−Madsen 2011 | 0.6931 | 0.1669 | 2.00 | [1.44; 2.77] | 2.7% |
| Fonseca 2021 | 0.1740 | 0.1209 | 1.19 | [0.94; 1.51] | 3.3% |
| Ford 1998 | 0.0953 | 0.1637 | 1.10 | [0.80; 1.52] | 2.8% |
| Gage 2015 | 0.4055 | 0.0910 | 1.50 | [1.25; 1.79] | 3.6% |
| Gentile 2021 | 0.4762 | 0.0488 | 1.61 | [1.46; 1.77] | 4.0% |
| Goodman 2000 | 0.8372 | 0.1192 | 2.31 | [1.83; 2.92] | 3.3% |
| Goodwin 2013 | 0.6419 | 0.2502 | 1.90 | [1.16; 3.10] | 1.9% |
| Hiles 2015 | 0.9594 | 0.3756 | 2.61 | [1.25; 5.45] | 1.1% |
| Jackson 2019 | 0.1570 | 0.1123 | 1.17 | [0.94; 1.46] | 3.4% |
| Kang 2010 | 0.5822 | 0.4118 | 1.79 | [0.80; 4.01] | 1.0% |
| Kim 2022 | −0.1054 | 0.0815 | 0.90 | [0.77; 1.06] | 3.7% |
| Korhonen 2017 | 0.4574 | 0.1403 | 1.58 | [1.20; 2.08] | 3.0% |
| Lam 2005 | 0.6831 | 0.1613 | 1.98 | [1.44; 2.72] | 2.8% |
| Luijendijk 2008 | 0.4511 | 0.3131 | 1.57 | [0.85; 2.90] | 1.4% |
| Meng 2017 | 0.3001 | 0.0788 | 1.35 | [1.16; 1.58] | 3.8% |
| Monroe 2021 | 0.5008 | 0.1703 | 1.65 | [1.18; 2.30] | 2.7% |
| Monshouwer 2021 | 1.3350 | 0.3641 | 3.80 | [1.86; 7.76] | 1.2% |
| Murphy 2003 | 0.2852 | 0.1972 | 1.33 | [0.90; 1.96] | 2.4% |
| Najafipour 2021 | −0.5621 | 0.7438 | 0.57 | [0.13; 2.45] | 0.4% |
| do Nascimento 2015 | 0.1222 | 0.1135 | 1.13 | [0.90; 1.41] | 3.4% |
| Raffetti 2019 | 0.4055 | 0.4571 | 1.50 | [0.61; 3.67] | 0.8% |
| Ren 2021 | 0.4886 | 0.0977 | 1.63 | [1.35; 1.97] | 3.6% |
| Rudaz 2017 | −0.1165 | 0.4233 | 0.89 | [0.39; 2.04] | 0.9% |
| Sánchez−Villegas 2021 | 0.4055 | 0.1299 | 1.50 | [1.16; 1.93] | 3.2% |
| Storeng 2020 | 0.2390 | 0.1355 | 1.27 | [0.97; 1.66] | 3.1% |
| Tanaka 2011 | 0.3507 | 0.3501 | 1.42 | [0.71; 2.82] | 1.2% |
| Tsai 2013 | 0.0296 | 0.1176 | 1.03 | [0.82; 1.30] | 3.3% |
| Werneck 2022 | 0.2852 | 0.1931 | 1.33 | [0.91; 1.94] | 2.4% |
| Weyerer 2013 | 0.2927 | 0.1338 | 1.34 | [1.03; 1.74] | 3.1% |
| Zhang 2018 | 0.6043 | 0.2553 | 1.83 | [1.11; 3.02] | 1.8% |
| Zimmerman 2009 | −0.5108 | 0.2581 | 0.60 | [0.36; 0.99] | 1.8% |
| **Random effects model** | | | **1.47** | **[1.34; 1.60]** | **100.0%** |
| **Prediction interval** | | | | **[0.92; 2.33]** | |

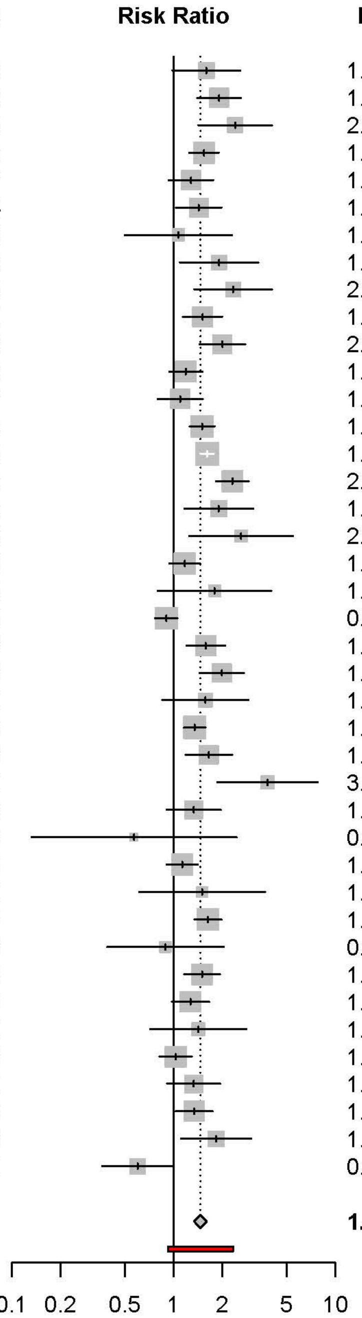

Heterogeneity: $I^2 = 69\%$, $\tau^2 = 0.0504$, $p < 0.01$

**eFigure 3.** Unadjusted meta-analysis of tobacco use and anxiety disorders

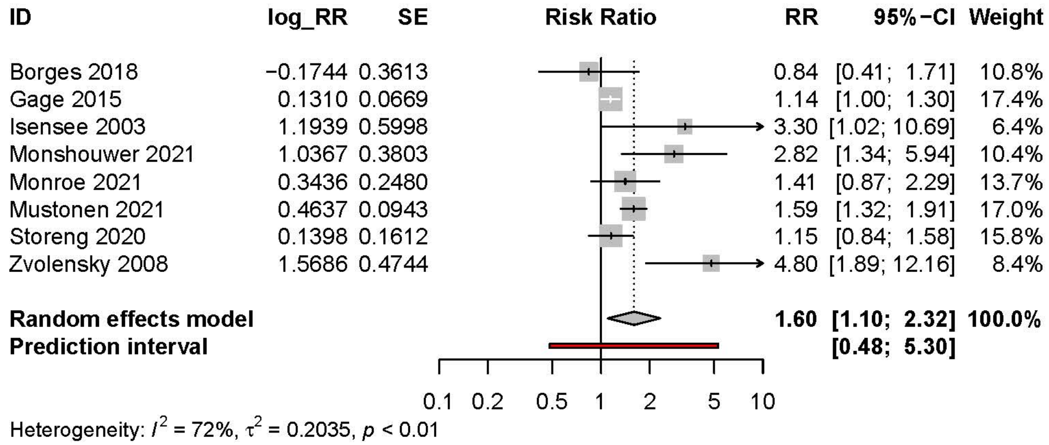

**eFigure 4.** Unadjusted meta-analysis of tobacco use and psychotic disorders

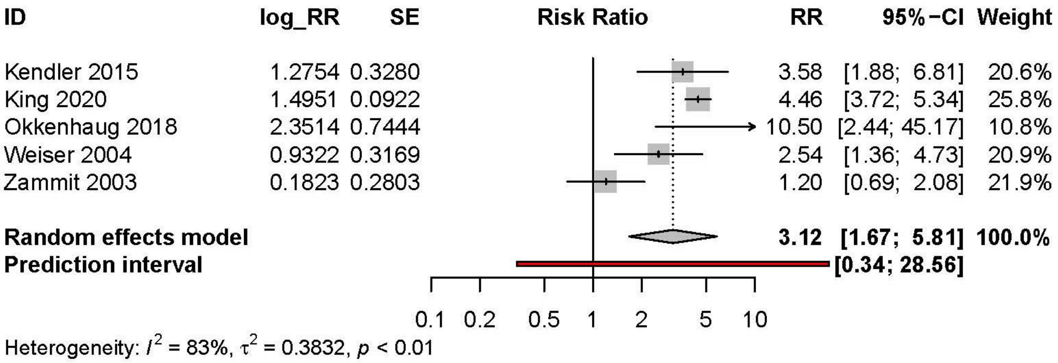

**eFigure 5.** Unadjusted meta-analysis of cannabis use and mood disorders

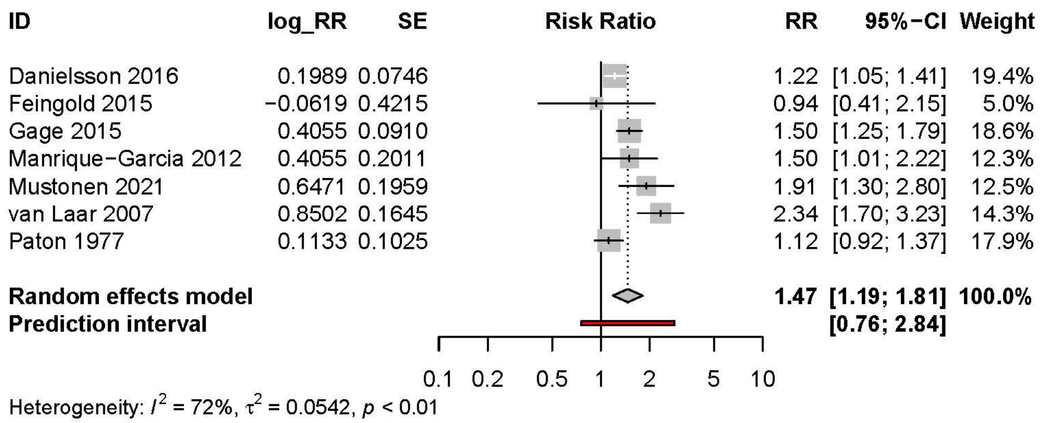

**eFigure 6.** Unadjusted meta-analysis of cannabis use and anxiety disorders

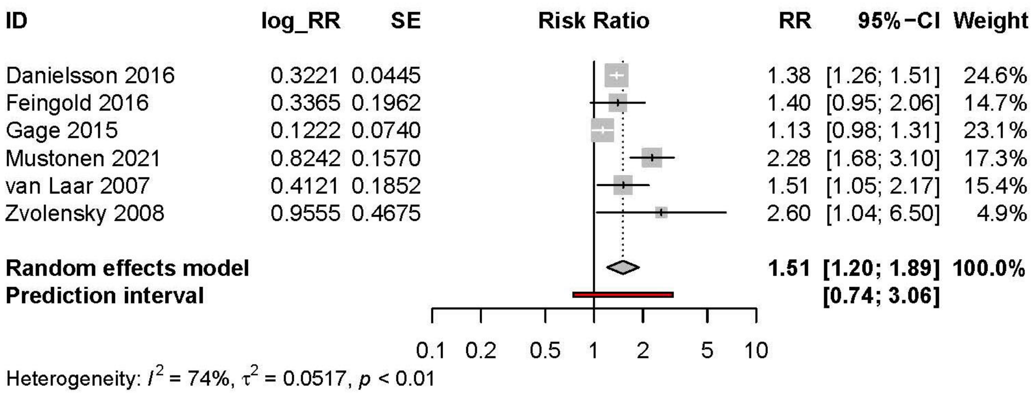

**eFigure 7.** Unadjusted meta-analysis of cannabis use and psychotic disorders

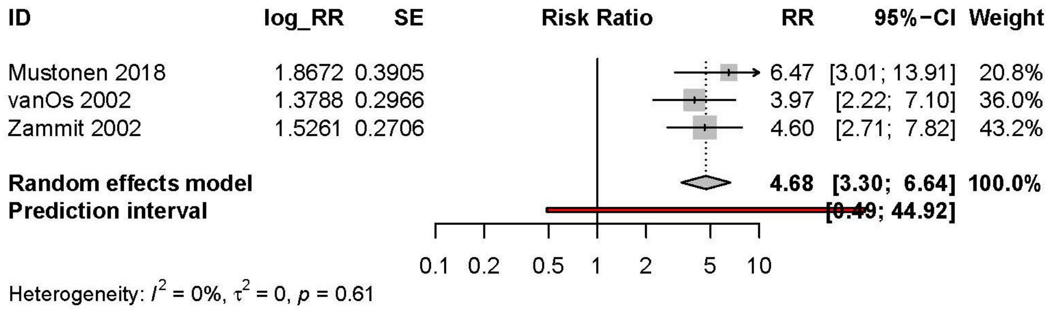

**eFigure 8.** Summary matrix for tobacco use and mood disorders

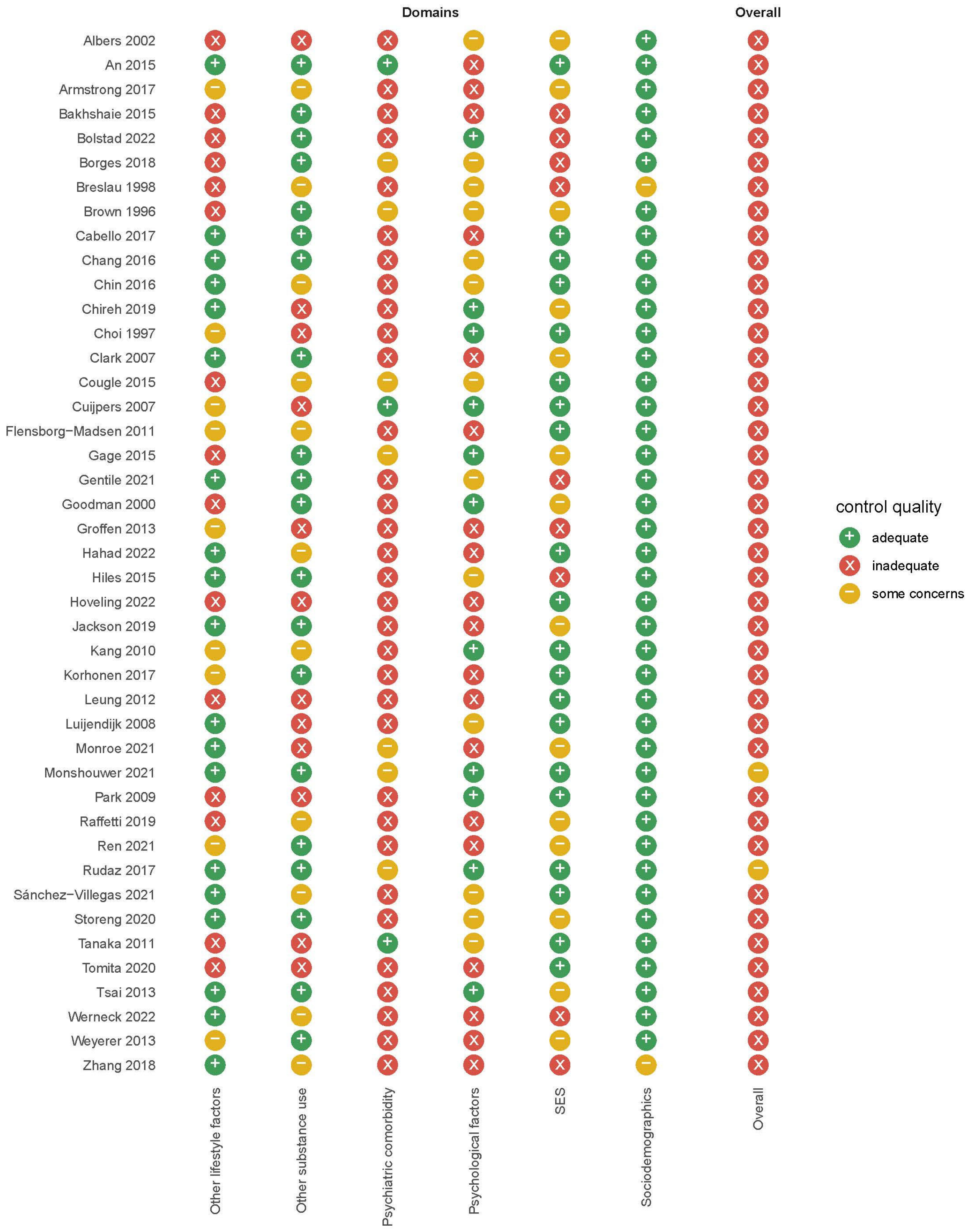

**eFigure 9.** Summary matrix for tobacco use and anxiety disorders

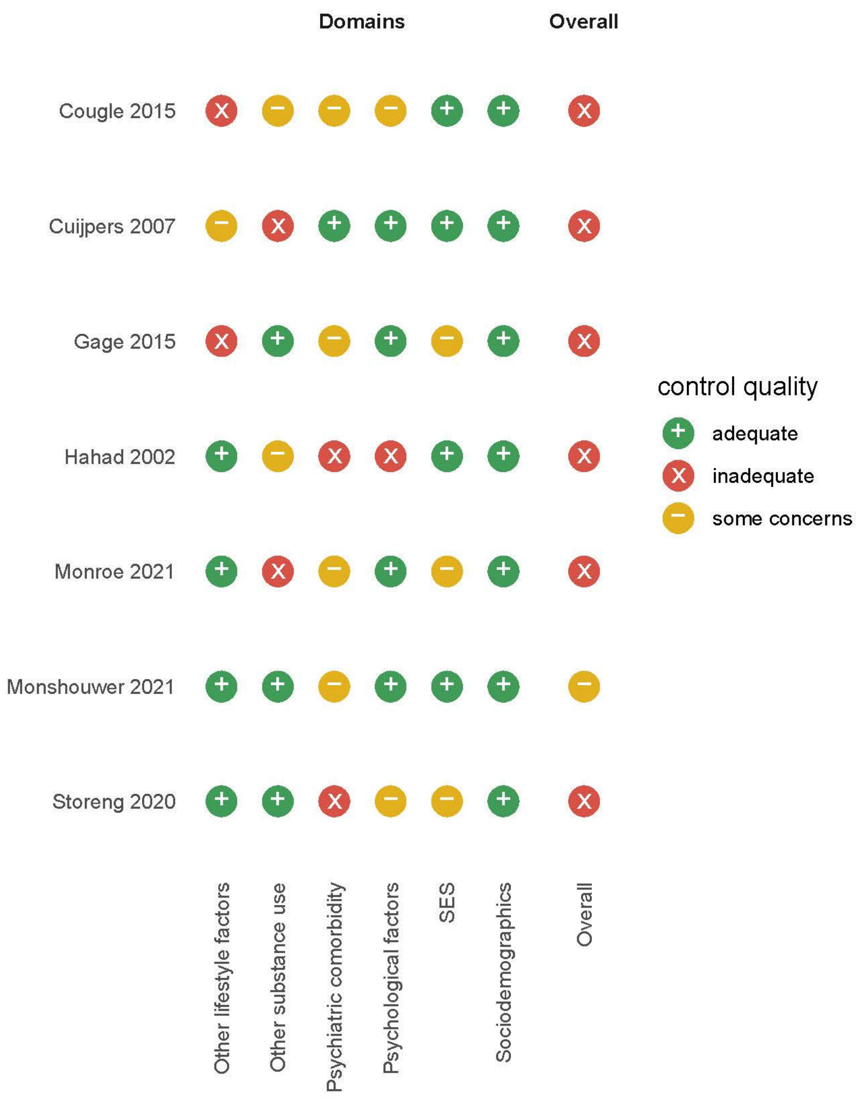

**eFigure 10.** Summary matrix for tobacco use and psychotic disorders

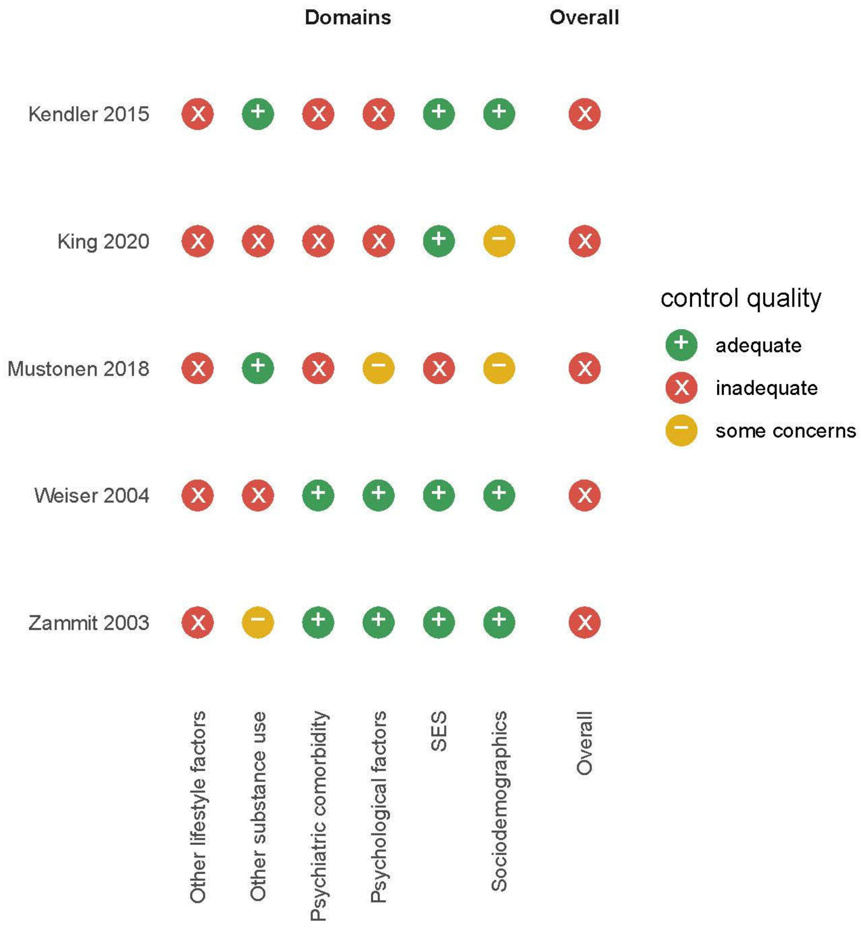

**eFigure 11.** Summary matrix for cannabis use and mood disorders

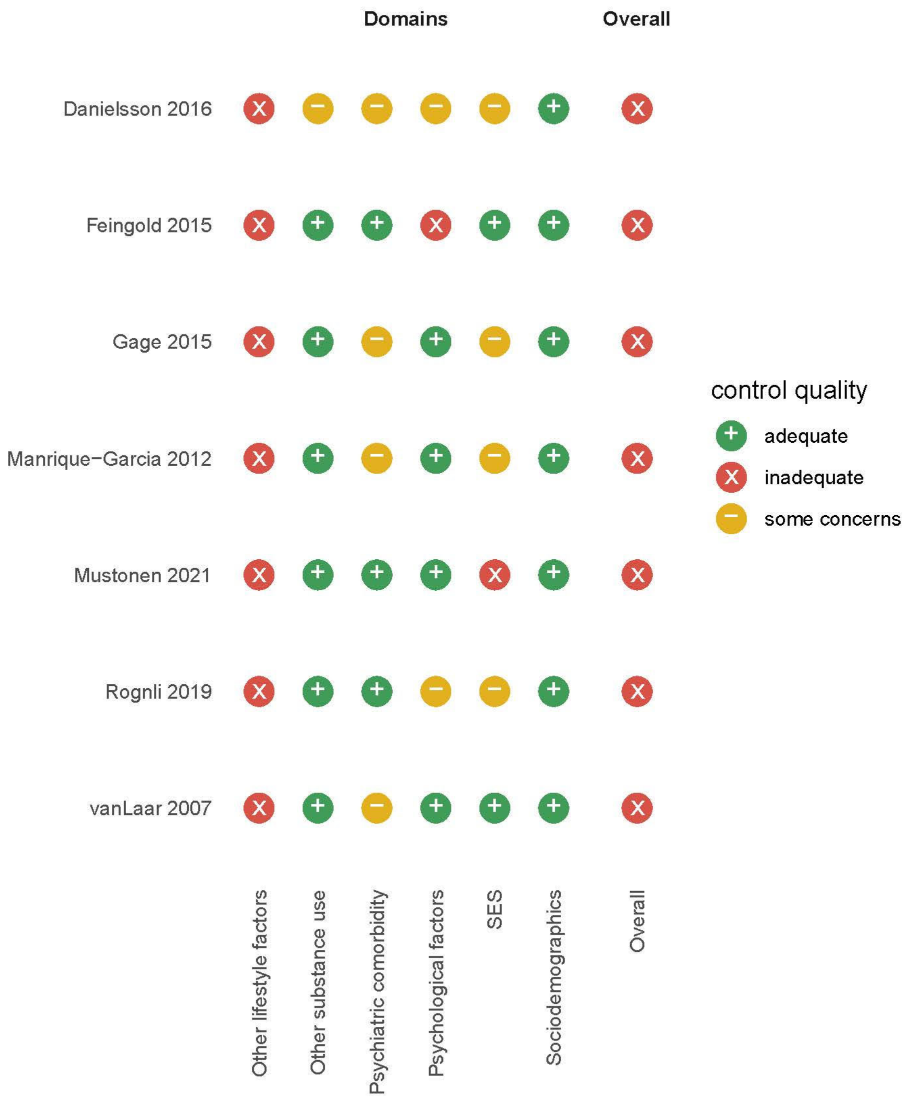

**eFigure 12.** Summary matrix for cannabis use and anxiety disorders

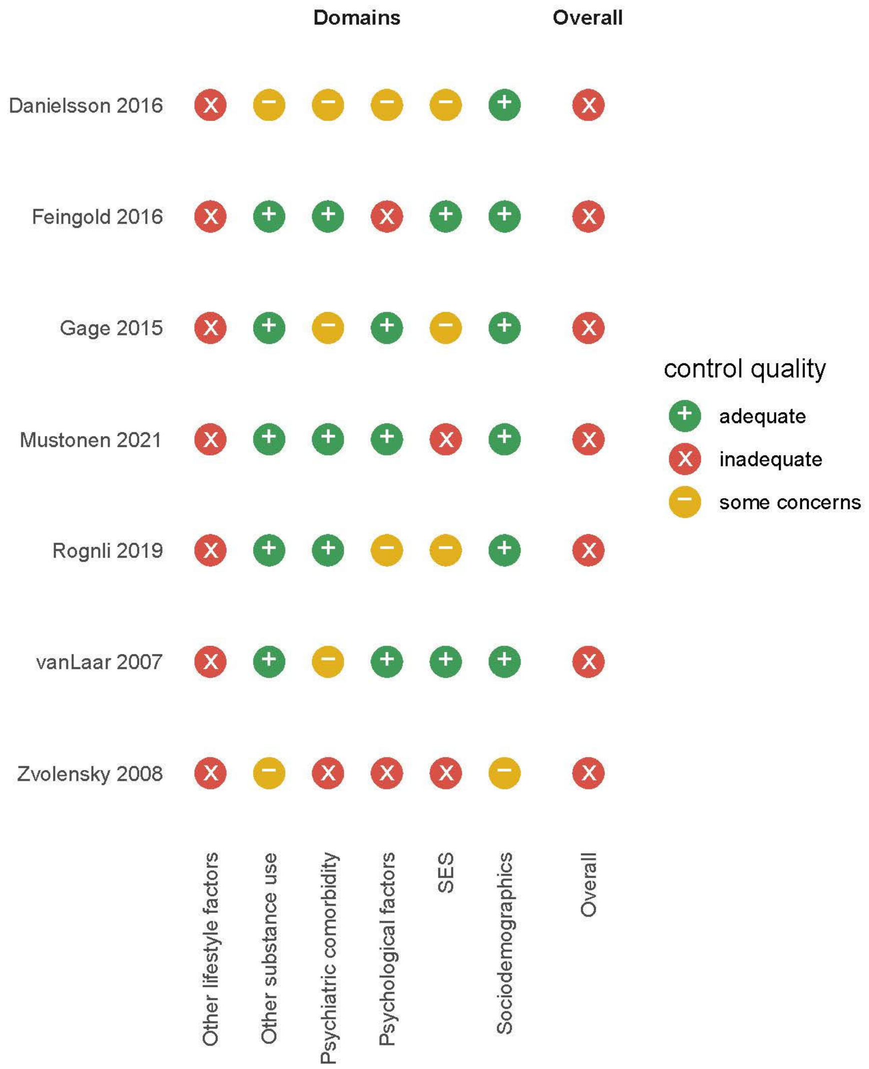

**eFigure 13.** Summary matrix for cannabis use and psychotic disorders

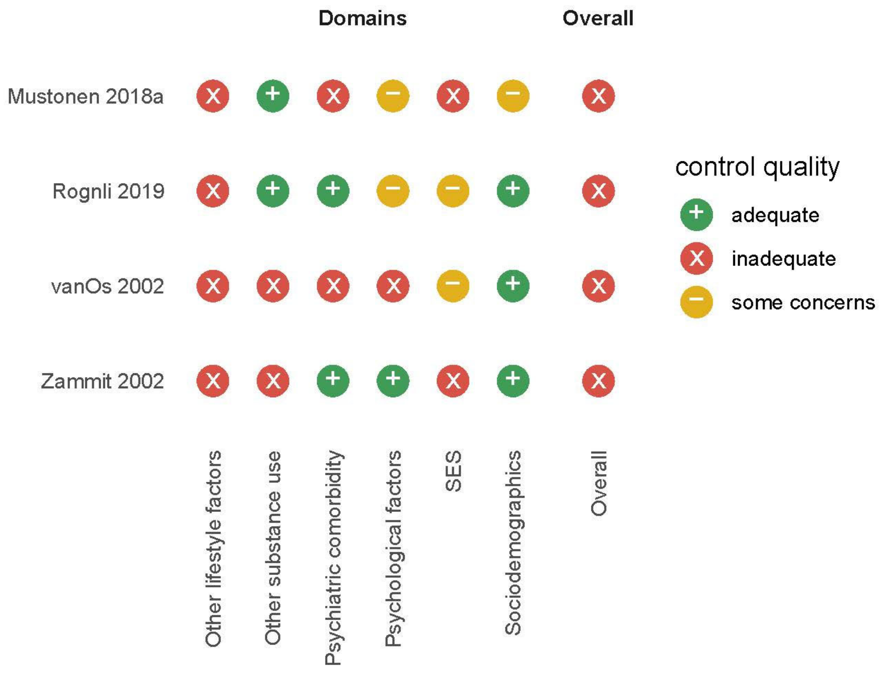

**eFigure 14.** Doi plot and LFK index for tobacco use and mood disorders

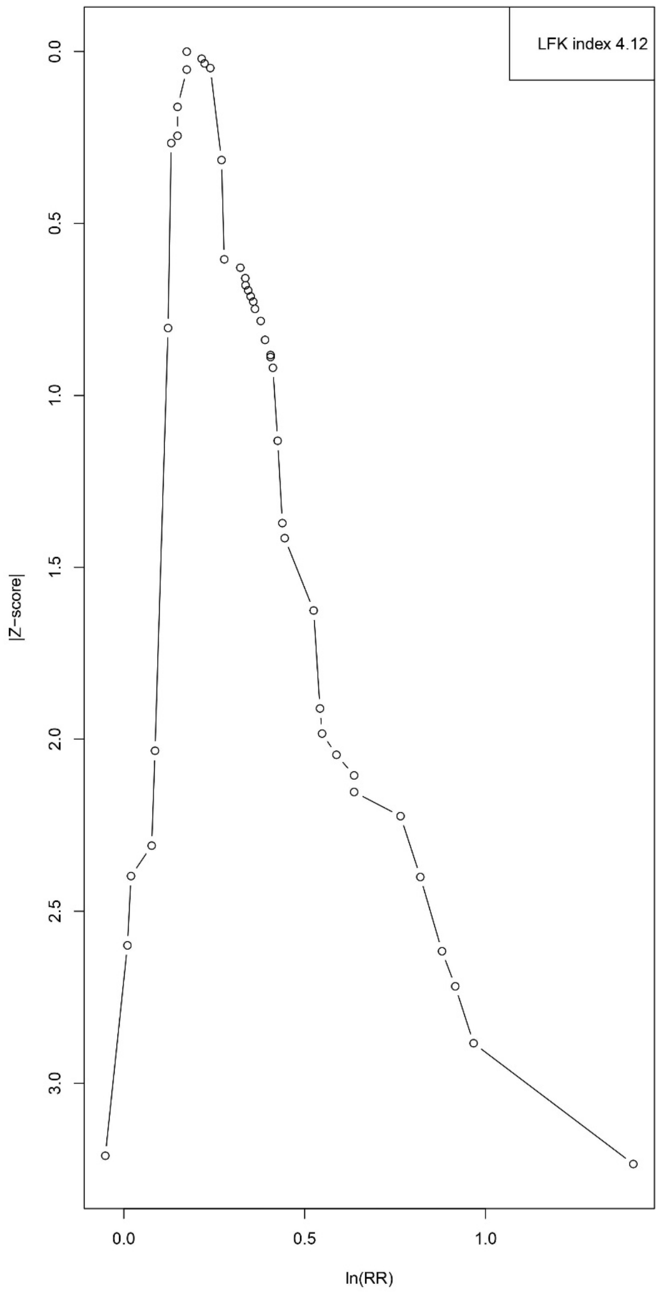

**eFigure 15.** Doi plot and LFK index for tobacco use and anxiety disorders

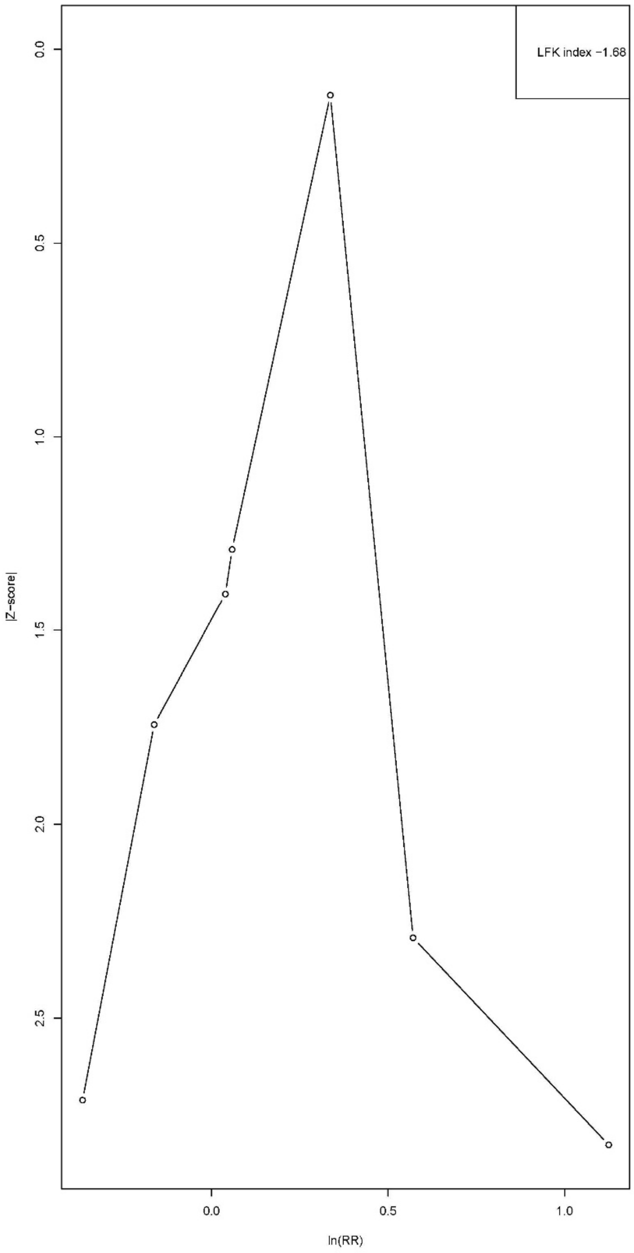

**eFigure 16.** Doi plot and LFK index for tobacco use and psychotic disorders

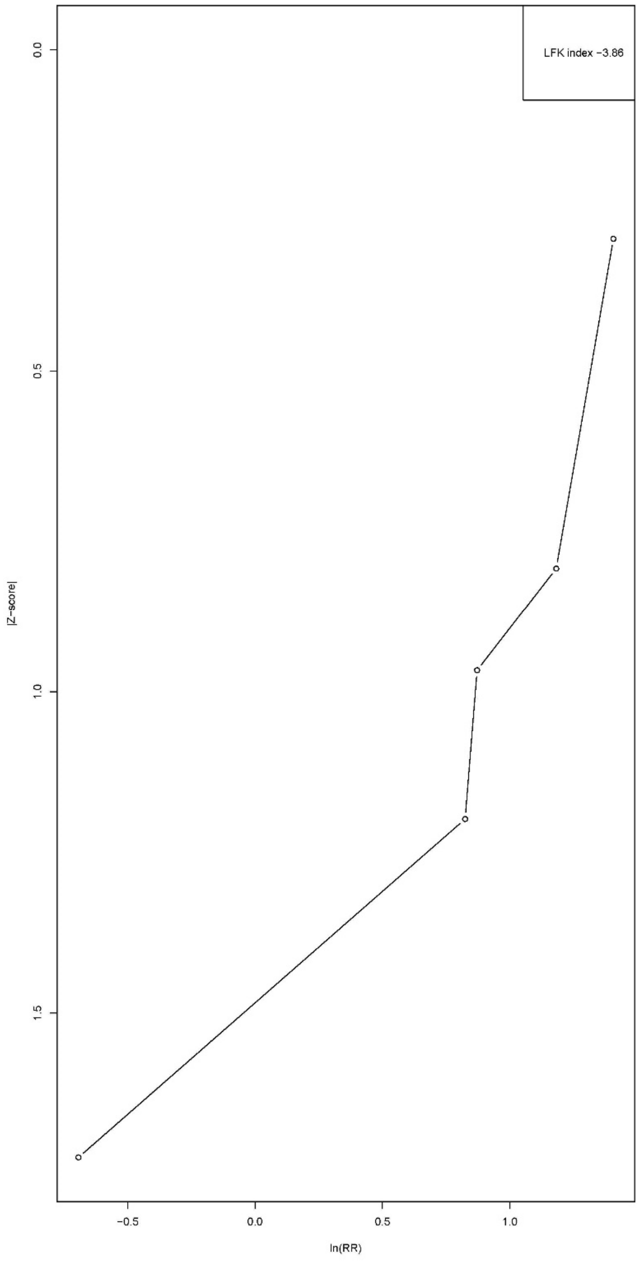

**eFigure 17.** Doi plot and LFK index for cannabis use and mood disorders

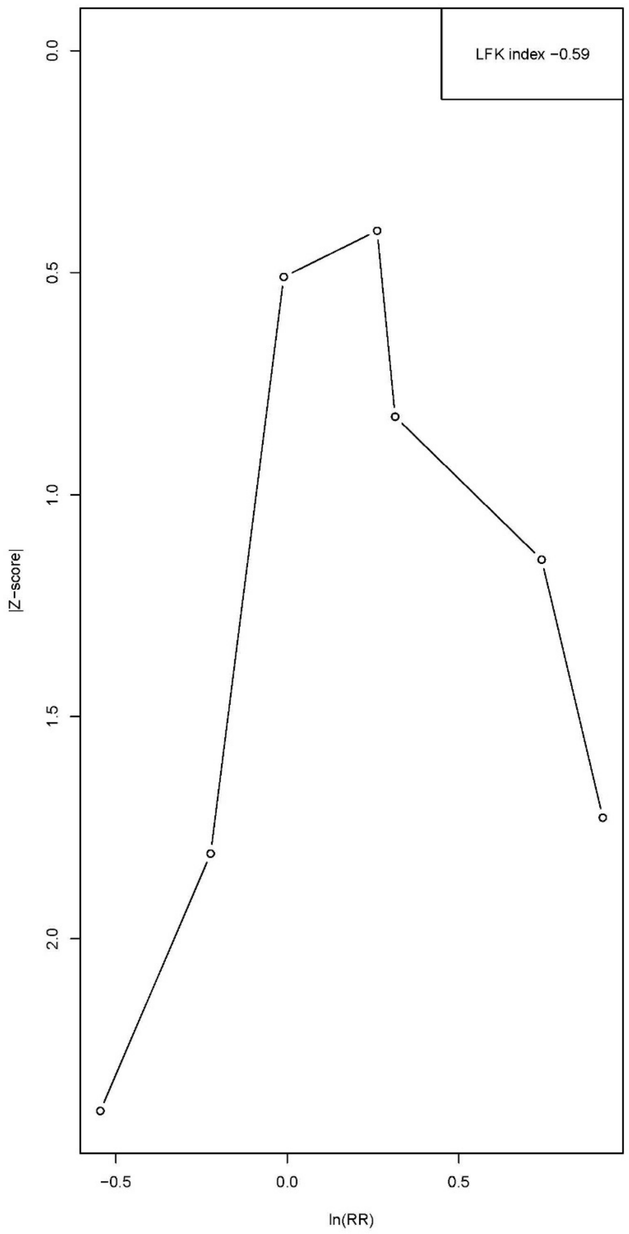

**eFigure 18.** Doi plot and LFK index for cannabis use and anxiety disorders

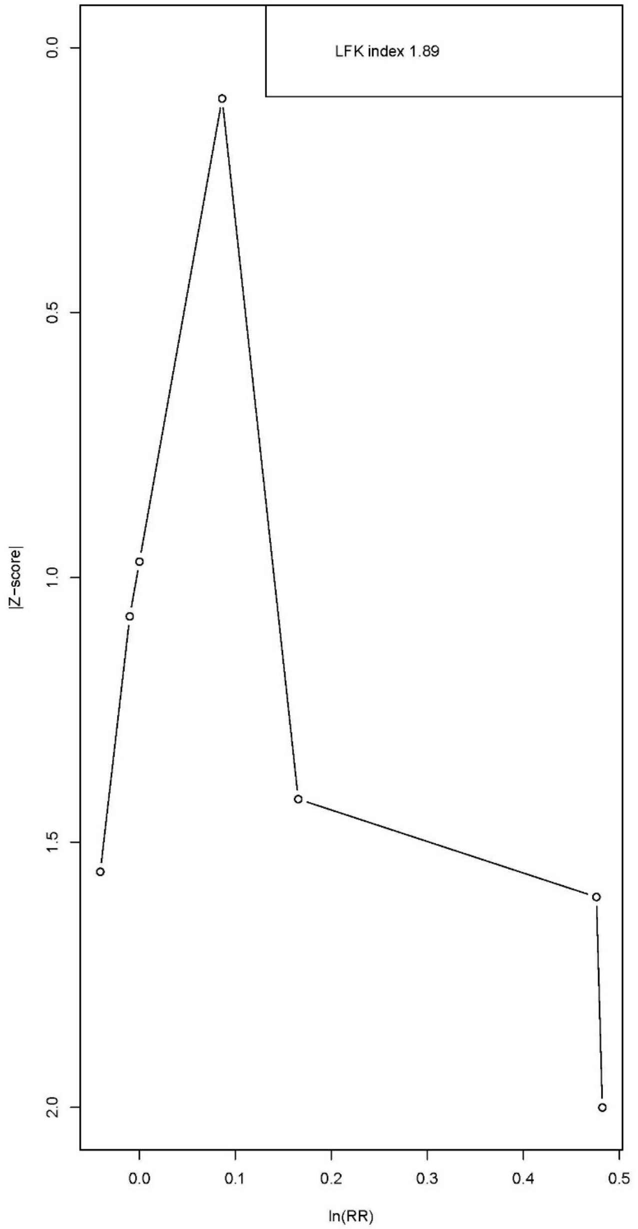

**eFigure 19.** Doi plot and LFK index for cannabis use and psychotic disorders

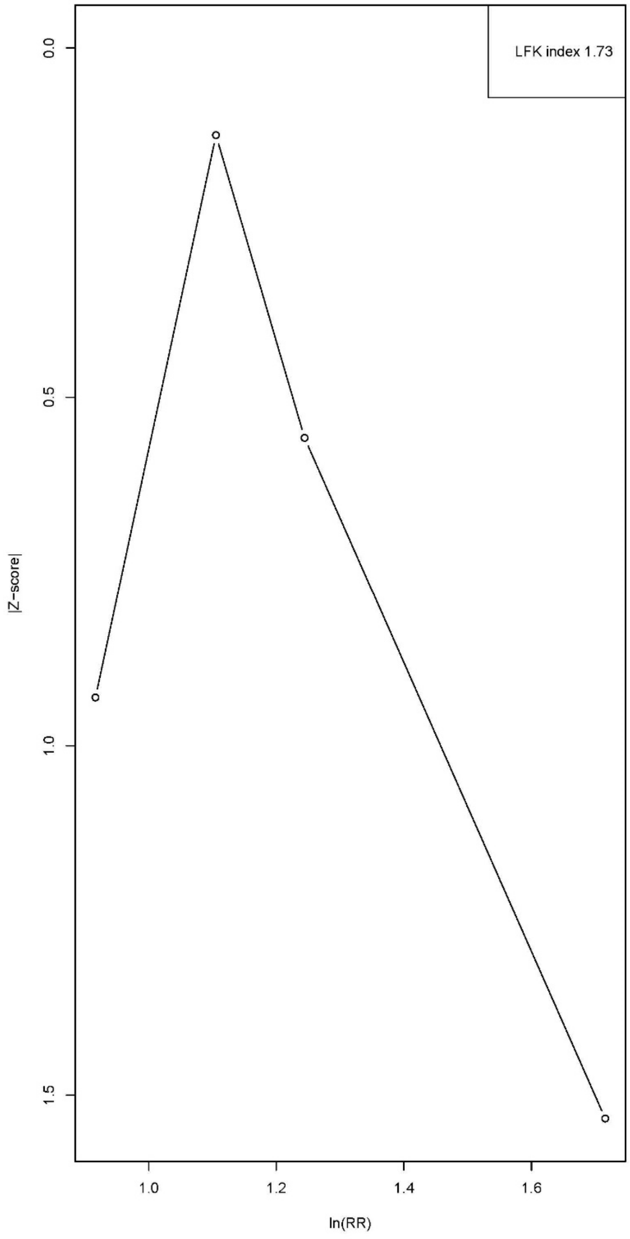
